# Understanding the role and adoption of artificial intelligence techniques in rheumatology research: an in-depth review of the literature

**DOI:** 10.1101/2022.11.04.22281930

**Authors:** Alfredo Madrid-García, Beatriz Merino-Barbancho, Alejandro Rodríguez-González, Benjamín Fernández-Gutiérrez, Luis Rodríguez-Rodríguez, Ernestina Menasalvas-Ruiz

## Abstract

The major and upward trend in the number of published research related to rheumatic and musculoskeletal diseases, in which artificial intelligence plays a key role, has exhibited the interest of rheumatology researchers in using these techniques to answer their research questions. In this review, we analyse the original research articles that combine both worlds in a five-year period (2017-2021). In contrast to other published papers on the same topic, we first studied the review and recommendation articles that were published during that period, including up to October 2022, as well as the publication trends. Secondly, we review the published research articles and classify them into one of the following categories: disease classification, disease prediction, predictors identification, patient stratification and disease subtype identification, disease progression and activity, and treatment response. Thirdly, we provide a table with illustrative studies in which artificial intelligence techniques have played a central role in more than twenty rheumatic and musculoskeletal diseases. Finally, the findings of the research articles, in terms of disease and/or data science techniques employed, are highlighted in a discussion. Therefore, the present review aims to characterise how researchers are applying data science techniques in the rheumatology medical field. The most immediate conclusions that can be drawn from this work are: multiple and novel data science techniques have been used in a wide range of rheumatic and musculoskeletal diseases including rare diseases; the sample size and the data type used are heterogeneous, and new technical approaches are expected to arrive in the short-middle term.

**Highlights:**

- The rheumatology research community is increasingly adopting novel AI techniques
- There is an upward trend in the number of articles that combine AI and rheumatology
- Rheumatic and musculoskeletal rare diseases are gaining from AI techniques
- Independent validation of the models should be promoted

## 1. Introduction

### 1.1. Clinical and technical background

Rheumatic and musculoskeletal diseases (RMDs), for a list of acronyms the reader is referred to *Supplementary Excel File Acronyms*, are defined by the major scientific societies of rheumatology, European Alliance of Associations for Rheumatology (EULAR) and American College of Rheumatology (ACR), as a heterogeneous group of more than 200 diseases and syndromes present in all age segments and in both genders, affecting not only joints, bones, cartilage, tendons, ligaments, nerves, blood vessels, and muscles, but also internal organs [1]. The aetiology and pathophysiology of RMDs can be variable. From a genetic, environmental, postural hygiene, and physical injury perspective, to immunological system disorders, such as inflammation derived from autoimmune responses, infections, or mechanical deterioration of tendons, muscles, and bones. This group of diseases is commonly characterised by chronicity, pain, fatigue, disability, motion dysfunction, and larger female and elder affectation; having a negative impact on life expectancy and quality of life (QoL) of patients. The economic burden associated with RMDs is not negligible and has recently been under the spotlight, as these diseases are responsible for the loss of productivity costs and the costs derived from sick leave and work disability [2]. Concisely, RMDs have a high overall prevalence, a significant economic burden, a deleterious impact on patients’ QoL, and some particularities that hinder patients’ management, making them unique and complex.

From a data science perspective, RMDs also have their own particularities and challenges. To begin with, RMDs data are usually longitudinal, because of the long patient follow-up, which can range from weeks to decades. Therefore, new approaches that seek to take advantage of these data, such as group-based multi-trajectory modeling (GBMT) analyses are emerging [3]. Moreover, RMDs data tend to be heterogeneous and multidimensional. Not only clinical and demographic data but also image, genomic, and -to a lesser extent-sensor data have been used to characterise the patient’s disease, the disease progression or the treatment response and its effect. For instance, the disease progression can be studied with radiological progression measures obtained from medical images. Other data sources and types, such as patient-reported outcomes measures (PROMs) (e.g., health-related quality of life (HRQoL)) are not uncommon in rheumatology [4]. In addition, data from different medical specialities, such as orthopedy, ophthalmology, pneumology, immunology, pharmacy, cardiology, or radiology, often complement the original RMDs data. In this scenario, the dimensionality of the data can increase significantly, especially with genomic data and genome-wide association studies (GWAS). The outpatient setting of most rheumatic clinics also has an impact on the way data are collected. These data often fall under the definition of real-world data (RWD). Although RWD is a valuable source of information, some of its limitations cannot be neglected, such as its less structured nature or the occurrence of biases (i.e., selection bias or informed consent bias), which may require additional processing [5]. In this regard, approaches based on natural language processing (NLP) and topic modelling have been proposed to characterise the evolution of rare diseases in RMDs clinical narratives [6].

The complexity of these data has led to the search for tools capable of modelling and capturing complex statistical interactions and patterns in the data. Researchers have found in artificial intelligence (AI) tools a suitable collection of techniques to extract knowledge from data. These tools have been applied to basic, clinical, and translational rheumatology research studies and to both autoimmune and non-autoimmune RMDs. Some of the supervised learning algorithms employed in rheumatology research studies for regression, classification, and inference are linear, logistic, Poisson regression; regularised linear models (i.e., least absolute shrinkage and selection operator (Lasso), Ridge and elastic net); decision trees (DT); support vector machines (SVM); Bayesian models; naive bayes (NB); k-nearest neighbors (KNN); random forest (RF); neural networks (NN) and boost-based algorithms such as gradient boosted models (GBM) or AdaBoost. These algorithms have been used for a wide range of applications, among them, to predict response to some biological treatments (e.g., anti-tumor necrosis factor (TNF)) [7], disease flare risk based on physical activity [8], and suicide risk in patients with fibromyalgia [9]. On its behalf, unsupervised learning algorithms have played a key role in dealing with high-dimensional data, such as gene expression data and biomarker identification. In this regard, principal component analysis (PCA) has been found to be extremely useful for dimensionality reduction when identifying biomarkers [10], avoiding overfitting and speeding up training time, and t-distributed stochastic neighbour embedding for visualisation [11]. Clustering algorithms have followed multiple strategies: connectivity-based clustering (e.g., hierarchical clustering), centroid-based clustering (e.g., k-means, fuzzy c-means), density-based clustering (e.g., DBSCAN) and probabilistic models (e.g., Gaussian mixture models (GMM)). Moreover, the ability of deep neural networks (DNN) to capture complex patterns have propitiated its use in computer vision and texture analysis tasks such as region of interest identification and segmentation in radiology images. In this regard, DL has been used to quantify the cartilage stage severity in osteoarthritis [12], radiological progression in rheumatoid arthritis (RA) [13] or lumbar spinal stenosis grading [14]. Furthermore, DL has also been employed satisfactorily with structure data from electronic health record (EHR) to forecast clinical outcomes using multiple network architectures, including convolutional neural networks (CNN), recurrent neural networks and long short-term memory networks [15]. Free text and unstructured data from clinical notes have been analysed following a NLP approach, taking advantage of algorithms such as latent dirichlet allocation (LDA) for topic modelling, which has been shown to be useful for disease classification into different meaningful subgroups [16]. Other NLP-based techniques such as *Word2vec* have been employed for rheumatic diseases phenotyping [17]. More recently, novel approaches have reached the RMDs world, such as few-shot learning (FSL) [18], or large language models such as ChatGPT [19].

The present review aims to characterise how rheumatology researchers are employing data science techniques and to study to what extent these techniques have been adopted by rheumatologists. Three main objectives can be distinguished:

1. To study the publication trends in a five-year time window based on the results of a query containing AI and RMDs terms.
2. To conduct a literature review of the original articles published during that five-year period.
3. To discuss the technical, ethical, and regulatory limitations of AI models with a particular focus on rheumatology.

In this review, we intentionally omitted the description and the implications of the different learning techniques and the most widely used algorithms. Since these topics have been covered extensively in previous reviews and usually account for a greater part of the manuscript, we decided to prioritise the description of the different studies. For a better understanding of those techniques, the reader is referred to *Supplementary Excel File Statistical Methods*.

### 1.2. Publication trends

The promising early results of AI techniques have been a decisive step toward its adoption among the different rheumatology research groups. This has been reflected in a growing number of publications, in recent years, in high-impact rheumatology specialised journals. In fact, it has led to the necessity of EULAR to elaborate good-practice recommendations when dealing with big data [20]. When running the Medline query presented in Section 2.1, including the five-year period of the state-of-the-art review (that is, 2017 to 2021), the upward trend can be easily appreciated. The number of published articles has grown by almost 300% from 2017 to 2021. See Figure 1.

**Figure 1:**
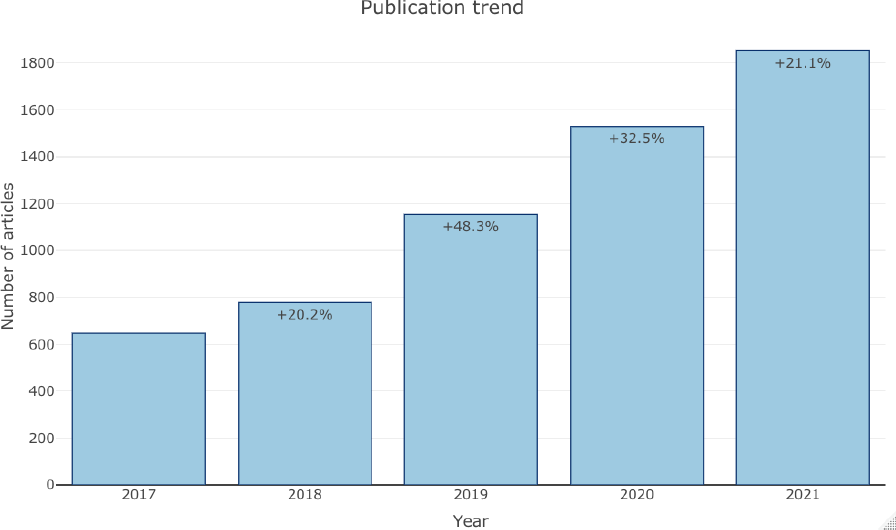
Publication trends in Medline when running the query presented in Section 2.1

### 1.3. Past reviews

Over the last few years different review and recommendation articles, in which AI and RMDs fields interact, have emerged.

In 2017, a review article addressed the ability of machine learning (ML) algorithms to discriminate between chronic pain patients (e.g., chronic pelvic pain, fibromyalgia, low back pain) and healthy controls [21].

In 2019, a systematic literature review informing EULAR recommendations for the use of big data and artificial intelligence in RMDs came to light [22], as well as a compilation of studies that covered the feasibility and clinical utility of ML to stratify patients and predict treatment response [23]. A review covering the methods used in the analysis of rheumatic and musculoskeletal clinical data was presented in [24]. Throughout the year, other review articles dealing with specific diseases were published: prediction models for osteoarthritis (OA) [25], the role of ML and cardiovascular risk assessment in RA patients [26], detecting early RA [27], the application of ML in axial spondyloarthritis (axSpA), and prediction of osteoporosis [28].

A year later, in 2020, the EULAR points to consider for the use of big data and artificial intelligence in RMDs were presented [20], along with two reviews of ML methods applied to rheumatic diseases [29, 30]. That same year, an introduction to the ethical issues of big data [31], as well as a review of the use of AI in rheumatology imaging [32] were available.

In 2021, the review article titled *’An introduction to machine learning and analysis of its use in rheumatic diseases’* [33] provided a distinguished overview of the current situation of ML techniques applied to rheumatology, with a description of the most commonly employed algorithms, as well as, examples of ML applications in rheumatology. Another review, with a strong educational purpose, evaluated the relevance of data science to the field of rheumatology [34]. In addition, a scoping literature review of ML approaches to improve disease management of patients with RA was published at the end of the same year [35]. Particular interest was also given to the use of ML solutions for osteoporosis in [36]. The applicability of AI in the management of RMDs, with data collected from wearable activity trackers, was examined in [37].

In 2022, some authors provided an outline of ML applications in musculoskeletal histopathology [38], and reviewed the ML methods applied to OA research [39], with a special emphasis on magnetic resonance imaging (MRI) [40]. Other authors addressed the use of three specific ML algorithms including logistic regression, in the diagnosis of rheumatic illnesses [41]. Another review, published in the middle of the year, discussed recent technologies and innovations that are expected to benefit clinical practice in the early 2030s, with regard to big data [42]. A narrative review of ML in RMDs for clinicians and researchers was published by Nelson AE et al. [43]. Furthermore, another narrative review [44] covered the applications of ML in systemic sclerosis (SS). A review of AI and deep learning (DL) [45] attempted to highlight the relevance of these techniques in the near future of the field of rheumatology. Finally, a review explored the opportunities and challenges of using RWD data focusing mainly on the electronic health record (EHR) data to advance clinical research in rheumatology [5].

The increasingly frequent appearance of these kinds of articles (i.e., review and recommendation) gives an idea of the general adoption of AI and ML in RMDs and in other medical specialities [46]. Unlike previous assessments, in the present review, technical details assume a secondary role and the writing is instead oriented towards facilitating a diverse range of original research examples that may be useful to the reader as a starting point for conducting its research in the field of AI.

## 2. Materials and Methods

A literature search was conducted to identify publications related to RMDs in which data science techniques played a relevant role. Firstly, results from Medline, Scopus and Web of Science were extracted. Lastly, specific searches were performed in the main rheumatology journals using their integrated search engines. The boolean operators AND and OR were used to streamline the procedure. The selected articles had to be indexed in PubMed (i.e., with a PubMed Identifier (PMID)) at the time of the search (that is, the index dates were used instead of the publication dates).

### 2.1. Search in Medline

The search consisted of two stages. In the first stage, articles published from January, 1^st^ 2017 to June, 17^th^ 2020 were retrieved. In the second stage, articles published from that date to February, 22^th^ 2021, were collected.

The Medline search strategy included a combination of keywords and Medical Subject Headings (MeSH) terms. Due to the large number of keywords related to RMDs and AI and potential omissions, the search strategy did not specify, for example, a concrete type of disease or a concrete AI technique or algorithm. The keywords and MeSH terms from Table 1 were used to build the Medline query:

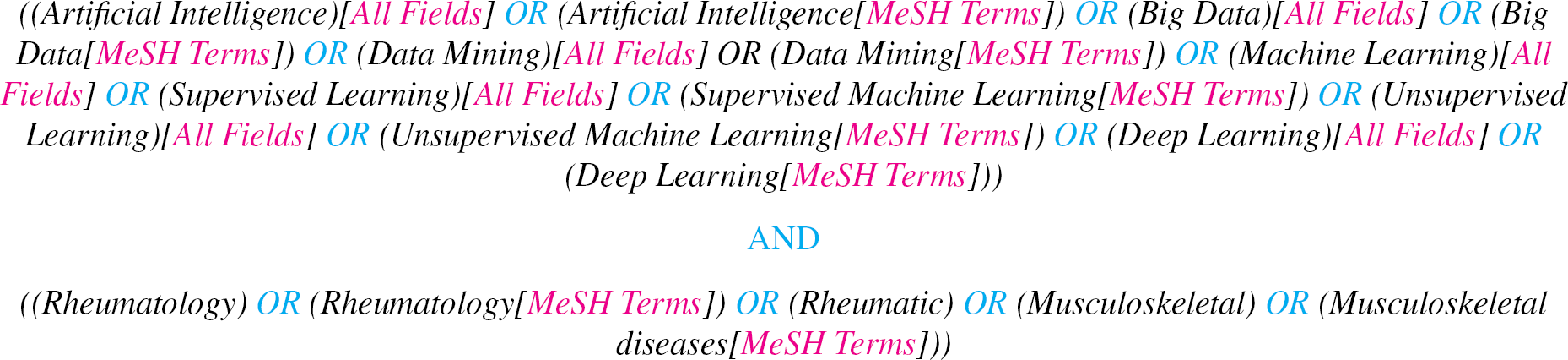

**Table 1.**
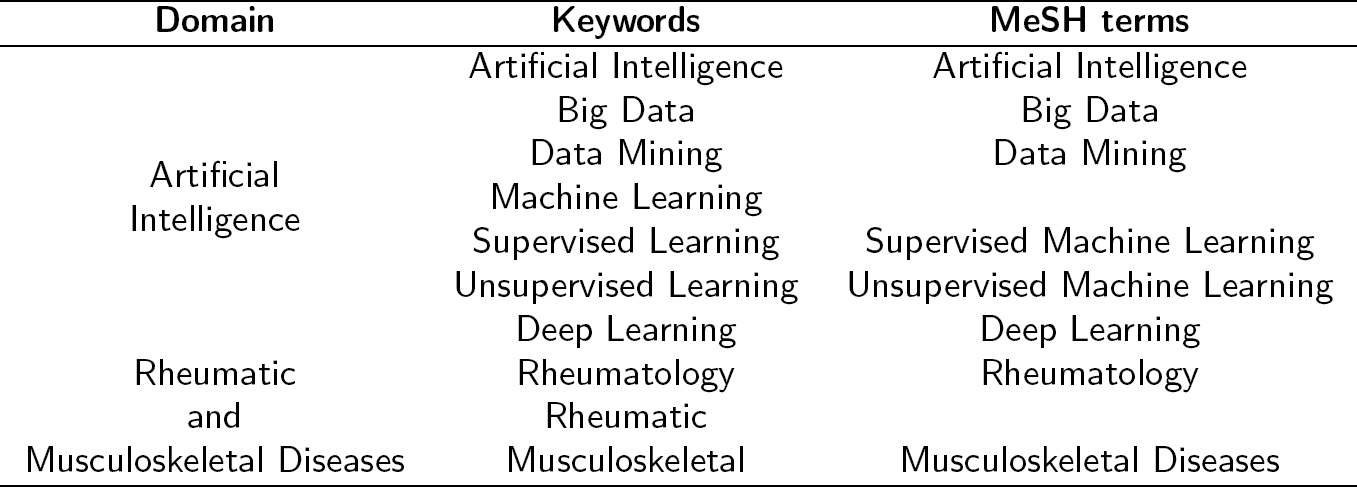
Keywords and MeSH terms used in the Medline search

### 2.2. Search in Scopus

The Scopus query was restricted to the article title, keywords, and abstract. Only results from January, 1^st^ 2017 to February, 22^th^ 2021 indexed in Medline (i.e., with a PubMed identifier (PMID)) were included. The query performed in Scopus was:

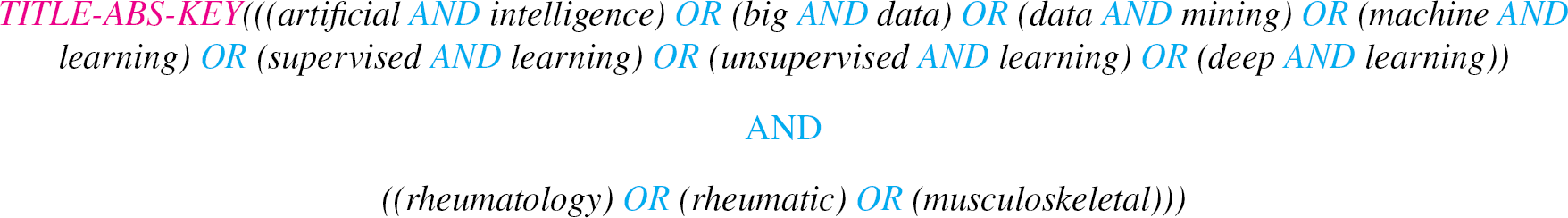

### 2.3. Search in Web of Science

The Web of Science search was similar to the Scopus search. The query performed was as follows:

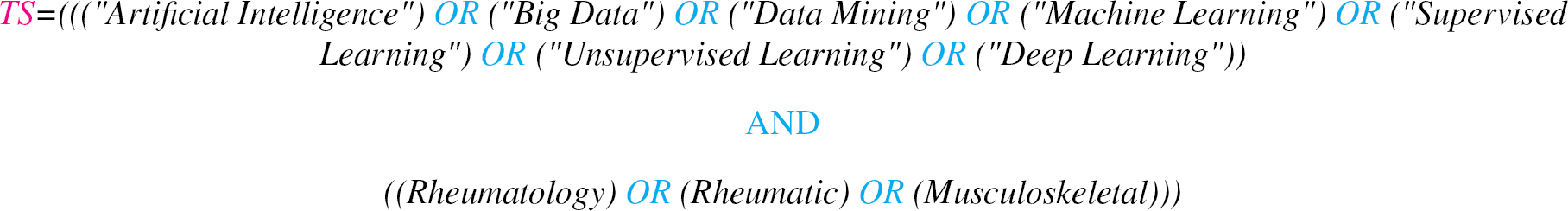

### 2.4. Search in rheumatology journals

Articles published in Q1 and Q2 rheumatology journals (according to 2019 Journal Citation Reports) were retrieved, excluding those classified as ‘Congress’, ‘Abstract’ or ‘Miscellaneous’. The decision to limit this search to Q1 and Q2 journals was made to ensure that the articles retrieved had a high impact. This restriction only applies to this search. The journals included in this search were: *Nature Reviews Rheumatology, Annals of the Rheumatic Diseases, Arthritis & Rheumatology, Rheumatology, Therapeutic Advances in Musculoskeletal Disease, Osteoarthritis and Cartilage, Seminars in Arthritis and Rheumatism, Arthritis Research & Therapy, Arthritis Care & Research, Current Opinion in Rheumatology, Current Rheumatology Reports, Joint Bone Spine, Rheumatology and Therapy, Journal of Rheumatology, and Clinical and Experimental Rheumatology*.

The query used in the search engine of the different journals was:

> “Machine Learning”

## 3. Literature review

The number of records identified through the database search steps described in the previous sections was 4,325. Table 2 the initial number of articles retrieved from the different sources is shown. From this point on, different exclusion criteria were applied. Figure 2, shows the exclusion and inclusion criteria.

**Table 2.** Number of articles retrieved and with PMID in the different search engines

| Source | Number of records identified | PMID |
| --- | --- | --- |
| MedLine | 2,071 | 2,071 |
| Scopus | 1,282 | 492 |
| Web of Science | 731 | 433 |
| Rheumatology journals | 241 | 241 |

**Figure 2:**
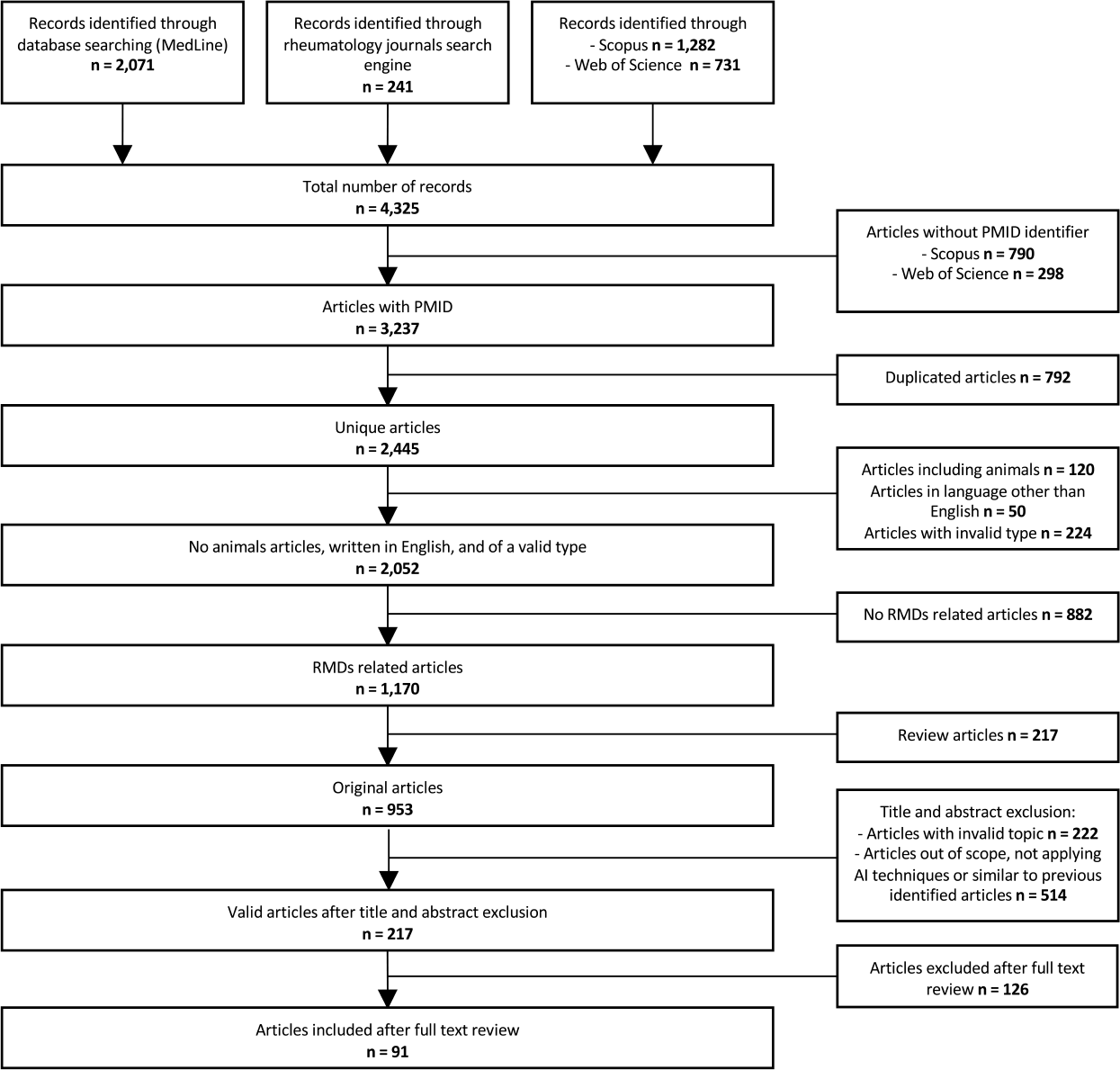
State of the art inclusion and exclusion criteria

To sum up, we included unique articles, with a PMID, written in English, without animal involvement. On the other hand, we excluded articles identified as *analysis, annotation, biography, case report, cohort profile, comment, correspondence, correspondence response, editorial, editorial comment, epilogue, erratum, guideline, letter, meta-analysis, methodology, news, news & analysis, opinion, overview, perspective, protocol, research highlight, research report, response, video article*.

We also removed articles not related to RMDs; articles covering the following topics: *medical surgical procedures such as arthroplasty or arthroscopy, biomechanics, cancer (excluding lymphoma), muscle and bone malignancies, education, force simulation, gait, image generation, reconstruction, joint replacement, orthopaedics, radiology, rehabilitation and other non-pharmacological interventions, robotics, simulation and surgery*; articles out of scope, not applying AI techniques or similar to previously identified ones.

The articles remaining after the elimination of duplicates (i.e., exclusion criteria 2) are shown in *Supplementary Excel File Unique Articles*. Finally, the article title, the identifiers (i.e., DOI and PMID), the journal, the publication year, the disease, the algorithms/techniques, the programming language, the number of patients, the validation type, the objective, and the authorship information are available in the *Supplementary Excel File Included Articles*.

### 3.1. Classification of topics and predictors

Hereafter, a review of the rheumatology research articles in which data science techniques were used, over a five-year period, 2017-2021, is conducted. Different categories have been proposed to classify research articles. For instance, the scoping review presented in [35] suggested three main categories and eight subcategories to describe RA studies in which ML was applied. On the other hand, authors in [33] suggested a different categorisation when describing ML techniques applied to RMDs, Table 3.

**Table 3.** Categorisations proposals made in other reviews

| Kedra et al. [35] |  | Kingsmore et al. [33] |  |
| --- | --- | --- | --- |
| Diagnosis | EHR | Patient classification | EHR and clinical data |
|  | Biological samples |  | Imaging and biometric data |
|  | Imaging and image recognition |  | Urinalysis, flowcytometry, and genomics |
|  | Sensors | Risk classification and outcome prediction |  |
| Monitoring | Disease | Predicting treatment response or candidates for treatment |  |
|  | Comorbidities |  |  |
| Prediction | Response to treatment | Patient clustering to determine disease subtypes |  |
|  | Outcomes |  |  |

As shown, there is no standardised way to classify medical research articles in which data science approaches are used. The proposal followed in this review tries to strike a balance between resolution and significance. Hence, the articles are categorised into six main topics, based on the intended use of the techniques employed, and three subcategories, based on the types of the variables; Table 4. When a study could be classified into more than one category, it was assigned to the most complex category (i.e., the design of the study is becoming increasingly sophisticated) according to the order shown in Figure 3. This figure shows a scheme of the different categories proposed and how they relate to each other. If a study combines predictors that fall into more than one of the subcategories, the article is classified according to the most complex variable (i.e., the difficulty associated with acquiring or processing the data). Figure 4 shows an outline of the different subcategories. A full description of the categories and subcategories proposed can be found in the *Supplementary File Categories*.

**Table 4.** Proposed classification of topics with six main categories and three subcategories, depending on the predictor type

| Categories | Subcategories |
| --- | --- |
| Disease identification and prediction | Clinical, demographic and biomarker data accessible within routine clinical practice |
| Disease classification |  |
| Patient stratification and disease subtype identification | Complex molecular biomarkers |
| Disease progression and activity |  |
| Treatment response | Image |
| Predictors identification |  |

**Figure 3:**
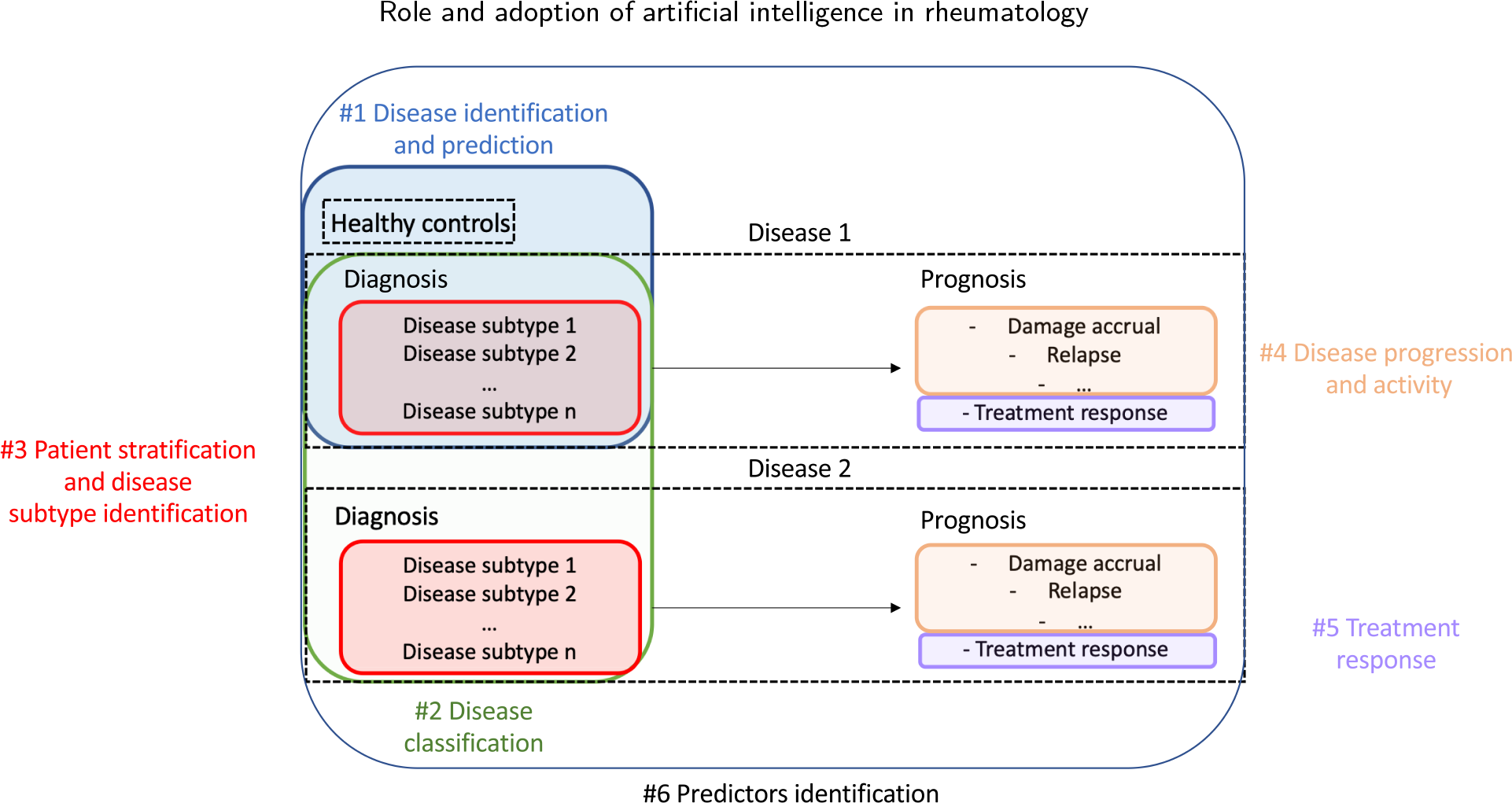
Proposed classification into six main categories

**Figure 4:**
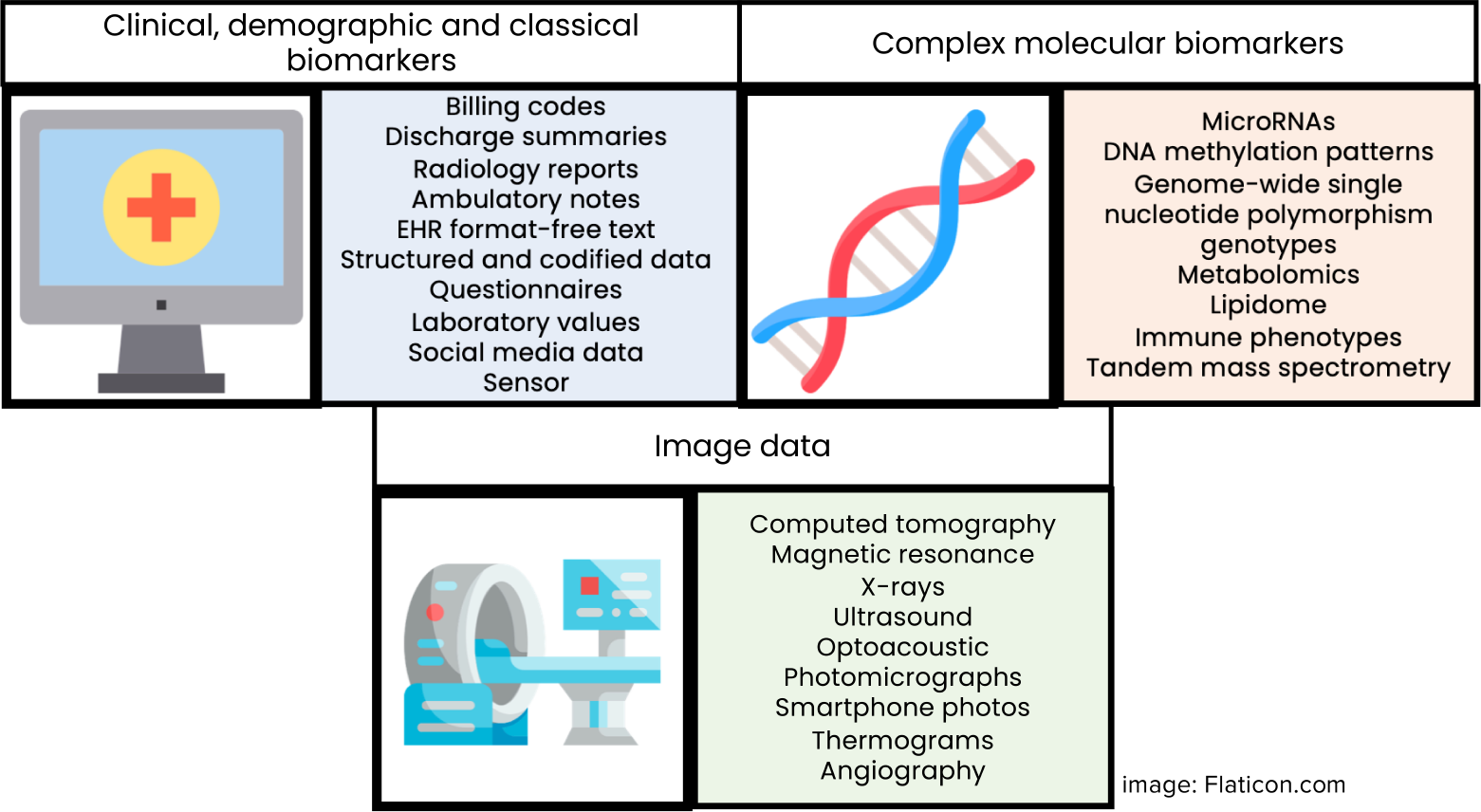
Data sources considered in each subcategory. For more details the reader is referred to *Supplementary File Categories*

### 3.2. Disease identification and prediction

This section describes studies that attempt to classify patients with concrete RMDs versus controls (i.e., healthy patients or patients without the disease of interest), or that attempt to identify patients with a particular condition using EHR data, generally with the aid of NLP.

#### 3.2.1. Clinical and demographic

The consequences of delayed diagnosis and treatment extend beyond patient well-being (e.g., increased flares and organ dysfunction) and can cause significant economic burdens at different levels. This can be exacerbated by the lack of diagnostic criteria or outdated disease definitions. Under this assumption, early diagnosis predictive models have been developed for ankylosing spondylitis (AS), after applying mutual information for feature selection [47]. In this study, the authors trained different algorithms such as SVM, RF or GBM, and compared their performance against more traditional methods like linear regression and clinically-based models. The purpose of this study was the early identification of AS patients based on medical and pharmacy claims history. Of 228,471 patients, without a history of AS, the ML model predicted that 1,923 patients would develop AS. However, only 120 of the predicted patients developed AS. A total of 1,242 out of 228,471 finally developed the disease. While the linear regression model accounted for a higher Area Under the Curve (AUC), 0.71, compared to the ML model, 0.63, the false positive rate and positive predictive value (PPV) values were 4.67% and 2.55%, for the linear regression and 0.79% and 6.24% for the ML model. As predictors, three groups of categories were used: diagnosis (e.g., unspecified inflammatory spondylopathy, sacroiliitis), procedure (e.g., HLA typing single antigen, radiological examination sacroiliac joints) and prescription (e.g., propoxyphene, sulfasalazine).

In recent years, different research groups have been working to identify axSpA patients in large datasets with both structured and unstructured data. As some authors stated, identifying axSpA patients for observational studies is a challenge, partly due to the lack of codification for the different disease phenotypes. Historically, axSpA International Statistical Classification of Diseases and Related Health Problems (ICD) codes and manual chart review have been used for identification. Recently, three models to identify patients with axSpA have been proposed by a US research group [48]. These models considered a different number of unique predictors from 16 to 49, including concepts extracted with the help of NLP techniques (e.g., sacroiliitis, HLA-B27), demographics (e.g., race, ethnicity), laboratory (e.g., CCP), healthcare utilisation (e.g., number of rheumatology visits), ICD codes and comorbidity index variables (i.e., Charlson Comorbidity Index). The authors used the RF algorithm to quantify an axSpA risk score and concluded that the most complete algorithm, which included 49 variables, had the highest overall performance, obtaining a 0.65 F1 score.

Previously, a research group from the UK [49] had applied unsupervised and supervised algorithms, also using concepts extracted after applying NLP, to accurately identify axSpA patients in an EHR from an enriched axSpA cohort. In this regard, the authors generated a list of candidate axSpA concepts with the surrogate assisted feature extraction method and selected the six most informative concepts with Lasso (e.g., number of ‘AS’, sacroiliitis, spondylitis concepts), after processing the clinical reports with NLP. Furthermore, they tested an unsupervised implementation, multimodal automated phenotyping, which combined information from three key domains (i.e., ICD codes, NLP concepts and healthcare utilisation). This last approach had the highest sensitivity and PPV, 0.78 and 0.80, respectively, with an AUC value of 0.92. Lastly, the authors concluded that axSpA patients could be accurately identified in EHR by incorporating narrative data.

Observational studies on rare diseases are often limited by sample size. Therefore, automatic methods to accurately identify patients suffering from these diseases in the EHR are key to maximising the number of cases and achieving enough statistical power to draw valid conclusions. With this in mind, authors in [50] aimed to implement different approaches to broadly capture SS patients from an EHR, using meaningful variables such as ICD codes (e.g., 710.1, M34*), laboratory data (e.g., positive anti-nuclear antibody (ANA)) and keywords (e.g., Raynaud’s phenomenon). Accordingly, the performance of rule-based, classification and regression trees, and RF algorithms was compared. Although the authors showed that ML-based algorithms were not the highest performing ones, in terms of PPV and F-score, 0.84 and 0.88 respectively, compared to rule-based algorithms, 0.90 and 0.91, they were not as time-consuming to develop and validate as the last ones. In addition, the authors highlighted some potential advantages of ML algorithms: they do not require specific domain knowledge and can be automatically tuned to easily identify the optimal model parameters.

Given the sheer volume of text notes in the EHRs of some centres, identifying patients who meet certain criteria can be a daunting task. In lupus research, studies to identify patients in EHR and to predict risk probabilities have been carried out. For instance, authors from [17] presented a study in which they used different text classifiers techniques for identifying systemic lupus erythematosus (SLE) patients while reviewing common and more advanced NLP approaches (e.g., bag-of-words (BOWs), Concept Unique Identifiers (CUIs), *Word2vec*). On this basis, the authors showed a pipeline with three clearly separated pathways and two main split nodes, based on how the textual data were transformed and processed into features. If the final word representation was based on word/CUIs frequency in each clinical note (i.e., data resulting from applying BOWs and CUIs), a RF classifier was trained for extracting the variable importance and for feature selection; followed by NN, RF, NB and SVM classifiers. On the other hand, if the word representation was based on vectors for each word presented in a document (e.g., *Word2vec*), bayesian inversion was conducted. When benchmarking the results from the test set, the AUC scores ranged from 0.80 to 0.99. Although the novelty method proposed (i.e., *Word2vec* bayesian inversion) was not the best-performing one, authors concluded that it is promising for identifying patients, since it has fewer dependencies, lower testing, and is more scalable than previous approaches. Among the stemmed terms identified through the variable importance method, “C3”, “sle”, “graviti”, “sole” and “phurin” were the most relevant ones.

Changes in clinical practice, disease management, or the emergence of new therapeutic drugs and procedures can constitute a source of heterogeneity, making it difficult for algorithms to generalise. Studies that assess the temporal validity of predictive models, developed years ago, over time have been carried out [51]. With a seven-year difference, the authors compared the performance of a RA logistic regression algorithm, trained in 2010, against a new logistic regression model that included new ICD-10 codes (e.g., M05.x) and RA treatments (e.g., adalimumab, golimumab). ICD codes, prescription, laboratory results (e.g., rheumatoid factor, anti-cyclic citrullinated peptides (CCP)), and NLP concepts extracted from narrative data and stored in a new electronic medical record were used. The authors evaluated the performance of the ancient algorithm in the updated data, as well as the performance of the updated algorithm using the same model coefficients as the first one while incorporating new variables. The researchers finally concluded that the performance of the old algorithm was similar to the new one when validating with updated data.

To reduce the time-consuming manual chart review, and to minimise the inclusion of false positives when using standardised billing codes, Leiden and Erlangen researchers developed a workflow for building a ML algorithm capable of accurately identifying patients with RA from clinical narratives using NLP [52]. A pipeline of five steps containing word segmentation and trigram aggregation, lowercase conversion, stopwords removal, word normalisation with lemmatisation and vectorisation with term frequency-inverse document frequency was applied for processing the free text notes. Then, NB, NN, RF, SVM, DT, GBM classification models were trained for obtaining a probability score of rheumatoid arthritis. The final aim of this research was to implement a broadly applicable workflow to equip centres with their own high-performing algorithm. Therefore, this research study placed a greater emphasis on the technical aspects. The most relevant terms, identified by the best-performing classifier, SVM, include “ra”, “mtx”, and “erosive”.

Medical imaging techniques can be one of the most reliable methods to diagnose certain pathologies. However, image acquisition equipment is not always present in medical facilities, or its availability is limited or not guaranteed. A study for the early OA detection, using exclusively clinical (e.g., hypertension, diabetes mellitus, dyslipidemia, stroke), demographic (e.g., gender, age, region, marital status, education) and lifestyle data (e.g., body mass index (BMI), obesity, alcohol intake, self-reported status of health) has been conducted in [53]. In this study, the authors highlighted the relevance of building an OA classification model when imaging data are missing. The 24 initially proposed features, from 5,749 subjects, underwent through PCA, and a DNN was trained with the resultant components. The authors achieved an AUC value of 0.77.

The importance of topic modelling and NLP for feature extraction and text structuring in RMDs was shown in the context of pseudogout disease [54]. As the authors reported, identifying pseudogout in large datasets can be challenging due to several factors: its incidence is not well-characterized and there exists a lack of specific billing codes. Almost ten million narrative notes from more than 50,000 patients were processed, and 73 Unified Medical Language System codes were applied to count mentions of those concepts. Then, two filters based on billing codes and NLP concepts, followed by a random selection of patients (n = 900) were applied. Subsequently, with the aid of a novelty topic modelling algorithm, sureLDA, followed by penalised regression, a pseudogout propensity score was estimated and regression models were computed to predict the probability of pseudogout. Sixty-two input features including ICD codes, laboratory codes, and NLP counts were used as the input of the sureLDA algorithm. The regression models included the sureLDA score, the counts of the NLP concept “pseudogout” and the presence of synovial fluid crystal analysis. Finally, five different algorithms, including and excluding the topic modelling score were compared. The PPV, and the AUC values ranged from 0.79 to 0.81 and from 0.70 to 0.86 when using the topic modelling approaches.

#### 3.2.2. Molecular biomarkers

The diagnosis of certain diseases, such as JIA, depends primarily on clinical presentations, which can be highly variable between patients. On the other hand, deep immunophenotyping techniques (e.g., flow cytometry) provide a comprehensive characterisation of the immune cells of patients with an autoimmune disease. In this context, RF, SVM and artificial neural network (ANN) algorithms have been trained to discriminate patients with JIA from healthy controls using deep immunophenotyping characteristics [55]. Up to 42 immunological parameters (e.g., iNKT cells, *γδ* T cells, CD4+ T cells) that could act as a disease signature were collected from 128 patients. After training the models, the authors found that the discrimination ability of RF was superior to the rest of the models, achieving a 0.89 AUC value, being the number of iNKT cells the key contributing feature to discriminate JIA patients from healthy controls. With this study, researchers showed that ML can take advantage of immunophenotyping to identify immune signatures that correlate with JIA disease.

The existing diagnostic techniques may not be sufficiently sensitive to detect the subtle changes that occur during the early stages of OA, when it is still an asymptomatic condition. In [56], authors used data from the Osteoarthritis Initiative (OAI) to identify serum antibodies that could predict radiographic knee OA in asymptomatic individuals, measured with the KL scale, that will develop the disease during a follow-up period of 96 months. Clinical variables (e.g., age, sex, BMI, WOMAC) including and excluding a molecular biomarker (i.e., MAT2*β*-AAb) were used to build stepwise multivariable logistic regression models. An AUC value of 0.76 was achieved in the replication-independent cohort.

#### 3.2.3. Image

Certain anatomical structures can act as a disease’s sign or symptom and, therefore, be used for diagnosing pathologies. For instance, halo sign detection in giant cell arteritis (GCA), is an example of such structures. In a recently published study [57], authors used 1,311 colour doppler US images from 137 patients to train a U-Net, a CNN for semantic segmentation, able to detect halo sign based on a pixel score metric, achieving a 0.83 AUC in the test set. For enriching the training set, data augmentation was performed.

On the other hand, sacroiliac joint erosion is an early symptom of AS. A study compared the performance of ML and DL classifiers in detecting erosion in a set of 681 sacroiliac joint computed tomography (CT) images from 53 patients. For the former, ML, the authors extracted texture features and built a set of predictors with grey-level co-occurrence matrices (e.g., energy, contrast, correlation, homogeneity, and dissimilarity) and local binary patterns (LBP) (e.g., count of LBP with a concrete pattern). Then, different classifiers (e.g., KNN, RF) were trained and compared. For the latter, DL, they used and modified a pre-trained model, InceptionV3, via transfer learning. The authors concluded that DL outperformed, 0.97 AUC, the ML algorithms and a radiologist with 9 years of experience in terms of sensitivity and specificity [58].

Due to its superior textural contrast resolution, MRI is widely acknowledged as the most sensitive technique for the early detection of inflammatory sacroiliitis. On this basis, authors in [59], calculated features from the histogram of MRI images (e.g., kurtosis, skewness, maximum pixel value) and trained three ML methods, including SVM, KNN and multilayer perceptron (MLP), for computer-aided classification of active inflammatory sacroiliitis to assist in the spondyloarthritis classification. Some of the patients developed axSpA and some others OA, fibromyalgia, gout, or psychiatric disorders. The best classifier, KNN, achieved an accuracy value of 0.80.

Combining variables from different domains to improve the performance of DL methods was a concern of some researchers that tried to predict early hip osteoporosis to avoid fractures. Following an ensemble method, investigators from Japan [60] achieved a 0.93 AUC in the diagnosis of osteoporosis of the proximal femoral, after training five well-known pre-trained networks (i.e., ResNet18, ResNet34, GoogleNet, EfficientNet b3, EfficientNet b4) with a set of 1,131 hip dual-energy X-ray absorptiometry images plus four clinical covariates: age, sex, BMI, and history of hip fracture. The addition of clinical variables improved the AUC and the accuracy by 2.58% and 4.78% on average.

Identifying patients who are at risk of bone fractures remains a significant challenge in the field of osteoporosis. The researchers in [61] fitted 15 classifiers (i.e., linear models, KNN, SVM, tree-based models and ensemble models) to a cohort of 92 osteoporosis patients to predict fragility fractures from MRI data. The authors extracted different features from the images: mechanical (e.g., elastic and shear modulus), physical (e.g., mean bone volume), topological, and statistical. Other features, such as BMI or age, were also considered and PCA was used for reducing the original 55 features into different datasets comprising 5, 30 and 55 variables. Finally, up to 6 different datasets were built. The average F1 score obtained was 0.63 for the all-features dataset.

Other studies with a strong computer vision involvement have been proposed. For example, central sarcopenia detection was the main goal in [62]. In this study, CT scans of 102 patients were segmented and analysed using a pre-trained CNN, U-Net. Five muscles (e.g., psoas, quadratus), and five lumbar spine levels (i.e., L1-L5), were included and the performance of the system was evaluated using Dice similarity coefficients. Dice values between 0.80 and 0.93 and between 0.82 and 0.93 were obtained in the test set for the muscles and the spine levels, respectively. Although the authors of the study highlighted the potential use of this system in the detection of central sarcopenia, no predictive models were developed from the results shown here. Hence, this article could be categorised into another topic depending on the intended use of the segmentation system.

In certain pathological conditions, radiological interpretation by an operator can be subjective and potentially influenced by biases. Under this premise, novelty detection models to screen for myopathies and find rare presentations of myopathic disease (i.e., myositis), have been presented in [63]. In this study, the authors used 3,586 US images from 35 controls and 54 patients with myositis, annotated the dataset (i.e., Myositis3K dataset) and tried different novelty detection algorithms based on discriminative DL networks and generative methods. Regarding the discriminative method, authors proposed the following pipeline: the use of deep embeddings to obtain a new representation of the US images, the use of PCA and t-SNE for dimensionality reduction, and the computation of a novelty score. Regarding the generative methods, they used generative adversarial networks (GANs). The best-performing approach, the discriminative method, resulted in a 0.72 AUC value. This approach used SVM to calculate the novelty score.

Eventually, new imaging techniques have been proposed that utilize the microvascular changes at the finger nailfold, which can aid in the diagnosis of SS. The authors in [64] used optoacoustic imaging techniques (i.e., raster-scanning optoacoustic mesoscopy) to obtain images from 23 SS patients and 19 healthy controls. Then, researchers used a pre-trained CNN to classify the images, obtaining an AUC value of 0.90 and demonstrating the capabilities of optoacoustic imaging for SS diagnosis.

### 3.3. Disease classification

This section describes studies that attempt to classify patients with different rheumatology conditions, regardless of trying to also compare patients with healthy controls at the same time (e.g., pSS/SLE, pSS/controls, SLE/controls).

#### 3.3.1. Clinical and demographic

Establishing the correct diagnosis at an early disease stage can condition the patient’s future health outcomes. In some cases, the initial course of a disease may not be clear, as symptoms can be shared among competitive diseases, which clinical management is totally different. In this regard, Yu SC et al. [65] built different classification models (i.e., SVM with different kernels, RF) to distinguish lupus lymphadenitis (n = 19) from Kikuchi disease (n = 81) using 32 clinicopathological characteristics: 6 clinical features (e.g., age, sex, biopsy site/method, the extent of lymphadenopathy), 10 histological features (e.g., area involved, necrosis), and 16 C4d immunohistochemistry staining results (e.g., endothelial staining in necrotic area, staining at adipocytes) in a case series of one hundred patients. Models were externally validated, achieving high sensitivity, 1, and specificity, 0.96. However, the validation cohort was small and extremely unbalanced, with only two cases of lupus lymphadenitis. This study highlighted the relevance of AI techniques in the study of rare diseases and their manifestations.

A model to produce SLE risk probabilities (i.e., unlikely, possible, likely, and definite) was presented in [66]. In this study, authors combined clinical and serological features from three SLE classification criteria (i.e., ACR 1997, SLICC 2012 and EULAR/ACR 2019) with non-criteria features (e.g., fatigue, lymphadenopathy, sicca) to calculate SLE risk probabilities. 401 SLE patients and 401 disease controls, including RA, undifferentiated connective tissue disease, pSS, scleroderma and vasculitis patients, among others, were included in the discovery cohort. After performing a correlation analysis, researchers created 20 panels of features, and each panel was used to train two algorithms, Lasso-logistic regression (LR) and RF. The best-performing model, Lasso-LR, achieved a 0.98 AUC in the validation cohort and included 14 features. Eventually, to facilitate the adoption of such a model into clinical practice, the authors used k-means clustering to detect unbiased risk probabilities partitions and to build a simple scoring system.

#### 3.3.2. Molecular biomarkers

Based on the findings of a previous study in which the authors found several plasma microRNAs (miRNAs) altered among patients with RA, researchers in [67] developed a model capable to classify RA patients from controls, AUC 0.71, SLE patients from controls, AUC 0.80, and RA from SLE patients, AUC 0.63, using panels of miRNAs biomarkers. For this purpose, RF and Lasso were employed. Whereas the former was used to capture a list of candidate miRNAs, the latter was used to select a final miRNAs panel that maximised discrimination between diseases. Although the panel differentiated RA and SLE patients from controls, it was unable to differentiate properly between patients with RA and SLE.

The T-cell receptor diversity and the potential use of this information to classify diseases was the initial hypothesis of a Chinese group of researchers, Liu et al. [68], that conducted an investigation for classifying RA (n = 206), SLE (n = 877) and healthy controls (n = 439). RF models to discriminate RA patients from controls, SLE patients from controls and RA from SLE patients were built, obtaining in all cases AUC values above 0.99.

The clinical courses of certain pathologies, such as RA, may exhibit unusual manifestation patterns and laboratory values (e.g., seronegative RA), therefore their diagnosis can be confused with other pathologies such as psoriatic arthritis (PsA). The authors of [69], used RF, NB, multivariate logistic regression and hierarchical clustering to identify patients suffering from seronegative RA (n = 49) and PsA (n = 73). They achieved an AUC value of 0.71 in the validation cohort, using demographic characteristics and serological concentrations of amino acids as predictors (i.e., age, gender, lipid ratios L6/L1, L5/L1, L2/L1, alanine, succinate and creatine phosphate), calculated with the aid of H nuclear magnetic resonance-based metabolomic and lipidomic analysis of serum.

Other clinically and immunologically related diseases are SLE and primary Sjögren’s Syndrome (pSS). As these diseases have common and specific manifestations, some researchers have investigated DNA methylation changes to detect disease-specific alterations. For instance, in [70], DNA methylation data were used to classify patients with SLE (n = 347), pSS (n = 100) and controls (n = 400), employing RF. Four different models of disease status were developed: SLE/control, pSS/control, pSS/SLE and positive Ro/SSA and La/SSB pSS patients and controls. In these models, methylation *β*-values were used to predict disease status. For that purpose, a feature selection of CpG sites based on linear regression was done beforehand. The authors achieved an AUC value between 0.83 (pSS/SLE) and 0.96 (pSS/control). This is, disease classification (i.e., pSS/SLE) was harder for the model than disease prediction (i.e., pSS and healthy controls).

Finally, the authors in [71] theorised that bacterial nucleic acids that may exist in synovial tissue and fluid could be involved in arthritis development, therefore, they studied bacterial nucleic acids in synovial fluids, with 16s rRNA gene amplicon sequencing, from 58 OA and 125 RA patients. The authors built SVM and RF classification models to discriminate between OA and RA patients, using operational taxonomic unit markers. They obtained a mean AUC value of 0.79.

#### 3.3.3. Image

In early disease stages, some inflammatory markers commonly used to distinguish one disease from another are not altered. Hence, its reliability may be reduced. Being aware of the early diagnosis difficulty in hand arthritis, researchers in [72] used DNN and transfer learning, InceptionV3, for classifying photographs of hands into OA, RA, and PsA. Firstly, they developed a DL model to estimate the probability that an image had one of those conditions. Then, they combined this information with validated questionnaires and a single examination technique to determine the most likely diagnosis in a patient presenting with hand arthritis. The number of participants was 280, and the algorithms employed were SVM, RF and LR. The accuracy oscillated between 0.78 and 0.97 when classifying OA and inflammatory arthritis, and between 0.90 and 0.95 when classifying RA and PsA.

The applicability of knee thermograms as a screening tool in the subclinical stages of RA diagnosis, and as a way to distinguish types of arthritis has been postulated by a group of Indian researchers [73]. Firstly, they employed different algorithms for inflamed region segmentation (e.g., k-means, Otsu). Then, they computed seven features from the region of interest (e.g., area, Euler number, perimeter), and trained four different linear SVM classification models, depending on the segmentation algorithms initially employed, to distinguish knee arthritis conditions from normal knees or pathological knees with unknown reason. The authors obtained an accuracy of 0.91 on this task. Secondly, they developed additional SVM models to classify RA knees from other types of arthritis. Three categories of features were extracted for that aim, texture features (n = 55), frequency level features (n = 2), and shape features (n = 7). Finally, a feature selection analysis was performed considering different approaches (i.e., recursive feature elimination and RELIEF). AUC values ranging from 0.67 to 0.72 were obtained.

### 3.4. Patient stratification and disease subtype identification

This section describes studies that attempt to stratify patients into meaningful subgroups that share similar characteristics (e.g., histological and/or molecular features, diseases), and that are different from other subsets of patients. Studies that attempt to classify patients into different activity groups are also included in this category. Although the main objective of these studies is the identification of subgroups of patients, usually employing clustering techniques, studies that also incorporate prediction models for classifying the patients into the subsets previously identified are also considered in this category.

#### 3.4.1. Clinical and demographic

Some researchers have tried to refine the spectrum of specific autoantibodies that can appear in multiple diseases with distinct phenotypes such as anti-Ku antibodies. With that aim, Spielmann et al. [74] tried to identify subgroups of anti-Ku-positive patients using hierarchical clustering methods. Three clusters of connective tissue disease patients (n = 42) with anti-Ku antibodies and with similar clinical and biological features and prognosis on a set of 28 clinico-biological features were found, 3 demographic features (i.e., sex, age, geographical origin), 17 features related to organ involvement (e.g., arthralgia, lupus rash), and 8 autoantibodies (e.g., rheumatoid factor, ACPA). Multiple correspondence analysis was used for dimensionality reduction. From a data science perspective, the relevance of this study lies in the discussion generated by the scientific community, on the suitability of the techniques employed, through letters to the publisher [75, 76, 77, 78].

Survival analyses can be conducted for different subgroups of patients after being identified by clustering algorithms, for diseases with increased mortality risk due to life-threatening complications. A hierarchical clustering approach was also conducted by Ogata et al. [79] to identify and clarify the characteristics of the subgroup of antiphospholipid syndrome (APS) patients with the poorest prognosis (n = 168) at the time of diagnosis. Up to 14 serological and clinical variables, including age at APS onset, SLE, hypertension, history of arterial thrombosis and IgG/IgM aCL were used as input to the algorithm. Three different clusters were identified after analysing the dendrogram visually. Although the clustering groups identified in this study were different compared to previously published results, which only considered serological data, the authors highlighted the existence of a cluster that accumulates cardiovascular risk events and arterial thrombosis events, with significantly higher mortality compared to the other clusters.

This unsupervised algorithm, hierarchical clustering, has also been applied to a dataset of 70 adult-onset Still’s disease (AOSD) inpatients [80]. Three distinct fever patterns were characterised after computing the ideal number of clusters, *k*, with the Krzanowski and Lai index. In view of the results obtained, after applying logistic regression to compare the prognosis of AOSD between the three groups, the authors concluded that a higher temperature at the time of diagnosis was associated with a higher risk of AOSD-related mortality. As input features, the authors considered both clinical and demographic features (e.g., sex, age, diabetes mellitus, clinical course, fever).

Another research group employed unsupervised ANN algorithms and graph-based approaches (i.e., semantic connectivity maps) to explore hidden trends and non-linear associations among clinical (e.g., disease duration, lymphadenopathy) and serological pSS features (e.g., rheumatoid factor, C4 level), and to predict lymphoma [81]. Finally, three subsets of patients with different characteristics (i.e., predominant glandular manifestations, anti-Ro/SSA positive patients and patients with vasculitis manifestations) were identified. This research group was also responsible for studying the increased prevalence of cardiovascular events in pSS patients using, again, unsupervised ANN algorithms (i.e., Auto-CM) and agglomerative hierarchical clustering [82]. With the first approach, authors found two different patterns of distributions in cardiovascular events risk factors.

Attempts have been made to identify patients predisposed to the development of lymphomas associated with pSS [83]. The dataset studied consisted of 449 pSS patients (76 of them with lymphoma) and 90 features including demographic (e.g., year at diagnosis, gender), and laboratory measures (e.g., C3, C4, haemoglobin). Two algorithms were trained, RF and extreme gradient boosting (XGBoost); and the prediction performance was assessed by cross-validation. While RF showed an AUC value of 0.83, XGBoost achieved a score of 0.88. C4 levels, the rheumatoid factor, and the focus score at first biopsy were the predictors with the highest relative importance.

Non-negative matrix factorisation (NMF), an unsupervised pattern recognition algorithm, was used to identify joint involvement patterns, understood as frequently co-involved joints, capable of predicting clinical phenotypes and disease trajectories in 640 JIA patients [84]. In this study, the authors raised the idea that using these joint patterns could be helpful to better classify JIA patients. Seven distinct joint patterns (i.e., pelvic girdle, fingers, wrists, toes, ankles, knees, indistinct), reproducible through external data set validation, with 119 patients, were identified.

#### 3.4.2. Molecular biomarkers

Researchers sought to identify subgroups of paediatric SLE patients to tailor the treatment plan based on the underlying pathogenesis of the disease [85]. They found two differentiated groups of paediatric SLE patients (n = 31), depending on the predominant component of the disease: autoimmune or autoinflammatory. K-means clustering was used for this purpose considering type I interferon (IFN) score, SLEDAI-2K and mean complement levels (i.e., C3 and C4 normalised values) variables. Finally, PCA was used to visualise the clustering results.

Some studies have compared the performance of classical serological biomarkers with gene signatures. In this regard, clusters enriched in active SLE, quantified using SLEDAI, were identified using hierarchical clustering in 140 SLE patients [86]. In this study, blood, serum, and clinical data were obtained together with three gene signatures, type I IFN, polymorphonuclear neutrophil and plasmablast. A bootstrap forest model was used to predict SLE activity (i.e., clinical SLEDAI ≥2) and to identify potential predictors related to the disease activity. The results were validated by performing multivariable logistic analyses. Among the variables with a higher predictive contribution, a composite score created by adding IFN-I genes to polymorphonuclear neutrophil genes showed the highest predictive power. Other classical serological variables considered were C4, C3 or glucocorticoids (GCs) dose.

On their behalf, the authors of [87] investigated how they could obtain meaningful clusters applying NMF in a cohort of 173 patients with SS, using skin gene expression profiles. Cophenetic coefficients and silhouette score metrics were used to determine the number of clusters, *k*. The authors achieved satisfactorily a four-cluster separation based on different activation levels of SS-relevant signalling pathways (e.g., Notch, TGF*β*). t-SNE was used to visualise the different groups.

Clinical variables and laboratory biomarkers from a cohort of 150 children with JIA were used to identify clusters with the help of GMM [88]. Visits at baseline and six months later were considered. With a feature selection process based on a variable contribution threshold, 191 features were embedded into three principal components which represented 35% (baseline visit) and 40% of variance (six months after the first visit) and retained 37 and 38 variables, respectively. Using the Bayesian information criterion, the authors found three and five clusters. Later, the researchers tried to compare the results of the clusters with currently defined JIA categories using circular plots. In light of the results, the authors concluded that the clusters did not match JIA categories, suggesting that the pathobiological processes are shared between JIA categories and fluctuate during the course of the disease.

Under the hypothesis that genetic and polygenic risk scores can accurately predict cases from controls with high specificity, and considering the recent advances in array technologies and their affordability, another case-control JIA classification research article was published [89]. This time, the authors pursued to build genomic risk scores (GRS) for diagnosis using single-nucleotide polymorphism (SNP)s and PCA. Lasso and elastic net were used as penalised regression models with the first 10 genetic principal components and sex as input features. The training cohort consisted of 2,324 cases and 5,181 controls. The external validation was performed with two cohorts with a total of 921 cases and 3,532 controls. After performing the analysis between cases, without discriminating JIA subtypes, and controls; subanalyses to consider each JIA subtype, according to ILAR, and all controls were also performed. A key idea of this study was the design of JIA subtype-specific GRS for the seven mutually exclusive categories of JIA.

Poppenberg et al. [90] considered different supervised ML algorithms (i.e., KNN, RF and SVM) to assess the association between JIA disease activity and transcriptomes from peripheral blood mononuclear cells. In this research, the authors defined active disease according to the presence of physical signs of synovitis in at least one joint. The identification of predictors (i.e., transcripts with the greatest predictive power) was performed with Lasso. Finally, the 35 genes identified (e.g., ACAP3, ARL2BP) were used as the input of four predictive models (i.e., KNN, RF, SVM). RF outperformed the rest of the models in the testing cohort, achieving a 0.94 AUC value.

Consensus clustering and k-means were the approaches chosen by the researchers to reveal three RA synovial gene expression subtypes using the top 500 most variable genes expressed in 45 RNA-seq samples (i.e., 39 RA and 6 OA synovium) [91]. The ideal number of clusters, *k* = 3, was visually confirmed according to likelihood scores and PCA was used to validate the clustering. After that, the authors used histology scores as modelling features in a standard SVM to predict the three RNA-seq subtypes. The AUC value of such models ranged from 0.59 to 0.88.

Extra-articular manifestations presented in some rheumatic diseases can contribute significantly to morbidity and mortality. With that in mind, PCA and Lasso techniques were used for patient stratification based on the combination of distinct serum protein biomarkers in RA-interstitial lung disease (ILD) and RA-no ILD patients, idiopathic pulmonary fibrosis patients and healthy controls [92]. The serum levels of 45 proteins consisting of cytokines, chemokines, growth factors, and remodelling proteins were measured using multiplex ELISA-based assessment, and the authors identified seven biomarker signatures (e.g., MMP-1, MMP-2, MMP-7, MMP-9, interleukin (IL)-1, receptor antagonist) that effectively differentiated both diseases, RA-ILD / RA-no ILD, achieving an AUC value of 0.93.

#### 3.4.3. Image

Clustering methods have been employed to determine to what extent disease subgroups, which have been speculated to be part of the same pathological condition, such as some forms of large vessel vasculitis, are anatomically distinct disease entities. For instance, Gribbons et al. [93], included 1,068 patients in a study to identify groups of patients with similar patterns of Takayasu’s arteritis (TAK) and GCA large vessel vasculitis. By defining 11 arterial territories (e.g., carotid, subclavian, axillary, renal, mesenteric, and aorta) and combining catheter-based, magnetic resonance, computed tomographic angiography, ultrasonography, and fluorodeoxyglucose positron emission tomography images; patients were clustered based on disease within those arterial territories. Silhouette score and gap-statistic methods were used for determining the optimal number of clusters, *k*, and k-means was chosen as the clustering algorithm. Six unique clusters of patients with distinct patterns of arterial involvement were found. The key aspect of this study was the sample size for such rare diseases, which facilitated the use of clustering techniques. The results of this research were proposed by the authors to be considered in future classification criteria for large vessel vasculitis.

This same research group, [94], in a previous study, also included a DT model to predict k-means cluster assignment based on 13 arterial territories in these two diseases, TAK and GCA, achieving a 0.87 accuracy in the replication cohort. Three clusters were identified with significant differences in the prevalence of arterial involvement. In both studies, independent cohorts were used for validation.

Doppler ultrasound images have proven to be helpful when training CNN architectures. Scientists have recently demonstrated its viability to automate the classification of disease activity into four degrees, using the EULAR-OMERACT Synovitis Scoring (EOSS), performing similarly to a human expert [95, 96]. In [95], authors proposed two pre-trained CNN, InceptionV3 and VGG-16, for automatically scoring the disease activity of RA patients, using images from the wrist and the hand of 40 patients with early or longstanding disease, achieving an accuracy of 0.75.

A year later, the same group of researchers [96] used a dataset of 1,678 ultrasound (US) images, and trained six CNN for binary classification, in a cascaded architecture; three CNN trained from the scratch and three trained on features extracted from InceptionV3, achieving a four-class accuracy of 0.84, and beating their previous results. In this cascaded architecture, the output of each CNN was a class prediction of the EOSS system. The same analysis was repeated using data augmentation for enriching the training set.

Moreover, smartphone pictures have been proposed to train SVM models capable of classifying hand arthritis photographs, based on the deformation of hand fingers, into three stages (i.e., early, moderate, late); and between healthy and unhealthy [97]. The accuracy obtained ranged between 0.77 for three-stage classification, and 0.97 for binary classification.

US images have also been used to discriminate between low and high-grade synovitis in inflammatory arthritis patients. In [98], the authors explored the ability of CNN and transfer learning using a pre-trained CNN, ResNet34, for discriminating between both synovitis grades, in 150 photomicrographs of 12 patients, achieving perfect accuracy.

### 3.5. Disease progression and disease activity

This section describes studies that aim to characterise the natural history of a disease or that aim to forecast disease prognosis, such as flares, or mortality. Patients included in these studies must be diagnosed in advance.

#### 3.5.1. Clinical and demographic

New approaches for characterising the natural history of diseases without the need for manual review are required, especially in diseases where symptoms are usually not collected in detail or in a structured way. Topic modelling has been applied to characterise the temporal evolution of ANCA-associated vasculitis (AAV) patients in [6]. With a follow-up of seven years, over 113,000 clinical notes from 660 patients were processed. Temporal trends, before and after the treatment initiation date for a diagnosis of AAV, were modelled with LDA, finding 90 different topics that included diagnosis (e.g., granulomatosis with polyangiitis), treatments (e.g., AAV specific-treatment), and comorbidities and complications of AAV (e.g., glomerulonephritis, infections). The authors showed the suitability of this unsupervised method to provide unique information on the clinical course of a disease that is not totally captured in the structured medical record data. As the researchers highlighted, identifying the topics in a clinical note is of special relevance in multiorgan diseases, where structured data fields are unlikely to reflect the full extent of signs, symptoms, comorbidities, and complications.

Calculating disease activity indices is not a usual practice in real-life settings due to its time-consuming nature. In [99], investigators developed a predictive model for SLE disease activity based on routinely available demographic, clinical, and laboratory data. With this model, they tried to overcome the current limitations of using a complex composite index such as SLEDAI-2K to measure disease activity. The authors used 16 clinical laboratory results (e.g., creatinine, C-reactive protein (CRP), C3, C4) along with sex, age and ethnicity demographic variables, and a multinomial logistic regression approach that was compared to the performance of a NB model. To select the best-performing model from the space search (i.e., 2^n^, where *n* = 16), an 0.82 AUC threshold was fixed. Models with up to 8 variables that did not include anti-dsDNA assay results were selected. Finally, researchers found that the multinomial approach overcomes the performance of NB, with 0.83 and 0.66 AUC values respectively. In light of the results, the researchers concluded that building a model to accurately calculate disease activity and that relies on routinely clinical parameters is feasible.

Predictive models to forecast disease activity have been proposed as a way to tailor current therapeutic approaches to prevent disease progression. One remarkable study on the application of DL techniques to RA patients was carried out by Norgeot et al. [15]. In their research, the authors aimed to forecast RA disease activity, measured with Clinical Disease Activity Index (CDAI), in future clinic visits using 45 structured variables from the EHR, including disease-modifying antirheumatic drugs (DMARDs), corticosteroids, CDAI score, erythrocyte sedimentation rate (ESR), CRP, anti-CCP, rheumatoid factor and demographic variables. In doing this, the authors imitated time series forecasting studies, creating sliding time windows of a fixed interval to model the longitudinal nature of the study. Permutation importance scores, to measure the contribution of each independent variable, were calculated. Convolutional and recurrent neural networks and dense and time-distributed layers were tested. The best model achieved a 0.91 AUC value in the test set.

Mortality prediction models can be useful for the management of autoimmune diseases. For instance, a RA study conducted by Spanish researchers, focused on developing a RA mortality predictive model, trained (n = 1,461) and validated (n = 280) using random survival forests, a supervised predictive algorithm [13]. In this study, nine demographic and clinical-related variables, such as age at RA diagnosis, gender or presence of rheumatoid factor; and collected two years after diagnosis, were included in the final model after a variable importance analysis according to their predictive ability. The authors identified three different mortality risk groups (i.e., low, intermediate, and high) using the predicted ensemble mortality. The prediction error in the validation cohort was 0.233.

Under the hypothesis that RA and axSpA flares are associated with physical activity, a French research group took advantage of wearable activity trackers and NB algorithm to study potential associations between flares and steps [8]. With 155 patients, 82 RA and 73 axSpA, and 1,339 weeks evaluated, the authors concluded that patient-reported flares were strongly linked to physical activity. To reach this result, different wearable data time levels of aggregation were considered, the binary variable flare/no flare was used as the dependent variable, and the performance of the models was evaluated using patient-reported flares as the gold standard, assessed every week.

Flares are not uncommon for people with chronic conditions. These events, characterised by a worsening of the disease, may require treatment changes, a longer hospitalisation stay, and increased healthcare costs. Inpatient gout flares predictors have been recently assessed in [100]. The aim of this study was to develop a prediction model for inpatient gout flare. Up to 52 potential variables from five different domains were evaluated, including demographics (e.g., age, sex, ethnicity), comorbidities (e.g., acute renal failure, arthritis, osteoporosis), admission (e.g., primary admission diagnosis), gout history (e.g., tophus) and laboratory data (e.g., pre-admission urate >0.36 mmol/L). The researchers followed three different approaches: a clinical knowledge-driven model (LR), a statistics-driven model (Lasso) and a DT model. Model validation was performed with bootstrapping. Based on the C-statistic value, C = 0.82, the first model was selected as the best-performing with only nine predictors. Almost half, 4 out of 9, of the selected variables were chosen by the three different models such as pre-admission urate>0.36 mmol/L or urate-lowering therapy adjustment. This study revealed the importance of building an intuitive model for clinicians, and feasible to implement in a routine hospital setting.

#### 3.5.2. Molecular biomarkers

The Osteoarthritis Initiative (OAI) database served as an independent cohort, n = 204, to validate SVM models trained to predict radiographic progression using peripheral blood leukocyte inflammatory gene expression (e.g., IL-1*β*, TNF*α*, and cyclooxygenase-2), clinical and demographic (e.g., age, sex, BMI), and imaging-related variables (e.g., osteophyte scoring) [101]. The models were trained with different combinations of variables, with the best model obtaining a 0.68 AUC value. A highlight of this study was the inclusion of biomarkers from different domains (i.e., clinical, image, gene) to maximise the performance of the models.

#### 3.5.3. Image

Relevant quantitative measures of joint degeneration and disease progression may require a thorough segmentation preprocessing step. Therefore, some researchers have focused on this first step as the main goal of their studies. In the case of knee OA, different authors have applied multiple DL topologies such as CNN [102, 103], conditional GANs [104] and holistically nested networks [105] for the segmentation of knee joint tissues, including, among others, femoral cartilage, tibial cartilage, patella, patellar cartilage, meniscus, quadriceps and patellar tendon, or infrapatellar fat pad. Although some of these articles did not implement a predictive model and just presented a segmentation system, we wanted to point out the relevance of such pre-processing step for conducting further research.

### 3.6. Treatment response

This section describes studies that attempt to study the response to a drug, regardless of how this response is measured (e.g., disease activity/progression, patient’s concern and perception, safety, efficacy, and appearance of adverse events). The central idea is that the patient has had to be exposed to a drug and a response to this exposure must have been registered.

#### 3.6.1. Clinical and demographic

In [106], the authors fed an ANN, among other ML algorithms like XGBoost, or RF, with clinical (i.e., age, sex, height, weight) and laboratory data (e.g., HLA-B27, white blood cell count, haemoglobin) from almost 600 AS patients to predict early-TNF responders, obtaining a 0.783 AUC value with the ANN model. In addition, a feature importance analysis based on gradient descent was helpful to determine that CRP and ESR were the most significant baseline characteristics for predicting early-TNF responders.

NLP techniques were used to identify arthralgia in clinical notes from inflammatory bowel disease patients, as a preliminary step to compare two different treatments (i.e., vedolizumab and TNFi), one of them apparently related to an increased risk of arthralgia due to adverse events [107]. More precisely, the positive mention of 37 unique concepts was recorded. Afterwards, an inverse probability of treatment weighting approach was followed, including eight variables such as age, sex, medication or follow-up time; and Cox proportional hazard regression models were computed. The study’s authors observed no notable rise in the likelihood of arthralgia linked to the use of vedolizumab when compared to TNFi. Thanks to narrative notes and NLP, authors were able to characterise the role of a drug (i.e., vedolizumab) in the appearance of a potential adverse effect (i.e., arthralgia), in a scenario in which coding was suboptimal.

Web scraping techniques have been used to extract data from social media networks in a variety of contexts. Treato, a deprecated data analytics service, was used in some of them. This service incorporated NLP processing pipelines including medical ontology mapping, classifiers, and sentiment analysis among others. For example, in an attempt to evaluate the suitability of social media as a data source for drug safety, some authors studied patient-reported herpes zoster events associated with arthritis medication (i.e., tofacitinib vs. other therapies) [108]. For this purpose, the authors used Treato to analyse and classify more than 785,000 posts mentioning inflammatory arthritis with a PPV of 0.91.

Another social media data web scrapping research that used Treato and LDA for topic modelling, has been carried out by Dzubur et al. [109], to examine AS patients’ knowledge, attitudes, and beliefs regarding biologic therapies. 27,000 posts from more than 600 social media sites were studied. The investigators found 112 topics, 67 of them focused on discussions related to AS biological therapy, such as the side effects, the biological attributes (e.g., dose and frequency) or the patients’ concerns (e.g., cancer risk, reproductive concerns).

The perceptions of RA patients to 13 DMARDs were assessed in [110] using Treato. This time, the NLP task aimed to identify medical concepts and extract patients’ self-reported descriptions of their experiences with various health conditions and medications, to eventually conduct sentiment analyses. The authors found that the ratio of patients with positive sentiment to biological and targeted synthetic DMARDs was higher than the ratio of patients with positive sentiment to conventional synthetic DMARDs. In addition, they showed that efficacy and side effects were the most frequently discussed topics.

Researchers have studied the response to methotrexate monotherapy [111] and to TNFi (i.e., etanercept) [112] in JIA patients using Disease Activity Score (DAS)44/ESR-3 indices. Regarding the former, [111], treatment response models before and after administration within three months were built in a cohort of 362 patients, using clinical (e.g., JIA subtype), demographic (e.g., gender, weight) and laboratory variables (e.g., ESR, IgG). The algorithms proven were XGBoost, SVM, LR and RF. A median importance ranking with ensemble methods was also computed. A set of ten predictors before (e.g., CRP, fibrinogen, active partial thrombin time) and six predictors after treatment administration (e.g., CD3+CD4+, CD3+CD8+) were chosen by the XGBoost algorithm. The performance of the models in both scenarios, before administration and before and after administration, was 0.97 and 0.99 AUC respectively.

Regarding the latter, [112], treatment response models before administration were built in a cohort of 87 patients. The algorithms proven were XGBoost, gradient-boosting decision trees, extremely randomized trees, LR and RF. From 47 pre-administration clinical and laboratory variables, four features that maximised the predictive performance were identified by XGBoost: tender joint count (TJC), time interval, lymphocyte percentage and weight. This model achieved a 0.79 AUC value.

The cardiovascular side effects of analgesics in 4,350 patients, retrieved from the OAI dataset, were modelled by an XGBoost prediction model along with a risk feature identification [113]. Of 300 demographics (e.g., age), anthropometry (e.g., BMI, mental summary scale), comorbidity (e.g., asthma), blood measures (e.g., diastolic), and physical activity (e.g., walk pace, heavy housework) features, the authors found and described the 20 most informative ones (e.g., age, radial pulse, asthma, weight). The model achieved a 0.92 AUC value.

The response to treatment has also been evaluated in PsA patients (n = 2,148), from four phase 3 trials (i.e., FUTURE 2 to 5), in [114]. In this original article, the efficacy of the starting dose of secukinumab, an IL-17A inhibitor, was evaluated thanks to the Bayesian elastic net ML algorithm. More specifically, the study sought to investigate whether there were specific baseline clinical characteristics that could predict which patients could gain additional benefit from the secukinumab 300 mg dose. With a cohort of 2,148 patients and 275 predictors, different efficacy endpoints (e.g., ACR20/50, PASI 75/90, PASDAD, HAQ-DI) were analysed at week 16. Forty clinical variables were used as covariates such as smoking status, presence of enthesitis, presence of dactylitis, or methotrexate use. Although there was no single predictor with enough discriminatory power, the authors found that there were common covariates for different endpoints, such as the presence of enthesitis at baseline. Furthermore, the authors also identified subpopulation groups that could benefit from the 300 mg dose over the 150 mg dose, such as patients treated without concomitant methotrexate or patients with psoriasis. The AUC scores ranged from 0.75 to 0.81 for the different endpoints.

Serious infections in RA patients under IL-6 inhibitor treatment have been studied in [115]. Researchers from Japan extracted data from a post-marketing adverse events-reporting database using text mining approaches to identify signs and symptoms before the development of serious infection (i.e., defined by the authors as those infections in which the patient attended the hospital). Once the signs and symptoms were extracted from clinical narratives 28 days before serious infection, a codification with MedDRA Preferred Terms, and a review to determine if they were already generally known as signs or symptoms of infection was done. As a result, the authors showed that more than 60% of patients with a confirmed date of serious infection diagnosis had signs or symptoms within 28 days before that diagnosis. The most common symptoms were pyrexia, pain, cough, and swelling.

Response to intravenous immunoglobulin therapy in patients with Kawasaki disease has been studied in [116]. Researchers applied seven ML algorithms, including regularised linear regression, tree-based and boosting algorithms; and obtained a 0.74 AUC with GBM. For that purpose, they combined 82 clinical and demographic (e.g., sex, age, weight), laboratory (e.g., blood urea nitrogen, white blood cell count) and sonography image measurements (e.g., left main coronary artery diameter). The importance of the features was evaluated with SHapley Additive exPlanations (SHAP).

Eventually, reinforcement learning and sequential decision-making algorithms have been implemented to promote physical activity in patients with chronic back pain (CBP) [117] in a smartphone application. In a rigorous manner, this research article does not study the response to a drug or procedure. However, we consider it particularly relevant due to the approach employed (i.e., reinforcement learning) and the non-autoimmune nature of the disease (i.e., back pain).

#### 3.6.2. Molecular biomarkers

A non-negligible number of treatment response studies have been conducted to assess the efficacy of new therapeutic lines in the RA population. The response to tumor necrosis factor inhibitor (TNFi) has been investigated in multiple studies. For example, in [118], penalised regression models were used to estimate changes in ESR and in the swollen joint count, two DAS28 composites, between 3 and 6 months after treatment initiation. Clinical and genotypic scores covariates (i.e., first 10 genetic principal components) were used to build the predictive models. Nonetheless, the authors were unable to find strong predictors of TNFi response among alleles linked to the development of RA.

In another study [119], authors predicted changes in disease activity scores 24 months after baseline assessment (i.e., ΔDAS28), and identified non-responders to anti-TNF treatments using different ML techniques (e.g., SVM, Ridge, RF, LR, and Gaussian process regression). Demographic (e.g., age, sex), clinical (e.g., baseline DAS, methotrexate dose), and genetic features (e.g., SNP) were included as predictors, although the last ones did not improve the prediction accuracy. The AUC value of the best model in the independent cohort was 0.62.

On its behalf, the authors of [120] based their research on determining how the transcriptomic and epigenetic profiles of immune cell types and whole peripheral blood mononuclear cells could help to predict the response to two different TNFi (i.e., etanercept and adalimumab), prior to treatment initiation, using RF models. For that purpose, authors built two predictive models, one for each treatment and considered 461 differentially expressed genes. After 6 months, the response to treatment was evaluated for a total of 80 patients. The authors discovered divergent gene signatures between different TNFi, suggesting a potentially different mechanism of action between them.

Furthermore, studies to assess the patient’s response to classical drugs, such as methotrexate, have been conducted. For instance, researchers obtained a 0.78 AUC value when training an L2-regularized logistic regression (i.e., Ridge) model, without clinical covariates, to predict responder patients according to EULAR criteria [121]. This score was achieved using gene transcripts expression ratio between 4 weeks and pre-treatment. RF and network-based models were also tested.

Another study tried to compare the performance of ML algorithms (i.e., Lasso, RF, XGBoost) with multivariable LR in the prediction of insufficient response to methotrexate in 355 RA patients, measured using DAS28-ESR > 3.2 [122]. Sixteen clinical, laboratory and genetic variables were initially considered. After feature selection with Lasso, six predictors (i.e., TJC, Health Assessment Questionnaire (HAQ), BMI, smoking, ESR) remained. Based on the AUC results, authors concluded that there was no benefit in using ML algorithms (i.e., AUC Lasso 0.76, AUC RF 0.71, AUC XGBoost 0.70), over logistic regression, AUC 0.77.

Treatment response to conventional synthetic DMARDs and radiographic progression were evaluated in 144 early RA patients with the aid of logistic and Lasso regression models [123]. The LR model was used to identify a minimal set of clinical predictors, while the Lasso model was used to identify genes that could improve the clinical model. From 16 baseline clinical covariates, 8 (e.g., rheumatoid factor titer, disease duration, DAS28) were selected following a stepwise variable selection. These eight clinical variables were combined with 46 gene variables. The best result, AUC = 0.93, was achieved after penalising the 54 variables with Lasso. Eight final variables remained (e.g., rheumatoid factor, SDC1, MMP10). Hierarchical clustering was also used for the identification of pathotype-specific gene expression markers.

Liu et al. [7] developed a TNF blocker treatment response (i.e., etanercept) predictive model after evaluating quantitative changes in IgG galactosylation, alone and in combination with AS associated SNPs. 92 patients were considered for that purpose. Up to eight ML models were developed with SVM, 0.87 AUC, and flexible discriminant analysis, 0.82 AUC, as the best-performing ones. As input features, several indexes such as BASDAI, BASFI, ESR were considered.

Treatment response to GCs, commonly used as the first-line therapy in patients with AOSD, has been studied by a research group from China [124]. The motivation of the investigators was to balance the side effects and the effectiveness of the treatment, considering clinical and laboratory features (i.e., four neutrophil extracellular traps proteins). With this in mind, the authors developed two SVM models. The first tried to assess whether the proteins could serve as biomarkers and the second to predict if the levels of these circulating proteins could predict treatment response in terms of resistance to low-dose GCs. To evaluate the second outcome, the authors categorised the treatment response into a binary variable with low and high GCs levels as the dependent variable. The AUC values for the first and the second outcomes were 0.88 and 0.91, respectively.

#### 3.6.3. Image

DL and ML algorithms to assess treatment response have also been developed in the context of radiological images. For instance, Chandrika et al. [125] presented an architecture to assess bisphosphonate response in 28 patients with chronic non-bacterial osteitis. The number of included patients was 28 and the number of pairs of images 55. The proposed architecture consists of two components followed by an ensemble method, which classifies scans as “improved”, “worse”, or “stable”. The first, used a pre-trained CNN, InceptionV3, to extract features, embeddings, and representations that were used in a linear logistic model to produce a probability score. The second component used unsupervised clustering techniques to label the images and SVM to produce a probability score. Although the results were not remarkable (i.e., low specificity and accuracy), presumably due to class imbalance and the low number of training examples, this study showed that rare RMDs research could also benefit from data science techniques.

### 3.7. Predictors identification

This section describes studies in which their main stated objective is the identification and discovery of predictors associated with a particular outcome (e.g., disease diagnosis, disease worsening). Studies that have as their main objective the creation of a predictive model and conduct predictors identification as a preliminary step are described elsewhere.

#### 3.7.1. Clinical and demographic

The study of pain in RMDs is closely related to the concept of healthy ageing and the QoL of individuals. Understanding pain, its mechanisms, and how it emerges is essential for the development of cost-effective care programmes. Pain-associated arthritis predictors have been studied in [126]. Using the J48 DT algorithm, researchers predicted pain from 5,721 arthritis patients, regardless of the arthritic condition, with a 0.86 accuracy. From an initial set of over 200 predictors, including demographic (e.g., income, race, employment, education), PROMs (e.g., Short Form 12 (SF-12)), laboratory (e.g., times tested for HbA1c) and socio-behavioural characteristics, researchers built the final predictive model with just 12 variables (e.g., diabetes, general health, education, moderate activities). Of them, the physical component summary score from the SF-12 was the most meaningful one. Due to the cross-sectional nature of the study, the authors could not determine whether the predictors identified were the cause or the effect of pain conditions, this is, causality.

Identifying disease-worsening predictors could be helpful in defining inclusion and exclusion criteria for clinical trials. The identification of SS disease worsening and mortality predictors was the aim of Becker et al. [127]. The SS disease worsening definition was agreed upon by an expert group that considered different clinical events, such as renal crisis, decreased forced vital capacity or death. A total of 42 variables were studied, including demographic variables (e.g., age, disease duration), laboratory parameters (e.g., ANA, anti-neutrophil cytoplasmic antibody), other medical speciality domain variables (e.g., digital ulcer, synovitis, dyspnoea), and the European Scleroderma Study Group activity index variables. 228 out of 706 patients met the criteria for disease worsening. Lasso towards with multiple missing data imputation techniques were applied to find the eight most relevant predictors. Of them, five were strongly associated with disease progression (i.e., age, active digital ulcers, CRP, lung fibrosis, and muscle weakness). The validation was evaluated with bootstrap, achieving a C-index of 0.705.

Despite the prevalence of chronic neck pain, the study of its risk factors is limited. Blood serum samples, from 128 matched patients (i.e., 54 cases and 54 controls), were analysed by researchers to identify the correlation between 12 variables (e.g., age, BMI, zinc, albumin) with chronic neck pain using an ANN [128]. Following a feature selection using univariate analysis, vitamin D and ferritin were used as input for the ANN model. The model achieved 0.85 accuracy, 0.86 sensitivity and 0.83 specificity.

#### 3.7.2. Molecular biomarkers

The pathogenesis mechanisms of many autoimmune diseases remain unknown. To shed some light on the development of a disease, some authors date back to the earliest stages of cell development for which aberrant methylation occurs. For instance, in [129], the authors explored epigenetic defects in B cell development patterns of SLE patients, and took advantage of statistical learning methods to identify the most informative genes involved in transitional B cells. Hence, RF, Lasso, and Ridge regression were used for classifying healthy from SLE patients (n = 80) in an epigenetic context, in which DNA methylation signatures and their relevance to patients’ ethnicity were considered. To this end, the algorithms were tested across ethnicity groups in an independent validation cohort. Flow cytometry and methylation arrays were used for the measurement of DNA methylation levels. Finally, the authors found 60 CpGs that reached genome-wide significance for methylation differences in SLE patients compared to controls.

Non-invasiveness and discriminatory power are among the most desirable characteristics of a biomarker. For example, investigators from [130] used a novel protein microarray to screen and quantify 1,000 urinary protein biomarkers of lupus nephritis. Of these, 17 were selected for ELISA validation in an independent cohort. Lasso and RF were used for variable selection, Bayesian networks were used to uncover interdependencies between clinical indices and the biomarkers; and PCA, t-SNE and graph-based clustering, for dimensionality reduction and representation. Eventually, eight of the identified biomarkers (e.g., urine Angptl4, L-selectin, TPP1) successfully distinguished active lupus nephritis patients from active non-renal lupus patients, obtaining an AUC that ranged from 0.65 to 0.96.

The study of differences in terms of biomarker composition between children and adults is a topic that could elucidate the pathophysiological mechanisms of diseases such as pSS. For instance, authors in [131] hypothesised that the saliva of pSS children would have chemokines, cytokines, and biomarkers similar to those of pSS adults. Under this hypothesis, the predictive power of chemokines, cytokines, and biomarkers in saliva from pSS patients (n = 11) and healthy controls (n = 16) was evaluated using eight different classifiers (i.e., SVM, RF, NB, Gaussian process, AdaBoost, LR). From an initial set of 105 biomarkers associated with lymphocyte and mononuclear cell functions, 43 predictors remained after applying five feature selection methods (e.g., correlation, information gain ratio). Of them, 35 had previously been reported to be present in adult pSS patients. The predictors were grouped into sets of features. In a further step, hierarchical clustering and PCA validation were performed. Once trained, the best-performing model was KNN with a 0.93 AUC value. This AUC was achieved with only two predictors: IL-27 and chemokine (C-C motif) ligands 4.

Early diagnosis of certain diseases, such as uveitis, becomes even more important when delayed diagnosis can lead to blindness. Riahi et al. [132] investigated the interactions of ERAP1 polymorphisms in developing Beçhet’s disease using a non-parametric data mining technique, able to detect gene-gene or SNP-SNP interactions, called model-based multifactor dimensionality reduction (MB-MDR). The authors included 1,524 matched patients, 748 cases and 776 controls, collected a peripheral blood sample, and evaluated eleven SNPs. After applying MB-MDR, authors found a SNP, TT genotype of rs1065407, that had a significant synergistic effect on the disease (i.e., the allele increase the disease risk); and two SNPs, TT genotype of rs30187 and AA genotype of rs469876, that had significant antagonistic effects on Beçhet’s disease.

Moreover, researchers in [133], also used serum samples to search for potential miRNAs biomarkers in patients with Behçet’s disease (n = 10), sarcoidosis (n = 17) and Vogt–Koyanagi–Harada disease (n = 13). After using PCA to discriminate the different biological samples, the authors used RF to generate a variable importance measure. As a result, the researchers were able to identify three miRNAs (i.e., miR-4708-3p, miR-4323, and let-7g-3p) as the best predictors for each of the diseases studied, with AUC values of 0.96, 0.85 and 0.82 respectively.

One of the main objectives of the authors in [134], was to evaluate the predictive ability of inflammatory biomarkers along with other predictors (e.g., the severity of depression and anxiety) in patients with fibromyalgia. To this end, three different MLP models were trained to predict the Widespread Pain Index, the Symptom Severity Scale and the fibromyalgia diagnosis with 11 predictors (e.g., four immune biomarkers such as IL-6, IL-10; components of anxiety and depression scales, comorbidities, aerobic activity). With an AUC value of 0.91, the quality of sleep, the perceived stress scale, and the hospital anxiety were among the most predictive variables for a diagnosis of fibromyalgia.

Circulating protein biomarkers, capable of distinguishing between active vasculitis and remission in GCA (n=60), TAK (n=29), polyarteritis nodosa (n=26) and eosinophilic granulomatosis with polyangiitis patients (n=37), have been identified in a study conducted by the Vasculitis Clinical Research Consortium [135]. In this study, 22 serum proteins (e.g., IL-6, IL-8, IL-15, IFN-*γ*), potentially linked to vasculitis, were measured from samples collected during active and remission periods. A decision tree algorithm, J48, was used to identify biomarkers capable of distinguishing between active and inactive GCA, obtaining an accuracy of 0.31 and 0.87 respectively.

### 3.8. Illustrative studies

Table 5 shows a subcollection of the revised research articles that exemplifies the wide variety of RMDs studies in which AI has been an essential tool. This table structure is inspired by [33]. With the articles presented in this table, more than 20 different diseases are covered, exhibiting the following:

- AI techniques can be used for multiple purposes: as the tools necessary to perform the primary statistical analysis or as complementary tools that help researchers achieve their main objectives. Supervised learning (classification, regression), unsupervised learning (clustering, topic modelling, dimension reduction, novelty detection, visualisation), reinforcement learning (recommendation systems), deep learning (computer vision) and other procedures and techniques, such as feature selection or transfer learning, are being used in RMDs research. For instance, Lasso, and tree-based algorithms (e.g., RF, DT, XGBoost) are commonly used supervised techniques, while PCA and clustering algorithms (e.g., k-means, GMM) unsupervised.
- The sample size of the input data can range from a few patients to thousands. Large cohorts of patients are not indispensable to take advantage of AI techniques in RMDs research studies. In addition, the input data can come from multiple sources: clinical and demographic data, gene data, image data, and wearable activity tracker and sensor data.
- The topics addressed with these techniques are varied, the algorithms used numerous, and the potential results promising. As an example, in rare diseases, AI techniques can also be useful and have a real impact, as they can be used to obtain new insights and findings from clinical notes.
- The 57% of the articles presented in Table 5 are published in rheumatology-specialised journals.

Some of the findings listed above have also been discussed in other reviews, such as [29].

## 4. Discussion and conclusion

In this review, we have explored the clinical and technical background of RMDs that motivate the employment of AI techniques for research. Different contributing factors, such as the longitudinal, multidimensional and heterogeneous nature of data seem to facilitate the adoption of such techniques. We have also introduced the review articles published since 2017. These articles have addressed multiple topics, all of them with a strong rheumatology component: wearable activity trackers, bioethical perspectives, RWD, DL and so on. Then, we performed a literature review considering four different sources. The articles retrieved in this review were classified into six thematic categories. For each category, we made a distinction depending on the main predictor type of the study. Furthermore, we provided a table with more than twenty illustrative studies that exemplify the wide variety of RMDs in which AI techniques have penetrated. We also mentioned the limitations of ML algorithms with a particular focus on rheumatology.

From the 91 articles finally included, we could appreciate some interesting findings. For example, it seems that the RMDs that account for the majority of the research studies in which data science techniques are used more assiduously are: OA, osteoporosis, RA and SLE. It is hypothesised that the following factors may contribute to that fact:

- RA is the most prevalent systemic autoimmune disease among rheumatic inflammatory musculoskeletal diseases [136], while the prevalence of OA is remarkable, being one of the leading causes of disability [137].
- As medical images can be used to diagnose and quantify the status and progression of a disease, especially in OA, computer vision and DL algorithms may have attracted researchers, with a special interest in those techniques.

In addition, PCA, dimensionality reduction, and Lasso, variable selection, techniques appear to be highly popular.

On the other hand, data mining techniques have made it possible to study and characterise rare diseases in a different way, obtaining valuable information and uncovering new patterns that probably would not have been discovered otherwise. For instance, unsupervised learning techniques, and more concretely clustering, have been decisive in characterising disease subgroups. Moreover, some research groups have recognised the potential of these techniques and have adopted them as relevant tools for knowledge extraction when studying the pathology of specific diseases. The Big Data Sjögren Project Consortium is an example of this [138].

AI adoption is growing yearly in rheumatology research, as shown by the trend in the number of publications retrieved in Medline in a five-year period. Not only common algorithms but also the latest advances are being employed. After the study inclusion period of this review, new approaches have blossomed and have been applied to RMDs research, highlighting the interest that AI raises in researchers. From FSL approaches for early RA prediction to the employment of SHAP and GBMT to stratify patients with RA according to the trend of disease activity [3]. Regarding the data science languages employed, R and Python are the most widespread, but others such as Matlab, Weka, SPSS, Stata or JMP have also been used. However, further efforts must be made to validate the models in independent cohorts. The EULAR points to consider highlighted in 2019 the importance of this issue [20]: *conclusions drawn from big data need independent validation (in other datasets) to overcome current limitations and to assure scientific soundness*. As shown in *Supplementary Excel File Included Articles*, this point to consider is not fully addressed, although it seems to be gaining relevance.

### 4.1. Limitations of machine learning algorithms

Many technical and ethical limitations of ML algorithms are not exclusive to clinical research but are also shared with other research fields. Focusing on the technical limitations, authors in [139] highlighted six main points that may affect the performance of a model, making a distinction between *bad algorithms* and *bad data*. Particularising the six points to the medical field:

1. **Insufficient training data**: autoimmune rheumatic diseases (e.g., mixed connective tissue disease, polymyositis, SS, vasculitis) with low prevalence may be particularly affected by this problem. The reduced number of cases can hinder researchers from drawing valid conclusions. Therefore, multi-centre studies and database sharing may be proposed as efficient approaches that may mitigate this problem. Technical efforts have also been made with different methods such as FSL. For instance, this type of ML approach has recently been applied to MRI images from RA patients [18].
2. **Non-representative training data**: algorithms not trained with representative data are likely to fail with unseen population instances. This translates into the need to consider patients with different demographic and clinical characteristics (e.g., ethnicity, educational level, and so on). Otherwise, the model will not be robust enough to ensure generalisation to new cases. In the clinical field, subtle differences, such as different patient management procedures and drugs employed between centres, may hinder the algorithm generalisation capability. A broader discussion is held in [140].
3. **Poor-quality data**: in contrast to the data generated in a clinical trial context where patients are thoroughly selected, and the recorded variables are perfectly defined; RWD is more complex, heterogeneous, noisy, less structured, and prone to missing data; with an increased risk of obtaining inaccurate results and erroneous conclusions [141].
4. **Irrelevant features**: feature engineering and feature extraction processes should ensure that the selected features are relevant to build statistical models. Sometimes, extracting clinical features is not straightforward, easily accessible (e.g., data protection regulation), or cost-efficient (e.g., genomic analysis). For example, in GWAS analysis there may be hundreds of thousands of gene variants, and only a few of them are useful to predict the patient’s disease state. Moreover, the addition of unnecessary features may degrade the performance of the model, adding noise to the data.
5. **Overfitting**: the lack of generalisation capability to new cases can lead to useless models. Gathering more training data to minimise this problem is not always easy in clinical practice (especially for rare diseases), and reducing noise and outliers can be time-consuming, when possible. The *high dimensionality problem*, which characterises genetic studies, can be especially exacerbated by overfitting. Furthermore, in low-dimensional data, overfitting can have a non-negligible impact if the relationship between the outcome and the set of predictor variables is not strong [142]. Regularisation techniques (e.g., feature selection, less flexible models, constraints) constitute the most immediate solution to fight overfitting.
6. **Underfitting**: when a model is unable to capture the underlying data structure, the quality of the results is diminished. This occurs when a wrong assumption about the dataset is done, and a relatively simple algorithm is used for capturing complex data patterns. A better feature engineering and a statistical model with more parameters can be used as a solution to this problem. Hence, linear models, widely used in the medical research field, should be complemented by other non-linear models when the data behaviour is unclear.

Focusing on ethical limitations, authors in [143] provided an in-depth review to describe ongoing efforts and challenges showing a pipeline of ethical ML in health. These ethical limitations have a direct impact on the already discussed technical limitations. Some of the points addressed in that review are:

1. **Problem selection**: if the proposed research questions focus on the health needs of advantaged groups, disparities and injustices may occur. Therefore, racial, gender and global health injustices should be considered as factors that may influence the selection of a research problem. These disparities have been identified in different RMDs, such as RA [144], OA [145], gout [146] and specifically in SLE, which disproportionately affects women and minorities [147, 148]. AI models that also include disadvantaged groups should be promoted, for better model generalisation and for social inclusion.
2. **Data collection**: data on group membership (i.e., race and ethnicity) are not always collected. This may hamper model bias evaluation on different ethnic groups. Moreover, models trained on imbalanced data may perform worse on specific population groups and, therefore, those would benefit to a lesser extent from the advances of AI in healthcare. Noise and data loss may occur at different levels. For instance, in RCT, the study cohorts might not be representative of general patient populations. On its behalf, in EHR, stigmatising language collected in free text notes can have an impact on language models [149], leading to biased results [150]. Finally, population-specific data losses (e.g., undocumented patients with limited access to public healthcare services) should be taken into account when evaluating the generalisability of a model, and when studying the mechanisms of missing data (e.g., missing data not at random).

Other specific limitations related to regulatory, replicability and reproducibility that gain special relevance in the medical research field, have been identified by some authors [35, 151]:

1. **Strict and complex regulatory requirements**, especially in the model implementation and adoption phases.
2. **Lack of standardisation**: data acquisition (e.g., manually, or digitally), and different medical procedures or treatment guidelines may complicate and slow the use of multiple databases to train algorithms; introducing heterogeneity and obstructing preprocessing steps. Multicentre studies should consider the different variables, QoL questionnaires (e.g., SF-12, EuroQol5D), and the variable measurement employed (e.g., mmol/L, mg/dl) in each participant centre to have comparable data.
3. **External validation**: the performance of the models should be evaluated on different validation cohorts to extract reliable conclusions. Otherwise, the generalisation ability of the model will probably remain unclear or perform poorly; however, this is not always possible. Cross-validation, bootstrapping, or unsupervised techniques may be useful when the external validation is not guaranteed.
4. **Explainability**: most flexible statistical learning algorithms, such as NN are frequently considered “black-box” models. This raises ethical issues, such as how findings that may have an impact on a patient’s health status can be applied without a clear perspective of why and how the model works the way it does. In general, when building prediction models, the researcher’s objective is to obtain the smallest set of characteristics capable of predicting an outcome with the highest predictive performance [152]. Easy, understandable, economical, and accessible predictors are usually preferred, since well-performing models are expected to be deployed on a large scale, regardless of the means or resources of the different centres or countries. When choosing and deploying the different algorithms, the researcher must consider a trade-off between interpretability and flexibility. Interpretable algorithms are preferred when the main goal is to find associations between a set of predictors and a dependent variable, as well as in inference studies. When the aim of the researcher is to build the best predictive or classification model, the chosen algorithm might be non-linear and/or complex, and therefore, end with an unexplainable model but with good performance. This limitation has recently been discussed in [153, 154], but the debate remains open, as the authors pointed out: *Black-box medical practice hinders clinicians from assessing the quality of model inputs and parameters. If clinicians cannot understand the decision-making, they might violate patients’ rights to informed consent and autonomy*. Finally, some technical efforts have been made to improve the explainability, such as SHAP.

Biases may appear at different stages of a research involving AI techniques if the limitations presented above are not adequately addressed. In [155], a thorough study of those biases at three levels (i.e., data-driven, algorithmic, and human bias) is presented.

The European Commission (EC) has been working on some of the issues listed above for years [156], and it is about to publish the first-ever legal framework on AI [157]. Expanding on this topic, the first Ethics Guidelines for Trustworthy Artificial Intelligence [158] created by the EC have highlighted seven key requirements for AI systems to be deemed trustworthy (i.e., human agency and oversight; technical robustness and safety; privacy and data governance; transparency; diversity non-discrimination and fairness; societal and environmental wellbeing; and accountability). How to apply them in the healthcare domain has been explained in [159]. The UK government has recently published the first National AI Strategy Action Plan [160]. Besides, reporting guidelines on the use of AI in healthcare to ensure fair and transparent research have been recently defined, such as TRIPOD-AI and PROBAST-AI [161], SPIRIT-AI and CONSORT-AI [162, 163] or STARD-AI [164]. A recently published review article has tackled the adherence in diagnostic and prognostic applications of ML in SLE patients using TRIPOD and PROBAST [165]. A more detailed description of these reporting guidelines is addressed in [166]. Finally, other recently published articles have provided a detailed outline of more specific limitations and points to consider that apply to RMDs [22, 20, 33].

### 4.2. Limitations of the review

This review article has some limitations:

- The keywords used during the search in the different sources may omit some potential articles. For instance, common acronyms AI or ML were not used and the keywords employed were not exactly the same in the different sources. As this was a general overview of the state-of-the-art, disease-specific searches were not conducted (e.g., replacing the terms *rheumatology, rheumatic and musculoskeletal* in the different queries with terms associated with a specific disease: RA, rheumatoid arthritis and so on). This may limit the number of articles retrieved. In addition, the rheumatology journals search may introduce some bias since this search was limited to Q1 and Q2 journals of a specific year. Furthermore, articles without a PMID were excluded. However, by combining four different data sources, we tried to reduce this shortcoming. Eventually, due to the usage of broad keywords that encompassed a wide range of categories, a considerable number of articles had to be excluded because they were not closely related to RMDs. This demonstrates the weakness of the terms used to find specific articles, and the use of more specific keywords would be recommended to reduce the number of false positives.
- Some readers may miss an introduction to the different learning methods, so the potentially interested audience in this review may be shortened. As explained in the Introduction section, this was made to maximise the discussion of the articles. However, we provided enough references for researchers interested in deepening the technical background, and we also provided a short description of the methods in the *Supplementary Excel File Statistical Methods*. Eventually, we used the *data science* and AI terms indistinctly. This is not entirely correct, as subtle differences exist.
- Numerous studies on data mining techniques in RMDs research have been published from the state-of-the-art cutoff date (i.e., February, 22^th^ 2021) to the date of this manuscript submission. From studies that pursue to distinguish PsA, seronegative, and seropositive RA patients based on hand MRI using an ANN [167], to studies that examine the validity of ML models in predicting GCA flares after GCs tapering [168]. However, the review presented here addresses the main topics in a detailed way, providing a detailed overview for researchers who want to apply AI in RMDs, regardless of this two-year gap.
- The classification proposed into six main topics, may not be suitable for capturing subtle differences between articles. In fact, establishing the topic of an article following this classification is sometimes arduous, as the frontier of the different topics is fuzzy. For instance, the *disease classification* and the *disease prediction* categories differ only in the presence of healthy and sick patients, rather than groups of patients with different pathologies. However, since the number of approaches in which *disease classification* is lower and less studied, we tried to give enough relevance to this particular case. We have also faced this issue when trying to assign articles to the *predictors identification* and *disease progression and activity* topics.

### 4.3. Conclusion

Recent regulation efforts; reporting guidelines for fair, trustworthy, and transparent research; ethical considerations and technical efforts may have played a crucial role in the growing adoption of AI by the rheumatology research community. The use of these techniques has not been restricted to specific RMDs but has been used in both autoimmune and non-autoimmune diseases. Besides, these techniques have not been limited to a specific data type or source, but have been applied to both structured and unstructured data, as well as to data coming from different sources. The growing interest in such techniques suggests that new advances and groundbreaking approaches are expected to be adopted in the following years by new and experimented research groups.

## A. Appendix

**Table 5:**
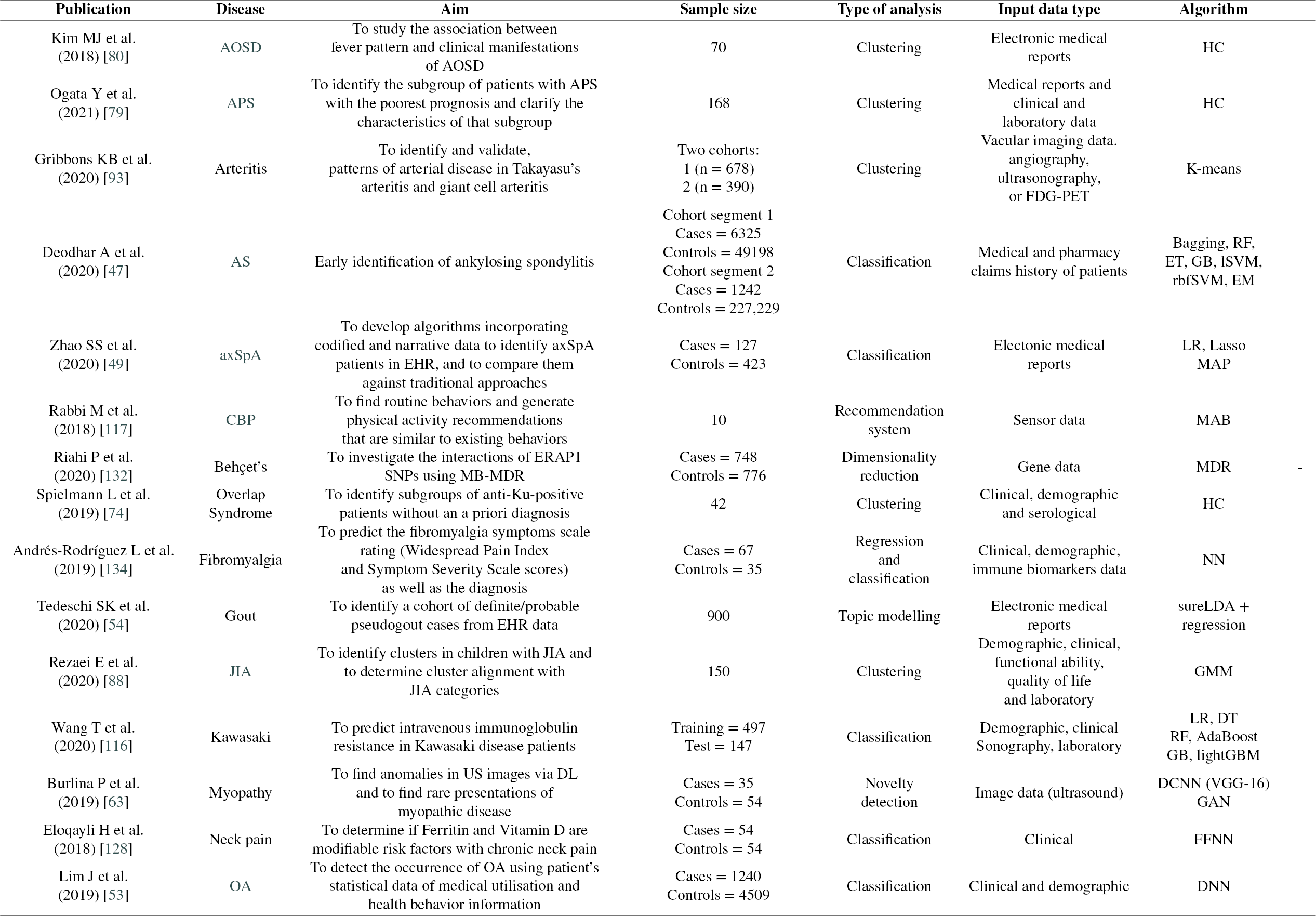
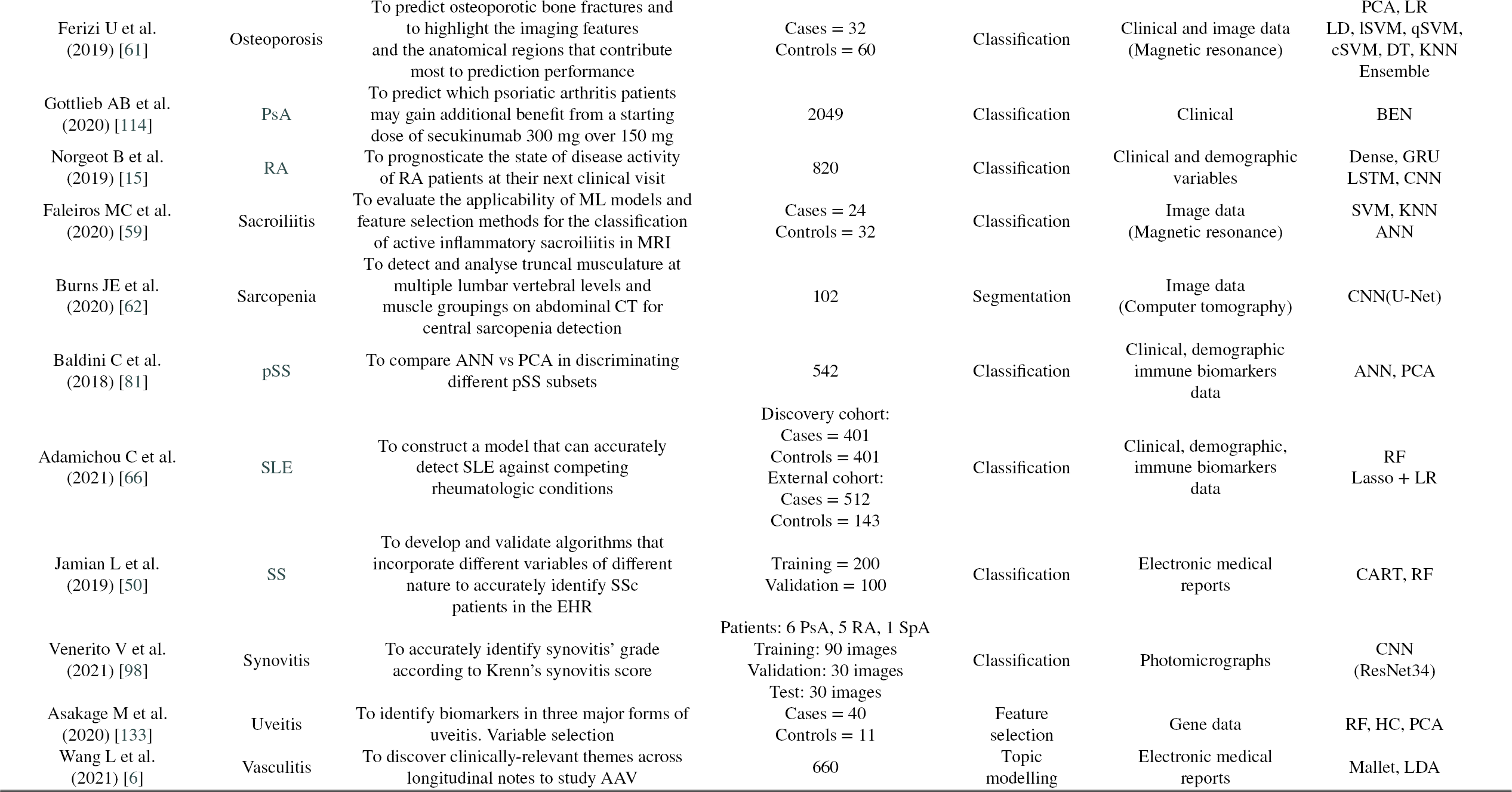
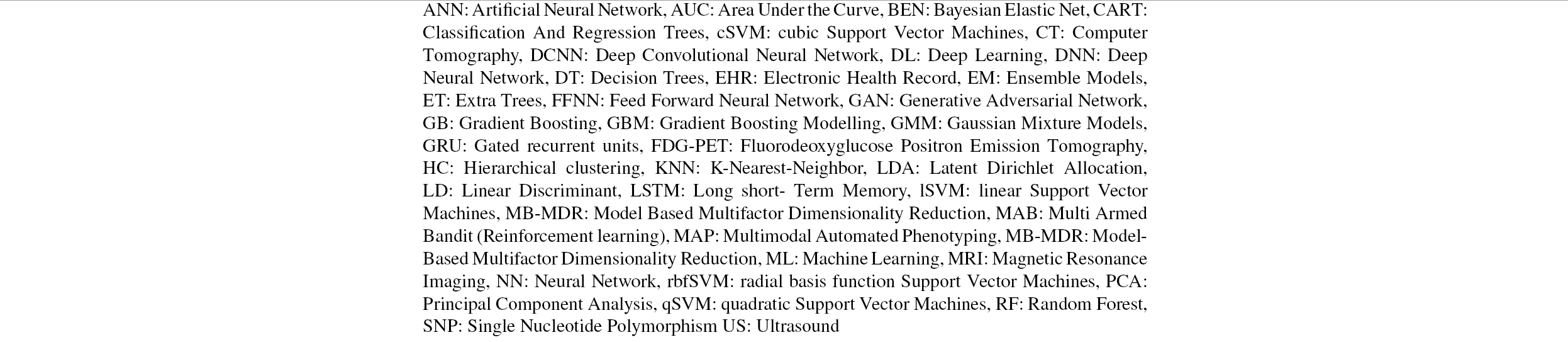
Examples of data mining techniques applied to different RMDs. Illustrative research articles

## CRediT authorship contribution statement

**Alfredo Madrid-García:** Conceptualization of this study, methodology, review, writing (original draft preparation). **Beatriz Merino-Barbancho:** Methodology, writing (original draft preparation). **Alejandro Rodríguez-González:** Conceptualization of this study. **Benjamín Fernández-Gutiérrez:** Conceptualization of this study. **Luis Rodríguez-Rodríguez:** Conceptualization of this study, methodology, review, writing (original draft preparation). **Ernestina Menasalvas-Ruiz:** Conceptualization of this study, methodology, writing (original draft preparation).

All of the authors were involved in the drafting and/or revising and/or publishing the manuscript.

**Acknowledgments**: **Lydia Abásolo-Alcázar** for her continuous feedback.

**Others**: ChatGPT was used to provide a brief description of the AI methods mentioned in this work in *Supplementary Excel File Statistical Methods*.

## Supporting information

Supplemental Excel File Acronyms

Supplemental Excel File Included Articles

Supplemental Excel File Unique Articles

Supplemental File Categories

## Data Availability

All data produced in the present work are contained in the manuscript

## Notes

This work was supported by the Instituto de Salud Carlos III, Ministry of Health, Madrid, Spain [RD21/002/0001]. The sponsor or funding organization had no role in the design or conduct of this research. The journal’s Fee was funded by the institution employing the senior author of the manuscript (Fundación Biomédica del Hospital Clínico San Carlos)

The authors declare there are no competing interests

### Competing Interest Statement

The authors have declared no competing interest.

### Funding Statement

This work was supported by the Instituto de Salud Carlos III, Ministry of Health, Madrid, Spain [RD21/002/0001]

### Summary of Updates

content restructuring

## Reference

[1] Désirée van der Heijde, David I Daikh, Neil Betteridge, Gerd R Burmester, Afton L Hassett, Eric L Matteson, Ronald van Vollenhoven, and Sharad Lakhanpal. Common language description of the term rheumatic and musculoskeletal diseases (rmds) for use in communication with the lay public, healthcare providers and other stakeholders endorsed by the european league against rheumatism (eular) and the american college of rheumatology (acr). Annals of the Rheumatic Diseases, 77(6):829–832, 2018. ISSN 0003-4967. doi: 10.1136/annrheumdis-2017-212565. URL https://ard.bmj.com/content/77/6/829.

[2] Stephen Bevan. Economic impact of musculoskeletal disorders (MSDs) on work in Europe. Best Practice & Research: Clinical Rheumatology, 29(3):356–373, 2015. ISSN 1521-6942. doi: http://dx.doi.org/10.1016/j.berh.2015.08.002. URL https://www.clinicalkey.com/#!/content/1-s2.0-S1521694215000947.

[3] Bon San Koo, Seongho Eun, Kichul Shin, Seokchan Hong, Yong-Gil Kim, Chang-Keun Lee, Bin Yoo, and Ji Seon Oh. Differences in trajectory of disease activity according to biologic and targeted synthetic disease-modifying anti-rheumatic drug treatment in patients with rheumatoid arthritis. Arthritis Research & Therapy, 24:233, 10 2022. ISSN 1478-6362. doi: 10.1186/s13075-022-02918-3.

[4] Jutta Richter, Christina Kampling, and Matthias Schneider. *Electronic Patient-Reported Outcome Measures (ePROMs) in Rheumatology*, pages 371–388. Springer International Publishing, Cham, 2016. ISBN 978-3-319-32851-5. doi: 10.1007/978-3-319-32851-5_15. URL https://doi.org/10.1007/978-3-319-32851-5_15.

[5] Rachel Knevel and Katherine P Liao. From real-world electronic health record data to real-world results using artificial intelligence. Annals of the Rheumatic Diseases, 2022. ISSN 0003-4967. doi: 10.1136/ard-2022-222626. URL https://ard.bmj.com/content/early/2022/09/23/ard-2022-222626.

[6] Liqin Wang, Eli Miloslavsky, John H. Stone, Hyon K. Choi, Li Zhou, and Zachary S. Wallace. Topic modeling to characterize the natural history of anca-associated vasculitis from clinical notes: A proof of concept study. Seminars in Arthritis and Rheumatism, 51(1):150–157, 2021. ISSN 0049-0172. doi: https://doi.org/10.1016/j.semarthrit.2020.10.012. URL https://www.sciencedirect.com/science/article/pii/S0049017220303115.

[7] Jing Liu, Qi Zhu, Jing Han, Hui Zhang, Yuan Li, Yanyun Ma, Dongyi He, Jianxin Gu, Xiaodong Zhou, John D Reveille, Li Jin, Hejian Zou, Shifang Ren, and Jiucun Wang. IgG Galactosylation status combined with MYOM2-rs2294066 precisely predicts anti-TNF response in ankylosing spondylitis. Molecular Medicine, 25(1):25, 2019. ISSN 1528-3658. doi: 10.1186/s10020-019-0093-2. URL https://doi.org/10.1186/s10020-019-0093-2.

[8] Laure Gossec, Frédéric Guyard, Didier Leroy, Thomas Lafargue, Michel Seiler, Charlotte Jacquemin, Anna Molto, Jérémie Sellam, Violaine Foltz, Frédérique Gandjbakhch, Christophe Hudry, Stéphane Mitrovic, Bruno Fautrel, and Hervé Servy. Detection of flares by decrease in physical activity, collected using wearable activity trackers in rheumatoid arthritis or axial spondyloarthritis: An application of machine learning analyses in rheumatology. Arthritis Care & Research, 71(10):1336–1343, 2019. doi: https://doi.org/10.1002/acr.23768. URL.

[9] Lindsey C. McKernan, Matthew C. Lenert, Leslie J. Crofford, and Colin G. Walsh. Outpatient engagement and predicted risk of suicide attempts in fibromyalgia. Arthritis Care & Research, 71(9):1255–1263, 2019. doi: https://doi.org/10.1002/acr.23748. URL https://onlinelibrary.wiley.com/doi/abs/10.1002/acr.23748.

[10] Oliver S. Burren, Guillermo Reales, Limy Wong, John Bowes, James C. Lee, Anne Barton, Paul A. Lyons, Kenneth G. C. Smith, Wendy Thomson, Paul D. W. Kirk, and Chris Wallace. Genetic feature engineering enables characterisation of shared risk factors in immune-mediated diseases. Genome Medicine, 12:106, 12 2020. ISSN 1756-994X. doi: 10.1186/s13073-020-00797-4.

[11] Anas Z. Abidin, Botao Deng, Adora M. DSouza, Mahesh B. Nagarajan, Paola Coan, and Axel Wismüller. Deep transfer learning for characterizing chondrocyte patterns in phase contrast x-ray computed tomography images of the human patellar cartilage. Computers in Biology and Medicine, 95:24–33, 2018. ISSN 0010-4825. doi: https://doi.org/10.1016/j.compbiomed.2018.01.008. URL https://www.sciencedirect.com/science/article/pii/S0010482518300167.

[12] Valentina Pedoia, Berk Norman, Sarah N. Mehany, Matthew D. Bucknor, Thomas M. Link, and Sharmila Majumdar. 3d convolutional neural networks for detection and severity staging of meniscus and pfj cartilage morphological degenerative changes in osteoarthritis and anterior cruciate ligament subjects. Journal of Magnetic Resonance Imaging, 49(2):400–410, 2019. doi: https://doi.org/10.1002/jmri.26246. URL https://onlinelibrary.wiley.com/doi/abs/10.1002/jmri.26246.

[13] José M. Lezcano-Valverde, Fernando Salazar, Leticia León, Esther Toledano, Juan A. Jover, Benjamín Fernandez-Gutierrez, Eduardo Soudah, Isidoro González-Álvaro, Lydia Abasolo, and Luis Rodriguez-Rodriguez. Development and validation of a multivariate predictive model for rheumatoid arthritis mortality using a machine learning approach. Scientific Reports, 7(1):10189, 12 2017. ISSN 2045-2322. doi: 10.1038/s41598-017-10558-w. URL http://www.nature.com/articles/s41598-017-10558-w.

[14] Yuyu Ishimoto, Amir Jamaludin, Cyrus Cooper, Karen Walker-Bone, Hiroshi Yamada, Hiroshi Hashizume, Hiroyuki Oka, Sakae Tanaka, Noriko Yoshimura, Munehito Yoshida, Jill Urban, Timor Kadir, and Jeremy Fairbank. Could automated machine-learned mri grading aid epidemiological studies of lumbar spinal stenosis? validation within the wakayama spine study. BMC Musculoskeletal Disorders, 21:158, 12 2020. ISSN 1471-2474. doi: 10.1186/s12891-020-3164-1.

[15] Beau Norgeot, Benjamin S. Glicksberg, Laura Trupin, Dmytro Lituiev, Milena Gianfrancesco, Boris Oskotsky, Gabriela Schmajuk, Jinoos Yazdany, and Atul J. Butte. Assessment of a Deep Learning Model Based on Electronic Health Record Data to Forecast Clinical Outcomes in Patients With Rheumatoid Arthritis. JAMA Network Open, 2(3):e190606–e190606, 03 2019. ISSN 2574-3805. doi: 10.1001/jamanetworkopen.2019.0606. URL https://doi.org/10.1001/jamanetworkopen.2019.0606.

[16] Yanshan Wang, Yiqing Zhao, Terry M. Therneau, Elizabeth J. Atkinson, Ahmad P. Tafti, Nan Zhang, Shreyasee Amin, Andrew H. Limper, Sundeep Khosla, and Hongfang Liu. Unsupervised machine learning for the discovery of latent disease clusters and patient subgroups using electronic health records. Journal of Biomedical Informatics, 102:103364, 2020. ISSN 1532-0464. doi: https://doi.org/10.1016/j.jbi.2019.103364. URL https://www.sciencedirect.com/science/article/pii/S1532046419302849.

[17] Clayton A. Turner, Alexander D. Jacobs, Cassios K. Marques, James C. Oates, Diane L. Kamen, Paul E. Anderson, and Jihad S. Obeid. Word2Vec inversion and traditional text classifiers for phenotyping lupus. BMC Medical Informatics and Decision Making, 17(1):126, 12 2017. ISSN 1472-6947. doi: 10.1186/s12911-017-0518-1. URL https://bmcmedinformdecismak.biomedcentral.com/articles/10.1186/s12911-017-0518-1.

[18] Yanli Li, Denis P. Shamonin, Tahereh Hassanzadeh, Monique Reijnierse, Annette H.M. van der Helm-van Mil, and Berend Stoel. A simple but effective training process for the few-shot prediction task of early rheumatoid arthritis from MRI. In Medical Imaging with Deep Learning, 2022. URL https://openreview.net/forum?id=8fk23e6ftYP.

[19] Jun-hee Kim. Search for medical information and treatment options for musculoskeletal disorders through an artificial intelligence chatbot: Focusing on shoulder impingement syndrome. medRxiv, 2022. doi: 10.1101/2022.12.16.22283512. URL https://www.medrxiv.org/content/early/2022/12/19/2022.12.16.22283512.

[20] Laure Gossec, Joanna Kedra, Hervé Servy, Aridaman Pandit, Simon Stones, Francis Berenbaum, Axel Finckh, Xenofon Baraliakos, Tanja A Stamm, David Gomez-Cabrero, Christian Pristipino, Remy Choquet, Gerd R Burmester, and Timothy R D J Radstake. Eular points to consider for the use of big data in rheumatic and musculoskeletal diseases. Annals of the Rheumatic Diseases, 79(1):69–76, 2020. ISSN 0003-4967. doi: 10.1136/annrheumdis-2019-215694. URL https://ard.bmj.com/content/79/1/69.

[21] Jeff Boissoneault, Landrew Sevel, Janelle Letzen, Michael Robinson, and Roland Staud. Biomarkers for musculoskeletal pain conditions: Use of brain imaging and machine learning. Current Rheumatology Reports, 19:5, 1 2017. ISSN 1523-3774. doi: 10.1007/s11926-017-0629-9.

[22] Joanna Kedra, Timothy Radstake, Aridaman Pandit, Xenofon Baraliakos, Francis Berenbaum, Axel Finckh, Bruno Fautrel, Tanja A Stamm, David Gomez-Cabrero, Christian Pristipino, Remy Choquet, Hervé Servy, Simon Stones, Gerd Burmester, and Laure Gossec. Current status of use of big data and artificial intelligence in rmds: a systematic literature review informing eular recommendations. RMD Open, 5(2), 2019. doi: 10.1136/rmdopen-2019-001004. URL https://rmdopen.bmj.com/content/5/2/e001004.

[23] Aridaman Pandit and Timothy R. D. J. Radstake. Machine learning in rheumatology approaches the clinic. Nature Reviews Rheumatology, 16:69–70, 2 2020. ISSN 1759-4790. doi: 10.1038/s41584-019-0361-0.

[24] Tagkopoulos Ilias Kim Ki-Jo. Application of machine learning in rheumatic disease research. Korean J Intern Med, 34(4):708–722, 2019. doi: 10.3904/kjim.2018.349. URL http://www.kjim.org/journal/view.php?number=170155.

[25] Afshin Jamshidi, Jean-Pierre Pelletier, and Johanne Martel-Pelletier. Machine-learning-based patient-specific prediction models for knee osteoarthritis. Nature Reviews Rheumatology, 15(1):49–60, 1 2019. ISSN 1759-4790. doi: 10.1038/s41584-018-0130-5.

[26] Narendra N. Khanna, Ankush D. Jamthikar, Deep Gupta, Matteo Piga, Luca Saba, Carlo Carcassi, Argiris A. Giannopoulos, Andrew Nico-laides, John R. Laird, Harman S. Suri, Sophie Mavrogeni, A.D. Protogerou, Petros Sfikakis, George D. Kitas, and Jasjit S. Suri. Rheumatoid arthritis: Atherosclerosis imaging and cardiovascular risk assessment using machine and deep learning–based tissue characterization. Current Atherosclerosis Reports, 21:7, 2 2019. ISSN 1523-3804. doi: 10.1007/s11883-019-0766-x.

[27] Berend C. Stoel. Artificial intelligence in detecting early ra. Seminars in Arthritis and Rheumatism, 49(3, Supplement):S25–S28, 2019. ISSN 0049-0172. doi: https://doi.org/10.1016/j.semarthrit.2019.09.020. URL https://www.sciencedirect.com/science/article/pii/S0049017219306559. Advances in Targeted Therapies: Proceedings of the 2019 Meeting.

[28] Uran Ferizi, Stephen Honig, and Gregory Chang. Artificial intelligence, osteoporosis and fragility fractures. Current Opinion in Rheumatology, 31:368–375, 7 2019. ISSN 1040-8711. doi: 10.1097/BOR.0000000000000607.

[29] Mengdi Jiang, Yueting Li, Chendan Jiang, Lidan Zhao, Xuan Zhang, and Peter E Lipsky. Machine Learning in Rheumatic Diseases. Clinical Reviews in Allergy & Immunology, 60(1):96–110, 2021. ISSN 1559-0267. doi: 10.1007/s12016-020-08805-6. URL https://doi.org/10.1007/s12016-020-08805-6.

[30] Maria Hügle, Patrick Omoumi, Jacob M van Laar, Joschka Boedecker, and Thomas Hügle. Applied machine learning and artificial intelligence in rheumatology. Rheumatology Advances in Practice, 4(1), 02 2020. ISSN 2514-1775. doi: 10.1093/rap/rkaa005. URL https://doi.org/10.1093/rap/rkaa005.rkaa005.

[31] Amaranta Manrique de Lara and Ingris Peláez-Ballestas. Big data and data processing in rheumatology: bioethical perspectives. Clinical Rheumatology, 39:1007–1014, 2020. ISSN 1434-9949. doi: 10.1007/s10067-020-04969-w. URL https://doi.org/10.1007/s10067-020-04969-w.

[32] Berend Stoel. Use of artificial intelligence in imaging in rheumatology – current status and future perspectives. RMD Open, 6(1), 2020. doi: 10.1136/rmdopen-2019-001063. URL https://rmdopen.bmj.com/content/6/1/e001063.

[33] Kathryn M Kingsmore, Christopher E Puglisi, Amrie C Grammer, and Peter E Lipsky. An introduction to machine learning and analysis of its use in rheumatic diseases. Nature Reviews Rheumatology, 2021. ISSN 1759-4804. doi: 10.1038/s41584-021-00708-w. URL https://doi.org/10.1038/s41584-021-00708-w.

[34] David Soriano-Valdez, Ingris Pelaez-Ballestas, Amaranta Manrique de Lara, and Alfonso Gastelum-Strozzi. The basics of data, big data, and machine learning in clinical practice. Clinical Rheumatology, 40:11–23, 1 2021. ISSN 0770-3198. doi: 10.1007/s10067-020-05196-z.

[35] Joanna Kedra, Thomas Davergne, Ben Braithwaite, Hervé Servy, and Laure Gossec. Machine learning approaches to improve disease management of patients with rheumatoid arthritis: review and future directions. Expert Review of Clinical Immunology, 0(ja):null, 2021. doi: 10.1080/1744666X.2022.2017773. URL https://doi.org/10.1080/1744666X.2022.2017773. PMID: 34890271.

[36] Julien Smets, Enisa Shevroja, Thomas Hügle, William D Leslie, and Didier Hans. Machine learning solutions for osteoporosis—a review. Journal of Bone and Mineral Research, 36:833–851, 5 2021. ISSN 0884-0431. doi: 10.1002/jbmr.4292.

[37] Thomas Davergne, Joanna Kedra, and Laure Gossec. Wearable activity trackers and artificial intelligence in the management of rheumatic diseases. Zeitschrift für Rheumatologie, 80:928–935, 12 2021. ISSN 0340-1855. doi: 10.1007/s00393-021-01100-5.

[38] Maxwell A. Konnaris, Matthew Brendel, Mark Alan Fontana, Miguel Otero, Lionel B. Ivashkiv, Fei Wang, and Richard D. Bell. Computational pathology for musculoskeletal conditions using machine learning: advances, trends, and challenges. Arthritis Research & Therapy, 24:68, 12 2022. ISSN 1478-6362. doi: 10.1186/s13075-021-02716-3.

[39] Marie Binvignat, Valentina Pedoia, Atul J Butte, Karine Louati, David Klatzmann, Francis Berenbaum, Encarnita Mariotti-Ferrandiz, and Jérémie Sellam. Use of machine learning in osteoarthritis research: a systematic literature review. RMD Open, 8(1), 2022. doi: 10.1136/rmdopen-2021-001998. URL https://rmdopen.bmj.com/content/8/1/e001998.

[40] Francesco Calivà, Nikan K. Namiri, Maureen Dubreuil, Valentina Pedoia, Eugene Ozhinsky, and Sharmila Majumdar. Studying osteoarthritis with artificial intelligence applied to magnetic resonance imaging. Nature Reviews Rheumatology, 18:112–121, 2 2022. ISSN 1759-4790. doi: 10.1038/s41584-021-00719-7.

[41] Yuan Li and Linru Zhao. Application of machine learning in rheumatic immune diseases. Journal of Healthcare Engineering, 2022:1–9, 1 2022. ISSN 2040-2309. doi: 10.1155/2022/9273641.

[42] Diederik De Cock, Elena Myasoedova, Daniel Aletaha, and Paul Studenic. Big data analyses and individual health profiling in the arena of rheumatic and musculoskeletal diseases (rmds). Therapeutic Advances in Musculoskeletal Disease, 14:1759720X2211059, 1 2022. ISSN 1759-720X. doi: 10.1177/1759720X221105978. URL http://journals.sagepub.com/doi/10.1177/1759720X221105978.

[43] Amanda E. Nelson and Liubov Arbeeva. Narrative review of machine learning in rheumatic and musculoskeletal diseases for clinicians and researchers: biases, goals, and future directions. The Journal of Rheumatology, 2022. ISSN 0315-162X. doi: 10.3899/jrheum.220326. URL https://www.jrheum.org/content/early/2022/07/14/jrheum.220326.

[44] Francesco Bonomi, Silvia Peretti, Gemma Lepri, Vincenzo Venerito, Edda Russo, Cosimo Bruni, Florenzo Iannone, Sabina Tangaro, Amedeo Amedei, Serena Guiducci, Marco Matucci Cerinic, and Silvia Bellando Randone. The use and utility of machine learning in achieving precision medicine in systemic sclerosis: A narrative review. Journal of Personalized Medicine, 12(8), 2022. ISSN 2075-4426. doi: 10.3390/jpm12081198. URL https://www.mdpi.com/2075-4426/12/8/1198.

[45] Christopher McMaster, Alix Bird, David FL Liew, Russell R Buchanan, Claire E Owen, Wendy W Chapman, and Douglas EV Pires. Artificial intelligence and deep learning for rheumatologists: A primer and review of the literature. Arthritis & Rheumatology, 2022. doi: https://doi.org/10.1002/art.42296. URL https://onlinelibrary.wiley.com/doi/abs/10.1002/art.42296.

[46] Antonio Martinez-Millana, Aida Saez-Saez, Roberto Tornero-Costa, Natasha Azzopardi-Muscat, Vicente Traver, and David Novillo-Ortiz. Artificial intelligence and its impact on the domains of universal health coverage, health emergencies and health promotion: An overview of systematic reviews. International Journal of Medical Informatics, 166:104855, 2022. ISSN 1386-5056. doi: https://doi.org/10.1016/j.ijmedinf.2022.104855. www.sciencedirect.com/science/article/pii/S1386505622001691.

[47] Atul Deodhar, Martin Rozycki, Cody Garges, Oodaye Shukla, Theresa Arndt, Tara Grabowsky, and Yujin Park. Use of machine learning techniques in the development and refinement of a predictive model for early diagnosis of ankylosing spondylitis. Clinical Rheumatology, 39(4):975–982, 2020. ISSN 1434-9949. doi: 10.1007/s10067-019-04553-x. URL https://doi.org/10.1007/s10067-019-04553-x.

[48] Jessica A. Walsh, Shaobo Pei, Gopi K. Penmetsa, Rebecca S. Overbury, Daniel O. Clegg, and Brian C. Sauer. Identifying patients with axial spondyloarthritis in large datasets: Expanding possibilities for observational research. The Journal of Rheumatology, 2020. ISSN 0315-162X. doi: 10.3899/jrheum.200570. URL https://www.jrheum.org/content/early/2021/01/25/jrheum.200570.

[49] Sizheng Steven Zhao, Chuan Hong, Tianrun Cai, Chang Xu, Jie Huang, Joerg Ermann, Nicola J Goodson, Daniel H Solomon, Tianxi Cai, and Katherine P Liao. Incorporating natural language processing to improve classification of axial spondyloarthritis using electronic health records. Rheumatology, 59(5):1059–1065, 09 2019. ISSN 1462-0324. doi: 10.1093/rheumatology/kez375. URL https://doi.org/10.1093/rheumatology/kez375.

[50] Lia Jamian, Lee Wheless, Leslie J Crofford, and April Barnado. Rule-based and machine learning algorithms identify patients with systemic sclerosis accurately in the electronic health record. Arthritis Research & Therapy, 21(1):305, 2019. ISSN 1478-6362. doi: 10.1186/s13075-019-2092-7. URL https://doi.org/10.1186/s13075-019-2092-7.

[51] Sicong Huang, Jie Huang, Tianrun Cai, Kumar P Dahal, Andrew Cagan, Zeling He, Jacklyn Stratton, Isaac Gorelik, Chuan Hong, Tianxi Cai, and Katherine P Liao. Impact of ICD10 and secular changes on electronic medical record rheumatoid arthritis algorithms. Rheumatology, 59 (12):3759–3766, 05 2020. ISSN 1462-0324. doi: 10.1093/rheumatology/keaa198. URL https://doi.org/10.1093/rheumatology/keaa198.

[52] Tjardo D Maarseveen, Timo Meinderink, Marcel J T Reinders, Johannes Knitza, Tom W J Huizinga, Arnd Kleyer, David Simon, Erik B van den Akker, and Rachel Knevel. Machine learning electronic health record identification of patients with rheumatoid arthritis: Algorithm pipeline development and validation study. JMIR Medical Informatics, 8:e23930, 11 2020. ISSN 2291-9694. doi: 10.2196/23930.

[53] Jihye Lim, Jungyoon Kim, and Songhee Cheon. A deep neural network-based method for early detection of osteoarthritis using statistical data. International Journal of Environmental Research and Public Health, 16(7), 2019. ISSN 1660-4601. doi: 10.3390/ijerph16071281. URL https://www.mdpi.com/1660-4601/16/7/1281.

[54] Sara K. Tedeschi, Tianrun Cai, Zeling He, Yuri Ahuja, Chuan Hong, Katherine A. Yates, Kumar Dahal, Chang Xu, Houchen Lyu, Kazuki Yoshida, Daniel H. Solomon, Tianxi Cai, and Katherine P. Liao. Classifying pseudogout using machine learning approaches with electronic health record data. Arthritis Care & Research, 73(3):442–448, 2021. doi: https://doi.org/10.1002/acr.24132. URL https://onlinelibrary.wiley.com/doi/abs/10.1002/acr.24132.

[55] Erika Van Nieuwenhove, Vasiliki Lagou, Lien Van Eyck, James Dooley, Ulrich Bodenhofer, Carlos Roca, Marijne Vandebergh, An Goris, Stéphanie Humblet-Baron, Carine Wouters, and Adrian Liston. Machine learning identifies an immunological pattern associated with multiple juvenile idiopathic arthritis subtypes. Annals of the Rheumatic Diseases, 78(5):617–628, 2019. ISSN 0003-4967. doi: 10.1136/annrheumdis-2018-214354. URL https://ard.bmj.com/content/78/5/617.

[56] Maria Camacho-Encina, Vanesa Balboa-Barreiro, Ignacio Rego-Perez, Florencia Picchi, Jennifer VanDuin, Ji Qiu, Manuel Fuentes, Natividad Oreiro, Joshua LaBaer, Cristina Ruiz-Romero, and Francisco J Blanco. Discovery of an autoantibody signature for the early diagnosis of knee osteoarthritis: data from the osteoarthritis initiative. Annals of the Rheumatic Diseases, 78(12):1699–1705, 2019. ISSN 0003-4967. doi: 10.1136/annrheumdis-2019-215325. URL https://ard.bmj.com/content/78/12/1699.

[57] Christophe Roncato, Lior Perez, Antoine Brochet-Guégan, Caroline Allix-Béguec, Alizée Raimbeau, Giovanni Gautier, Christian Agard, Gaëtan Ploton, Simon Moisselin, Fanny Lorcerie, Guillaume Denis, Bruno Gombert, Elisabeth Gervais, and Olivier Espitia. Colour Doppler ultrasound of temporal arteries for the diagnosis of giant cell arteritis: a multicentre deep learning study. Clinical and experimental rheumatology, 38 Suppl 1(2):120–125, 2020. ISSN 0392-856X. URL http://www.ncbi.nlm.nih.gov/pubmed/32441644.

[58] Riel Castro-Zunti, Eun Hae Park, Younhee Choi, Gong Yong Jin, and Seok bum Ko. Early detection of ankylosing spondylitis using texture features and statistical machine learning, and deep learning, with some patient age analysis. Computerized Medical Imaging and Graphics, 82:101718, 2020. ISSN 0895-6111. doi: https://doi.org/10.1016/j.compmedimag.2020.101718. URL https://www.sciencedirect.com/science/article/pii/S0895611120300215.

[59] Matheus Calil Faleiros, Marcello Henrique Nogueira-Barbosa, Vitor Faeda Dalto, José Raniery Ferreira Júnior, Ariane Priscilla Magalhães Tenório, Rodrigo Luppino-Assad, Paulo Louzada-Junior, Rangaraj Mandayam Rangayyan, and Paulo Mazzoncini de Azevedo-Marques. Machine learning techniques for computer-aided classification of active inflammatory sacroiliitis in magnetic resonance imaging. Advances in Rheumatology, 60:25, 12 2020. ISSN 2523-3106. doi: 10.1186/s42358-020-00126-8.

[60] Norio Yamamoto, Shintaro Sukegawa, Akira Kitamura, Ryosuke Goto, Tomoyuki Noda, Keisuke Nakano, Kiyofumi Takabatake, Hotaka Kawai, Hitoshi Nagatsuka, Keisuke Kawasaki, Yoshihiko Furuki, and Toshifumi Ozaki. Deep learning for osteoporosis classification using hip radiographs and patient clinical covariates. Biomolecules, 10(11), 2020. ISSN 2218-273X. doi: 10.3390/biom10111534. URL https://www.mdpi.com/2218-273X/10/11/1534.

[61] Uran Ferizi, Harrison Besser, Pirro Hysi, Joseph Jacobs, Chamith S. Rajapakse, Cheng Chen, Punam K. Saha, Stephen Honig, and Gregory Chang. Artificial intelligence applied to osteoporosis: A performance comparison of machine learning algorithms in predicting fragility fractures from mri data. Journal of Magnetic Resonance Imaging, 49(4):1029–1038, 2019. doi: https://doi.org/10.1002/jmri.26280. URL https://onlinelibrary.wiley.com/doi/abs/10.1002/jmri.26280.

[62] Joseph E. Burns, Jianhua Yao, Didier Chalhoub, Joseph J. Chen, and Ronald M. Summers. A machine learning algorithm to estimate sarcopenia on abdominal ct. Academic Radiology, 27(3):311–320, 2020. ISSN 1076-6332. doi: https://doi.org/10.1016/j.acra.2019.03.011. URL https://www.sciencedirect.com/science/article/pii/S1076633219301655.

[63] Philippe Burlina, Neil Joshi, Seth Billings, I-Jeng Wang, and Jemima Albayda. Deep embeddings for novelty detection in myopathy. Computers in Biology and Medicine, 105:46–53, 2019. ISSN 0010-4825. doi: https://doi.org/10.1016/j.compbiomed.2018.12.006. URL https://www.sciencedirect.com/science/article/pii/S0010482518304049.

[64] Suhanyaa Nitkunanantharajah, Katja Haedicke, Tonia B. Moore, Joanne B. Manning, Graham Dinsdale, Michael Berks, Christopher Taylor, Mark R. Dickinson, Dominik Jüstel, Vasilis Ntziachristos, Ariane L. Herrick, and Andrea K. Murray. Three-dimensional optoacoustic imaging of nailfold capillaries in systemic sclerosis and its potential for disease differentiation using deep learning. Scientific Reports, 10(1): 16444, 2020. ISSN 2045-2322. doi: 10.1038/s41598-020-73319-2. URL https://doi.org/10.1038/s41598-020-73319-2.

[65] Shan-Chi Yu, Kung-Chao Chang, Hsuan Wang, Meng-Fang Li, Tsung-Lin Yang, Chun-Nan Chen, Chih-Jung Chen, Ko-Chin Chen, Chieh-Yu Shen, Po-Yen Kuo, Long-Wei Lin, Yueh-Min Lin, and Wei-Chou Lin. Distinguishing lupus lymphadenitis from Kikuchi disease based on clinicopathological features and C4d immunohistochemistry. Rheumatology, 60(3):1543–1552, 11 2020. ISSN 1462-0324. doi: 10.1093/rheumatology/keaa524. URL https://doi.org/10.1093/rheumatology/keaa524.

[66] Christina Adamichou, Irini Genitsaridi, Dionysis Nikolopoulos, Myrto Nikoloudaki, Argyro Repa, Alessandra Bortoluzzi, Antonis Fanouriakis, Prodromos Sidiropoulos, Dimitrios T Boumpas, and George K Bertsias. Lupus or not? sle risk probability index (slerpi): a simple, clinician-friendly machine learning-based model to assist the diagnosis of systemic lupus erythematosus. Annals of the Rheumatic Diseases, 80(6):758–766, 2021. ISSN 0003-4967. doi: 10.1136/annrheumdis-2020-219069. URL https://ard.bmj.com/content/80/6/758.

[67] Michelle J. Ormseth, Joseph F. Solus, Quanhu Sheng, Fei Ye, Qiong Wu, Yan Guo, Annette M. Oeser, Ryan M. Allen, Kasey C. Vickers, and C. Michael Stein. Development and validation of a microrna panel to differentiate between patients with rheumatoid arthritis or systemic lupus erythematosus and controls. The Journal of Rheumatology, 2019. ISSN 0315-162X. doi: 10.3899/jrheum.181029. URL https://www.jrheum.org/content/early/2019/05/13/jrheum.181029.

[68] Xiao Liu, Wei Zhang, Ming Zhao, Longfei Fu, Limin Liu, Jinghua Wu, Shuangyan Luo, Longlong Wang, Zijun Wang, Liya Lin, Yan Liu, Shiyu Wang, Yang Yang, Lihua Luo, Juqing Jiang, Xie Wang, Yixin Tan, Tao Li, Bochen Zhu, Yi Zhao, Xiaofei Gao, Ziyun Wan, Cancan Huang, Mingyan Fang, Qianwen Li, Huanhuan Peng, Xiangping Liao, Jinwei Chen, Fen Li, Guanghui Ling, Hongjun Zhao, Hui Luo, Zhongyuan Xiang, Jieyue Liao, Yu Liu, Heng Yin, Hai Long, Haijing Wu, huanming Yang, Jian Wang, and Qianjin Lu. T cell receptor #x1D6FD; repertoires as novel diagnostic markers for systemic lupus erythematosus and rheumatoid arthritis. Annals of the Rheumatic Diseases, 78 (8):1070–1078, 2019. ISSN 0003-4967. doi: 10.1136/annrheumdis-2019-215442. URL https://ard.bmj.com/content/78/8/1070.

[69] Margarida Souto-Carneiro, Lilla Tóth, Rouven Behnisch, Konstantin Urbach, Karel D Klika, Rui A Carvalho, and Hanns-Martin Lorenz. Differences in the serum metabolome and lipidome identify potential biomarkers for seronegative rheumatoid arthritis versus psoriatic arthritis. Annals of the Rheumatic Diseases, 79(4):499–506, 2020. ISSN 0003-4967. doi: 10.1136/annrheumdis-2019-216374. URL https://ard.bmj.com/content/79/4/499.

[70] Juliana Imgenberg-Kreuz, Jonas Carlsson Almlöf, Dag Leonard, Christopher Sjöwall, Ann-Christine Syvänen, Lars Rönnblom, Johanna K. Sandling, and Gunnel Nordmark. Shared and unique patterns of dna methylation in systemic lupus erythematosus and primary sjögren’s syndrome. Frontiers in Immunology, 10, 2019. ISSN 1664-3224. doi: 10.3389/fimmu.2019.01686. URL https://www.frontiersin.org/articles/10.3389/fimmu.2019.01686.

[71] Yan Zhao, Bin Chen, Shufeng Li, Lanxiu Yang, Dequan Zhu, Ye Wang, Haiying Wang, Tao Wang, Bin Shi, Zhongtao Gai, Jun Yang, Xueyuan Heng, Junjie Yang, and Lei Zhang. Detection and characterization of bacterial nucleic acids in culture-negative synovial tissue and fluid samples from rheumatoid arthritis or osteoarthritis patients. Scientific Reports, 8:14305, 12 2018. ISSN 2045-2322. doi: 10.1038/s41598-018-32675-w.

[72] Mark Reed, Timothy Le Souëf, and Elliot Rampono. A pilot study of a machine-learning tool to assist in the diagnosis of hand arthritis. Internal Medicine Journal, 52(6):959–967, 2022. doi: https://doi.org/10.1111/imj.15173. URL https://onlinelibrary.wiley.com/doi/abs/10.1111/imj.15173.

[73] Shawli Bardhan and Mrinal Kanti Bhowmik. 2-Stage classification of knee joint thermograms for rheumatoid arthritis prediction in subclinical inflammation. Australasian Physical & Engineering Sciences in Medicine, 42(1):259–277, 2019. ISSN 1879-5447. doi: 10.1007/s13246-019-00726-9. URL https://doi.org/10.1007/s13246-019-00726-9.

[74] Lionel Spielmann, Benoit Nespola, François Séverac, Emmanuel Andres, Romain Kessler, Aurélien Guffroy, Vincent Poindron, Thierry Martin, Bernard Geny, Jean Sibilia, and Alain Meyer. Anti-ku syndrome with elevated ck and anti-ku syndrome with anti-dsdna are two distinct entities with different outcomes. Annals of the Rheumatic Diseases, 78(8):1101–1106, 2019. ISSN 0003-4967. doi: 10.1136/annrheumdis-2018-214439. URL https://ard.bmj.com/content/78/8/1101.

[75] Alain Meyer, Lionel Spielmann, and François Séverac. On how to not misuse hierarchical clustering on principal components to define clinically meaningful patient subgroups. response to: ‘on using machine learning algorithms to define clinical meaningful patient subgroups’ by pinal-fernandez and mammen. Annals of the Rheumatic Diseases, 79(10):e129–e129, 2020. ISSN 0003-4967. doi: 10.1136/annrheumdis-2019-215868. URL https://ard.bmj.com/content/79/10/e129.

[76] Iago Pinal-Fernandez and Andrew Lee Mammen. On using machine learning algorithms to define clinically meaningful patient subgroups. Annals of the Rheumatic Diseases, 79(10):e128–e128, 2020. ISSN 0003-4967. doi: 10.1136/annrheumdis-2019-215852. URL https://ard.bmj.com/content/79/10/e128.

[77] Alain Meyer, Lionel Spielmann, and François Séverac. Response to ‘augmented vs. artificial intelligence for stratification of patients with myositis’ by mahler et al. Annals of the Rheumatic Diseases, 79(12):e163–e163, 2020. ISSN 0003-4967. doi: 10.1136/annrheumdis-2019-216014. URL https://ard.bmj.com/content/79/12/e163.

[78] Michael Mahler, Brenden Rossin, and Olga Kubassova. Augmented versus artificial intelligence for stratification of patients with myositis. Annals of the Rheumatic Diseases, 79(12):e162–e162, 2020. ISSN 0003-4967. doi: 10.1136/annrheumdis-2019-216000. URL https://ard.bmj.com/content/79/12/e162.

[79] Yusuke Ogata, Yuichiro Fujieda, Masanari Sugawara, Taiki Sato, Naoki Ohnishi, Michihito Kono, Masaru Kato, Kenji Oku, Olga Amengual, and Tatsuya Atsumi. Morbidity and mortality in antiphospholipid syndrome based on cluster analysis: a 10-year longitudinal cohort study. Rheumatology, 60(3):1331–1337, 09 2020. ISSN 1462-0324. doi: 10.1093/rheumatology/keaa542. URL https://doi.org/10.1093/rheumatology/keaa542.

[80] Min Jung Kim, Eun Young Ahn, Woochang Hwang, Youngjo Lee, Eun Young Lee, Eun Bong Lee, Yeong Wook Song, and Jin Kyun Park. Association between fever pattern and clinical manifestations of adult-onset Still’s disease: unbiased analysis using hierarchical clustering. Clinical and experimental rheumatology, 36(6 Suppl 115):74–79, 2018. ISSN 0392-856X. doi: 30582502. URL http://www.ncbi.nlm.nih.gov/pubmed/30582502.

[81] C. Baldini, F. Ferro, N. Luciano, S. Bombardieri, and E. Grossi. Artificial neural networks help to identify disease subsets and to predict lymphoma in primary Sjögren’s syndrome. Clinical and Experimental Rheumatology, 36:S137–S144, 2018. ISSN 1593098X.

[82] Elena Bartoloni, Chiara Baldini, Francesco Ferro, Alessia Alunno, Francesco Carubbi, Giacomo Cafaro, Stefano Bombardieri, Roberto Gerli, and Enzo Grossi. Application of artificial neural network analysis in the evaluation of cardiovascular risk in primary Sjögren’s syndrome: A novel pathogenetic scenario? Clinical and Experimental Rheumatology, 37(3):S133–S139, 2019. ISSN 1593098X.

[83] Vasileios C. Pezoulas, Themis P. Exarchos, Athanasios G. Tzioufas, Salvatore De Vita, and Dimitrios I. Fotiadis. Predicting lymphoma outcomes and risk factors in patients with primary sjögren’s syndrome using gradient boosting tree ensembles. In 2019 41st Annual International Conference of the IEEE Engineering in Medicine and Biology Society (EMBC), pages 2165–2168, 20191. doi: 10.1109/EMBC.2019.8857557.

[84] Simon W. M. Eng, Florence A. Aeschlimann, Mira van Veenendaal, Roberta A. Berard, Alan M. Rosenberg, Quaid Morris, Rae S. M. Yeung, and on behalf of the ReACCh-Out Research Consortium. Patterns of joint involvement in juvenile idiopathic arthritis and prediction of disease course: A prospective study with multilayer non-negative matrix factorization. PLOS Medicine, 16(2):1–22, 02 2019. doi: 10.1371/journal.pmed.1002750. URL https://doi.org/10.1371/journal.pmed.1002750.

[85] Alessandra Tesser, Luciana Martins de Carvalho, Paula Sandrin-Garcia, Alessia Pin, Serena Pastore, Andrea Taddio, Luciana Rodrigues Roberti, Rosane Gomes de Paula Queiroz, Virginia Paes Leme Ferriani, Sergio Crovella, and Alberto Tommasini. Higher interferon score and normal complement levels may identify a distinct clinical subset in children with systemic lupus erythematosus. Arthritis Research & Therapy, 22(1):91, 2020. ISSN 1478-6362. doi: 10.1186/s13075-020-02161-8. URL https://doi.org/10.1186/s13075-020-02161-8.

[86] François Chasset, Camillo Ribi, Marten Trendelenburg, Uyen Huynh-Do, Pascale Roux-Lombard, Delphine S Courvoisier, Carlo Chizzolini, and for the Swiss SLE Cohort Study (SSCS. Identification of highly active systemic lupus erythematosus by combined type I interferon and neutrophil gene scores vs classical serologic markers. Rheumatology, 59(11):3468–3478, 05 2020. ISSN 1462-0324. doi: 10.1093/rheumatology/keaa167. URL https://doi.org/10.1093/rheumatology/keaa167.

[87] Su-Jin Moon, Jung Min Bae, Kyung-Su Park, Ilias Tagkopoulos, and Ki-Jo Kim. Compendium of skin molecular signatures identifies key pathological features associated with fibrosis in systemic sclerosis. Annals of the Rheumatic Diseases, 78(6):817–825, 2019. ISSN 0003-4967. doi: 10.1136/annrheumdis-2018-214778. URL https://ard.bmj.com/content/78/6/817.

[88] Elham Rezaei, Daniel Hogan, Brett Trost, Anthony J Kusalik, Gilles Boire, David A Cabral, Sarah Campillo, Gaëlle Chédeville, Anne-Laure Chetaille, Paul Dancey, Ciaran Duffy, Karen Watanabe Duffy, Simon W M Eng, John Gordon, Jaime Guzman, Kristin Houghton, Adam M Huber, Roman Jurencak, Bianca Lang, Ronald M Laxer, Kimberly Morishita, Kiem G Oen, Ross E Petty, Suzanne E Ramsey, Stephen W Scherer, Rosie Scuccimarri, Lynn Spiegel, Elizabeth Stringer, Regina M Taylor-Gjevre, Shirley M L Tse, Lori B Tucker, Stuart E Turvey, Susan Tupper, Richard F Wintle, Rae S M Yeung, Alan M Rosenberg, and for the BBOP Study Group. Associations of clinical and inflammatory biomarker clusters with juvenile idiopathic arthritis categories. Rheumatology, 59(5):1066–1075, 09 2019. ISSN 1462-0324. doi: 10.1093/rheumatology/kez382. URL https://doi.org/10.1093/rheumatology/kez382.

[89] Rodrigo Cánovas, Joanna Cobb, Marta Brozynska, John Bowes, Yun R Li, Samantha Louise Smith, Hakon Hakonarson, Wendy Thomson, Justine A Ellis, Gad Abraham, Jane E Munro, and Michael Inouye. Genomic risk scores for juvenile idiopathic arthritis and its subtypes. Annals of the Rheumatic Diseases, 79(12):1572–1579, 2020. ISSN 0003-4967. doi: 10.1136/annrheumdis-2020-217421. URL https://ard.bmj.com/content/79/12/1572.

[90] Kerry E. Poppenberg, Kaiyu Jiang, Lu Li, Yijun Sun, Hui Meng, Carol A. Wallace, Teresa Hennon, and James N. Jarvis. The feasibility of developing biomarkers from peripheral blood mononuclear cell RNAseq data in children with juvenile idiopathic arthritis using machine learning approaches. Arthritis Research & Therapy, 21(1):230, 12 2019. ISSN 1478-6362. doi: 10.1186/s13075-019-2010-z. URL https://arthritis-research.biomedcentral.com/articles/10.1186/s13075-019-2010-z.

[91] Dana E. Orange, Phaedra Agius, Edward F. DiCarlo, Nicolas Robine, Heather Geiger, Jackie Szymonifka, Michael McNamara, Ryan Cummings, Kathleen M. Andersen, Serene Mirza, Mark Figgie, Lionel B. Ivashkiv, Alessandra B. Pernis, Caroline S. Jiang, Mayu O. Frank, Robert B. Darnell, Nithya Lingampali, William H. Robinson, Ellen Gravallese, the Accelerating Medicines Partnership in Rheumatoid Arthritis and Lupus Network, Vivian P. Bykerk, Susan M. Goodman, and Laura T. Donlin. Identification of three rheumatoid arthritis disease subtypes by machine learning integration of synovial histologic features and rna sequencing data. Arthritis & Rheumatology, 70(5): 690–701, 2018. doi: https://doi.org/10.1002/art.40428. URL https://onlinelibrary.wiley.com/doi/abs/10.1002/art.40428.

[92] Daniel J. Kass, Mehdi Nouraie, Marilyn K. Glassberg, Nitya Ramreddy, Karen Fernandez, Lisa Harlow, Yingze Zhang, Jean Chen, Gail S. Kerr, Andreas M. Reimold, Bryant R. England, Ted R. Mikuls, Kevin F. Gibson, Paul F. Dellaripa, Ivan O. Rosas, Chester V. Oddis, and Dana P. Ascherman. Comparative profiling of serum protein biomarkers in rheumatoid arthritis–associated interstitial lung disease and idiopathic pulmonary fibrosis. Arthritis & Rheumatology, 72(3):409–419, 2020. doi: https://doi.org/10.1002/art.41123. URL https://onlinelibrary.wiley.com/doi/abs/10.1002/art.41123.

[93] K. Bates Gribbons, Cristina Ponte, Simon Carette, Anthea Craven, David Cuthbertson, Gary S. Hoffman, Nader A. Khalidi, Curry L. Koening, Carol A. Langford, Kathleen Maksimowicz-McKinnon, Carol A. McAlear, Paul A. Monach, Larry W. Moreland, Christian Pagnoux, Kaitlin A. Quinn, Joanna C. Robson, Philip Seo, Antoine G. Sreih, Ravi Suppiah, Kenneth J. Warrington, Steven R. Ytterberg, Raashid Luqmani, Richard Watts, Peter A. Merkel, and Peter C. Grayson. Patterns of arterial disease in takayasu arteritis and giant cell arteritis. Arthritis Care & Research, 72(11):1615–1624, 2020. doi: https://doi.org/10.1002/acr.24055. URL https://onlinelibrary.wiley.com/doi/abs/10.1002/acr.24055.

[94] Ruchika Goel, K Bates Gribbons, Simon Carette, David Cuthbertson, Gary S Hoffman, George Joseph, Nader A Khalidi, Curry L Koening, Sathish Kumar, Carol Langford, Kathleen Maksimowicz-McKinnon, Carol A McAlear, Paul A Monach, Larry W Moreland, Aswin Nair, Christian Pagnoux, Kaitlin A Quinn, Raheesh Ravindran, Philip Seo, Antoine G Sreih, Kenneth J Warrington, Steven R Ytterberg, Peter A Merkel, Debashish Danda, and Peter C Grayson. Derivation of an angiographically based classification system in Takayasu’s arteritis: an observational study from India and North America. Rheumatology, 59(5):1118–1127, 10 2019. ISSN 1462-0324. doi: 10.1093/rheumatology/kez421. URL https://doi.org/10.1093/rheumatology/kez421.

[95] Jakob Kristian Holm Andersen, Jannik Skyttegaard Pedersen, Martin Sundahl Laursen, Kathrine Holtz, Jakob Grauslund, Thiusius Rajeeth Savarimuthu, and Søren Andreas Just. Neural networks for automatic scoring of arthritis disease activity on ultrasound images. RMD Open, 5(1), 2019. doi: 10.1136/rmdopen-2018-000891. URL https://rmdopen.bmj.com/content/5/1/e000891.

[96] Anders Bossel Holst Christensen, Søren Andreas Just, Jakob Kristian Holm Andersen, and Thiusius Rajeeth Savarimuthu. Applying cascaded convolutional neural network design further enhances automatic scoring of arthritis disease activity on ultrasound images from rheumatoid arthritis patients. Annals of the Rheumatic Diseases, 79(9):1189–1193, 2020. ISSN 0003-4967. doi: 10.1136/annrheumdis-2019-216636. URL https://ard.bmj.com/content/79/9/1189.

[97] Farhad Akhbardeh, Fartash Vasefi, Nick MacKinnon, Mohammad Amini, Alireza Akhbardeh, and Kouhyar Tavakolian. Classification and assessment of hand arthritis stage using support vector machine. In 2019 41st Annual International Conference of the IEEE Engineering in Medicine and Biology Society (EMBC), pages 4080–4083, 2019. doi: 10.1109/EMBC.2019.8857022.

[98] Vincenzo Venerito, Orazio Angelini, Gerardo Cazzato, Giuseppe Lopalco, Eugenio Maiorano, Antonietta Cimmino, and Florenzo Iannone. A convolutional neural network with transfer learning for automatic discrimination between low and high-grade synovitis: a pilot study. Internal and Emergency Medicine, 16:1457–1465, 2021. ISSN 1970-9366. doi: 10.1007/s11739-020-02583-x. URL https://doi.org/10.1007/s11739-020-02583-x.

[99] Alberta Hoi, Hieu T Nim, Rachel Koelmeyer, Ying Sun, Amy Kao, Oliver Gunther, and Eric Morand. Algorithm for calculating high disease activity in SLE. Rheumatology, 60(9):4291–4297, 01 2021. ISSN 1462-0324. doi: 10.1093/rheumatology/keab003. URL https://doi.org/10.1093/rheumatology/keab003.

[100] Kanon Jatuworapruk, Rebecca Grainger, Nicola Dalbeth, and William J. Taylor. Development of a prediction model for inpatient gout flares in people with comorbid gout. Annals of the Rheumatic Diseases, 79(3):418–423, 2020. ISSN 0003-4967. doi: 10.1136/annrheumdis-2019-216277. URL https://ard.bmj.com/content/79/3/418.

[101] Mukundan Attur, Svetlana Krasnokutsky, Hua Zhou, Jonathan Samuels, Gregory Chang, Jenny Bencardino, Pamela Rosenthal, Leon Rybak, Janet L Huebner, Virginia B Kraus, and Steven B Abramson. The combination of an inflammatory peripheral blood gene expression and imaging biomarkers enhance prediction of radiographic progression in knee osteoarthritis. Arthritis Research & Therapy, 22(1):208, 2020. ISSN 1478-6362. doi: 10.1186/s13075-020-02298-6. URL https://doi.org/10.1186/s13075-020-02298-6.

[102] Zhaoye Zhou, Gengyan Zhao, Richard Kijowski, and Fang Liu. Deep convolutional neural network for segmentation of knee joint anatomy. Magnetic Resonance in Medicine, 80(6):2759–2770, 2018. doi: https://doi.org/10.1002/mrm.27229. URL https://onlinelibrary.wiley.com/doi/abs/10.1002/mrm.27229.

[103] Felix Eckstein, Akshay S. Chaudhari, David Fuerst, Martin Gaisberger, Jana Kemnitz, Christian F. Baumgartner, Ender Konukoglu, David J Hunter, and Wolfgang Wirth. A deep learning automated segmentation algorithm accurately detects differences in longitudinal cartilage thickness loss – data from the fnih biomarkers study of the osteoarthritis initiative. Arthritis Care & Research, 74(6):929–936, 2022. doi: https://doi.org/10.1002/acr.24539. URL https://onlinelibrary.wiley.com/doi/abs/10.1002/acr.24539.

[104] Sibaji Gaj, Mingrui Yang, Kunio Nakamura, and Xiaojuan Li. Automated cartilage and meniscus segmentation of knee mri with conditional generative adversarial networks. Magnetic Resonance in Medicine, 84(1):437–449, 2020. doi: https://doi.org/10.1002/mrm.28111. URL https://onlinelibrary.wiley.com/doi/abs/10.1002/mrm.28111.

[105] Ruida Cheng, Natalia A. Alexandridi, Richard M. Smith, Aricia Shen, William Gandler, Evan McCreedy, Matthew J. McAuliffe, and Frances T. Sheehan. Fully automated patellofemoral mri segmentation using holistically nested networks: Implications for evaluating patellofemoral osteoarthritis, pain, injury, pathology, and adolescent development. Magnetic Resonance in Medicine, 83(1):139–153, 2020. doi: https://doi.org/10.1002/mrm.27920. URL https://onlinelibrary.wiley.com/doi/abs/10.1002/mrm.27920.

[106] Seulkee Lee, Yeonghee Eun, Hyungjin Kim, Hoon-Suk Cha, Eun-Mi Koh, and Jaejoon Lee. Machine learning to predict early TNF inhibitor users in patients with ankylosing spondylitis. Scientific Reports, 10(1):20299, 2020. ISSN 2045-2322. doi: 10.1038/s41598-020-75352-7. URL https://doi.org/10.1038/s41598-020-75352-7.

[107] Tianrun Cai, Tzu-Chieh Lin, Allison Bond, Jie Huang, Gwendolyn Kane-Wanger, Andrew Cagan, Shawn N Murphy, Ashwin N Ananthakrishnan, and Katherine P Liao. The Association Between Arthralgia and Vedolizumab Using Natural Language Processing. Inflammatory Bowel Diseases, 24(10):2242–2246, 05 2018. ISSN 1078-0998. doi: 10.1093/ibd/izy127. URL https://doi.org/10.1093/ibd/izy127.

[108] Jeffrey R. Curtis, Lang Chen, Phillip Higginbotham, W. Benjamin Nowell, Ronit Gal-Levy, James Willig, Monika Safford, Joseph Coe, Kaitlin O’Hara, and Roee Sa’adon. Social media for arthritis-related comparative effectiveness and safety research and the impact of direct-to-consumer advertising. Arthritis Research & Therapy, 19(1):48, 12 2017. ISSN 1478-6362. doi: 10.1186/s13075-017-1251-y. URL http://arthritis-research.biomedcentral.com/articles/10.1186/s13075-017-1251-y.

[109] Eldin Dzubur, Carine Khalil, Christopher V. Almario, Benjamin Noah, Deeba Minhas, Mariko Ishimori, Corey Arnold, Yujin Park, Jonathan Kay, Michael H. Weisman, and Brennan M. R. Spiegel. Patient concerns and perceptions regarding biologic therapies in ankylosing spondylitis: Insights from a large-scale survey of social media platforms. Arthritis Care & Research, 71(2):323–330, 2019. doi: https://doi.org/10.1002/acr.23600. URL https://onlinelibrary.wiley.com/doi/abs/10.1002/acr.23600.

[110] Chanakya Sharma, Samuel Whittle, Pari Delir Haghighi, Frada Burstein, Roee Sa’adon, and Helen Isobel Keen. Mining social media data to investigate patient perceptions regarding dmard pharmacotherapy for rheumatoid arthritis. Annals of the Rheumatic Diseases, 79(11): 1432–1437, 2020. ISSN 0003-4967. doi: 10.1136/annrheumdis-2020-217333. URL https://ard.bmj.com/content/79/11/1432.

[111] Xiaolan Mo, Xiujuan Chen, Hongwei Li, Jiali Li, Fangling Zeng, Yilu Chen, Fan He, Song Zhang, Huixian Li, Liyan Pan, Ping Zeng, Ying Xie, Huiyi Li, Min Huang, Yanling He, Huiying Liang, and Huasong Zeng. Early and accurate prediction of clinical response to methotrexate treatment in juvenile idiopathic arthritis using machine learning. Frontiers in Pharmacology, 10:1155, 2019. ISSN 1663-9812. doi: 10.3389/fphar.2019.01155. URL https://www.frontiersin.org/article/10.3389/fphar.2019.01155.

[112] Xiaolan Mo, Xiujuan Chen, Chifong Ieong, Song Zhang, Huiyi Li, Jiali Li, Guohao Lin, Guangchao Sun, Fan He, Yanling He, Ying Xie, Ping Zeng, Yilu Chen, Huiying Liang, and Huasong Zeng. Early prediction of clinical response to etanercept treatment in juvenile idiopathic arthritis using machine learning. Frontiers in Pharmacology, 11:1164, 2020. ISSN 1663-9812. doi: 10.3389/fphar.2020.01164. URL https://www.frontiersin.org/article/10.3389/fphar.2020.01164.

[113] Liangliang Liu, Ying Yu, Zhihui Fei, Min Li, Fang-Xiang Wu, Hong-Dong Li, Yi Pan, and Jianxin Wang. An interpretable boosting model to predict side effects of analgesics for osteoarthritis. BMC Systems Biology, 12(S6):105, 11 2018. ISSN 1752-0509. doi: 10.1186/s12918-018-0624-4. URL https://bmcsystbiol.biomedcentral.com/articles/10.1186/s12918-018-0624-4.

[114] Alice B Gottlieb, Philip J Mease, Bruce Kirkham, Peter Nash, Alejandro C Balsa, Bernard Combe, Jürgen Rech, Xuan Zhu, David James, Ruvie Martin, Gregory Ligozio, Ken Abrams, and Luminita Pricop. Secukinumab Efficacy in Psoriatic Arthritis: Machine Learning and Meta-analysis of Four Phase 3 Trials. JCR: Journal of Clinical Rheumatology, 27(6), 2021. ISSN 1076-1608. URL https://journals.lww.com/jclinrheum/Fulltext/2021/09000/Secukinumab_Efficacy_in_Psoriatic_Arthritis_.4.aspx.

[115] Tatsuya Atsumi, Yoshiaki Ando, Shinichi Matsuda, Shiho Tomizawa, Riwa Tanaka, Nobuhiro Takagi, and Ayako Nakasone. Prodromal signs and symptoms of serious infections with tocilizumab treatment for rheumatoid arthritis: Text mining of the japanese postmarketing adverse event-reporting database. Modern Rheumatology, 28(3):435–443, 2018. doi: 10.1080/14397595.2017.1366007. URL https://doi.org/10.1080/14397595.2017.1366007. PMID: 28880689.

[116] Tengyang Wang, Guanghua Liu, and Hongye Lin. A machine learning approach to predict intravenous immunoglobulin resistance in kawasaki disease patients: A study based on a southeast china population. PLOS ONE, 15(8):1–15, 08 2020. doi: 10.1371/journal.pone.0237321. URL https://doi.org/10.1371/journal.pone.0237321.

[117] Mashfiqui Rabbi, Min SH Aung, Geri Gay, M Cary Reid, and Tanzeem Choudhury. Feasibility and Acceptability of Mobile Phone–Based Auto-Personalized Physical Activity Recommendations for Chronic Pain Self-Management: Pilot Study on Adults. Journal of Medical Internet Research, 20(10):e10147, 10 2018. ISSN 1438-8871. doi: 10.2196/10147. URL http://www.jmir.org/2018/10/e10147/.

[118] Athina Spiliopoulou, Marco Colombo, Darren Plant, Nisha Nair, Jing Cui, Marieke JH Coenen, Katsunori Ikari, Hisashi Yamanaka, Saedis Saevarsdottir, Leonid Padyukov, S Louis Bridges Jr., Robert P Kimberly, Yukinori Okada, Piet L CM van Riel, Gertjan Wolbink, Irene E van der Horst-Bruinsma, Niek de Vries, Paul P Tak, Koichiro Ohmura, Helena Canhão, Henk-Jan Guchelaar, Tom WJ Huizinga, Lindsey A Criswell, Soumya Raychaudhuri, Michael E Weinblatt, Anthony G Wilson, Xavier Mariette, John D Isaacs, Ann W Morgan, Costantino Pitzalis, Anne Barton, and Paul McKeigue. Association of response to tnf inhibitors in rheumatoid arthritis with quantitative trait loci for cd40 and cd39. Annals of the Rheumatic Diseases, 78(8):1055–1061, 2019. ISSN 0003-4967. doi: 10.1136/annrheumdis-2018-214877. URL https://ard.bmj.com/content/78/8/1055.

[119] Yuanfang Guan, Hongjiu Zhang, Daniel Quang, Ziyan Wang, Stephen C. J. Parker, Dimitrios A. Pappas, Joel M. Kremer, and Fan Zhu. Machine learning to predict anti–tumor necrosis factor drug responses of rheumatoid arthritis patients by integrating clinical and genetic markers. Arthritis & Rheumatology, 71(12):1987–1996, 2019. doi: https://doi.org/10.1002/art.41056. URL https://onlinelibrary.wiley.com/doi/abs/10.1002/art.41056.

[120] Weiyang Tao, Arno N. Concepcion, Marieke Vianen, Anne C. A. Marijnissen, Floris P. G. J. Lafeber, Timothy R. D. J. Radstake, and Aridaman Pandit. Multiomics and machine learning accurately predict clinical response to adalimumab and etanercept therapy in patients with rheumatoid arthritis. Arthritis & Rheumatology, 73(2):212–222, 2021. doi: https://doi.org/10.1002/art.41516. URL https://onlinelibrary.wiley.com/doi/abs/10.1002/art.41516.

[121] Darren Plant, Mateusz Maciejewski, Samantha Smith, Nisha Nair, the RAMS Study Group the Maximising Therapeutic Utility in Rheumatoid Arthritis Consortium, Kimme Hyrich, Daniel Ziemek, Anne Barton, and Suzanne Verstappen. Profiling of gene expression biomarkers as a classifier of methotrexate nonresponse in patients with rheumatoid arthritis. Arthritis & Rheumatology, 71(5):678–684, 2019. doi: https://doi.org/10.1002/art.40810. URL https://onlinelibrary.wiley.com/doi/abs/10.1002/art.40810.

[122] Helen R. Gosselt, Maxime M. A. Verhoeven, Maja Bulatović-Ćalasan, Paco M. Welsing, Maurits C. F. J. de Rotte, Johanna M. W. Hazes, Floris P. J. G. Lafeber, Mark Hoogendoorn, and Robert de Jonge. Complex machine-learning algorithms and multivariable logistic regression on par in the prediction of insufficient clinical response to methotrexate in rheumatoid arthritis. Journal of Personalized Medicine, 11(1), 2021. ISSN 2075-4426. doi: 10.3390/jpm11010044. URL https://www.mdpi.com/2075-4426/11/1/44.

[123] Frances Humby, Myles Lewis, Nandhini Ramamoorthi, Jason A Hackney, Michael R Barnes, Michele Bombardieri, A. Francesca Setiadi, Stephen Kelly, Fabiola Bene, Maria DiCicco, Sudeh Riahi, Vidalba Rocher, Nora Ng, Ilias Lazarou, Rebecca Hands, Désirée van der Heijde, Robert B M Landewé, Annette van der Helm-van Mil, Alberto Cauli, Iain McInnes, Christopher Dominic Buckley, Ernest H Choy, Peter C Taylor, Michael J Townsend, and Costantino Pitzalis. Synovial cellular and molecular signatures stratify clinical response to csdmard therapy and predict radiographic progression in early rheumatoid arthritis patients. Annals of the Rheumatic Diseases, 78(6):761–772, 2019. ISSN 0003-4967. doi: 10.1136/annrheumdis-2018-214539. URL https://ard.bmj.com/content/78/6/761.

[124] Jinchao Jia, Mengyan Wang, Yuning Ma, Jialin Teng, Hui Shi, Honglei Liu, Yue Sun, Yutong Su, Jianfen Meng, Huihui Chi, Xia Chen, Xiaobing Cheng, Junna Ye, Tingting Liu, Zhihong Wang, Liyan Wan, Zhuochao Zhou, Fan Wang, Chengde Yang, and Qiongyi Hu. Circulating neutrophil extracellular traps signature for identifying organ involvement and response to glucocorticoid in adult-onset still’s disease: A machine learning study. Frontiers in Immunology, 11:2784, 2020. ISSN 1664-3224. doi: 10.3389/fimmu.2020.563335. URL https://www.frontiersin.org/article/10.3389/fimmu.2020.563335.

[125] Chandrika S Bhat, Mark Chopra, Savvas Andronikou, Suvadip Paul, Zach Wener-Fligner, Anna Merkoulovitch, Izidora Holjar-Erlic, Flavia Menegotto, Ewan Simpson, David Grier, and Athimalaipet V Ramanan. Artificial intelligence for interpretation of segments of whole body MRI in CNO: pilot study comparing radiologists versus machine learning algorithm. Pediatric Rheumatology, 18(1):47, 2020. ISSN 1546-0096. doi: 10.1186/s12969-020-00442-9. URL https://doi.org/10.1186/s12969-020-00442-9.

[126] Man Hung, Jerry Bounsanga, Fangzhou Liu, and Maren W. Voss. Profiling arthritis pain with a decision tree. Pain Practice, 18(5):568–579, 2018. doi: https://doi.org/10.1111/papr.12645. URL https://onlinelibrary.wiley.com/doi/abs/10.1111/papr.12645.

[127] Mike Becker, Nicole Graf, Rafael Sauter, Yannick Allanore, John Curram, Christopher P Denton, Dinesh Khanna, Marco Matucci-Cerinic, Janethe de Oliveira Pena, Janet E Pope, and Oliver Distler. Predictors of disease worsening defined by progression of organ damage in diffuse systemic sclerosis: a european scleroderma trials and research (eustar) analysis. Annals of the Rheumatic Diseases, 78(9):1242–1248, 2019. ISSN 0003-4967. doi: 10.1136/annrheumdis-2019-215145. URL https://ard.bmj.com/content/78/9/1242.

[128] Haytham Eloqayli, Ali Al-Yousef, and Raid Jaradat. Vitamin d and ferritin correlation with chronic neck pain using standard statistics and a novel artificial neural network prediction model. British Journal of Neurosurgery, 32(2):172–176, 2018. doi: 10.1080/02688697.2018.1436691. URL https://doi.org/10.1080/02688697.2018.1436691. PMID: 29447493.

[129] Megan E. Breitbach, Ryne C. Ramaker, Kevin Roberts, Robert P. Kimberly, and Devin Absher. Population-specific patterns of epigenetic defects in the b cell lineage in patients with systemic lupus erythematosus. Arthritis & Rheumatology, 72(2):282–291, 2020. doi: https://doi.org/10.1002/art.41083. URL https://onlinelibrary.wiley.com/doi/abs/10.1002/art.41083.

[130] Kamala Vanarsa, Sanam Soomro, Ting Zhang, Briony Strachan, Claudia Pedroza, Malavika Nidhi, Pietro Cicalese, Christopher Gidley, Shobha Dasari, Shree Mohan, Nathan Thai, Van Thi Thanh Truong, Nicole Jordan, Ramesh Saxena, Chaim Putterman, Michelle Petri, and Chandra Mohan. Quantitative planar array screen of 1000 proteins uncovers novel urinary protein biomarkers of lupus nephritis. Annals of the Rheumatic Diseases, 79(10):1349–1361, 2020. ISSN 0003-4967. doi: 10.1136/annrheumdis-2019-216312. URL https://ard.bmj.com/content/79/10/1349.

[131] M Paula Gomez Hernandez, Emily E Starman, Andrew B Davis, Miyuraj Harishchandra Hikkaduwa Withanage, Erliang Zeng, Scott M Lieberman, Kim A Brogden, and Emily A Lanzel. A distinguishing profile of chemokines, cytokines and biomarkers in the saliva of children with Sjögren’s syndrome. Rheumatology, 01 2021. ISSN 1462-0324. doi: 10.1093/rheumatology/keab098. URL https://doi.org/10.1093/rheumatology/keab098.keab098.

[132] Parisa Riahi, Anoshirvan Kazemnejad, Shayan Mostafaei, Akira Meguro, Nobuhisa Mizuki, Amir Ashraf-Ganjouei, Ali Javinani, Seyedeh Tahereh Faezi, Farhad Shahram, and Mahdi Mahmoudi. Erap1 polymorphisms interactions and their association with behçet’s disease susceptibly: Application of model-based multifactor dimension reduction algorithm (mb-mdr). PLOS ONE, 15(2):1–10, 02 2020. doi: 10.1371/journal.pone.0227997. URL https://doi.org/10.1371/journal.pone.0227997.

[133] Masaki Asakage, Yoshihiko Usui, Naoya Nezu, Hiroyuki Shimizu, Kinya Tsubota, Naoyuki Yamakawa, Masakatsu Takanashi, Masahiko Kuroda, and Hiroshi Goto. Comprehensive miRNA Analysis Using Serum From Patients With Noninfectious Uveitis. Investigative Ophthalmology & Visual Science, 61(11):4–4, 09 2020. ISSN 1552-5783. doi: 10.1167/iovs.61.11.4. URL https://doi.org/10.1167/iovs.61.11.4.

[134] Laura Andrés-Rodríguez, Xavier Borràs, Albert Feliu-Soler, Adrián Pérez-Aranda, Antoni Rozadilla-Sacanell, Belén Arranz, Jesús Montero-Marin, Javier García-Campayo, Natalia Angarita-Osorio, Michael Maes, and Juan V. Luciano. Machine learning to understand the immune-inflammatory pathways in fibromyalgia. International Journal of Molecular Sciences, 20(17), 2019. ISSN 1422-0067. doi: 10.3390/ijms20174231. URL https://www.mdpi.com/1422-0067/20/17/4231.

[135] Alicia Rodriguez-Pla, Roscoe L. Warner, David Cuthbertson, Simon Carette, Nader A. Khalidi, Curry L. Koening, Carol A. Langford, Carol A. McAlear, Larry W. Moreland, Christian Pagnoux, Philip Seo, Ulrich Specks, Antoine G. Sreih, Steven R. Ytterberg, Kent J. Johnson, Peter A. Merkel, and Paul A. Monach. Evaluation of potential serum biomarkers of disease activity in diverse forms of vasculitis. The Journal of Rheumatology, 2019. ISSN 0315-162X. doi: 10.3899/jrheum.190093. URL https://www.jrheum.org/content/early/2020/01/27/jrheum.190093.

[136] Axel Finckh, Benoît Gilbert, Bridget Hodkinson, Sang-Cheol Bae, Ranjeny Thomas, Kevin D. Deane, Deshiré Alpizar-Rodriguez, and Kim Lauper. Global epidemiology of rheumatoid arthritis. Nature Reviews Rheumatology, 9 2022. ISSN 1759-4790. doi: 10.1038/s41584-022-00827-y.

[137] David J Hunter and Sita Bierma-Zeinstra. Osteoarthritis. The Lancet, 393(10182):1745–1759, 2019. ISSN 0140-6736. doi: https://doi.org/10.1016/S0140-6736(19)30417-9. URL https://www.sciencedirect.com/science/article/pii/S0140673619304179.

[138] Nihan Acar-Denizli, Belchin Kostov, Manuel Ramos-Casals, and Sjögren Big Data Consortium. The big data sjögren consortium: a project for a new data science era. Clinical and experimental rheumatology, 37 Suppl 118:19–23, 2019. ISSN 0392-856X.

[139] Aurélien Géron. Hands-on Machine Learning with Scikit-Learn, Keras, and TensorFlow: Concepts, Tools, and Techniques to Build Intelligent Systems (3rd ed.). O’Reilly Media, Sebastopol, CA, 2022. ISBN 978-1492032649.

[140] Joseph Futoma, Morgan Simons, Trishan Panch, Finale Doshi-Velez, and Leo Anthony Celi. The myth of generalisability in clinical research and machine learning in health care. The Lancet Digital Health, 2:e489–e492, 9 2020. ISSN 25897500. doi: 10.1016/S2589-7500(20)30186-2.

[141] Hun-Sung Kim, Suehyun Lee, and Ju Han Kim. Real-world evidence versus randomized controlled trial: Clinical research based on electronic medical records. Journal of Korean Medical Science, 33, 2018. ISSN 1011-8934. doi: 10.3346/jkms.2018.33.e213.

[142] Jyothi Subramanian and Richard Simon. Overfitting in prediction models – is it a problem only in high dimensions? Contemporary Clinical Trials, 36(2):636–641, 2013. ISSN 1551-7144. doi: https://doi.org/10.1016/j.cct.2013.06.011. URL https://www.sciencedirect.com/science/article/pii/S1551714413001031.

[143] Irene Y. Chen, Emma Pierson, Sherri Rose, Shalmali Joshi, Kadija Ferryman, and Marzyeh Ghassemi. Ethical machine learning in healthcare. Annual Review of Biomedical Data Science, 4(1):123–144, 2021. doi: 10.1146/annurev-biodatasci-092820-114757. URL https://doi.org/10.1146/annurev-biodatasci-092820-114757.

[144] Kevin Yip and Iris Navarro-Millán. Racial, ethnic, and healthcare disparities in rheumatoid arthritis. Current Opinion in Rheumatology, 33, 2021. ISSN 1040-8711. URL https://journals.lww.com/co-rheumatology/Fulltext/2021/03000/Racial,_ethnic,_and_healthcare_disparities_in.3.aspx.

[145] Warachal E. Faison, P. Grace Harrell, and David Semel. Disparities across diverse populations in the health and treatment of patients with osteoarthritis. Healthcare, 9(11), 2021. ISSN 2227-9032. URL https://www.mdpi.com/2227-9032/9/11/1421.

[146] Jasvinder A. Singh. Racial and gender disparities among patients with gout. Current Rheumatology Reports, 15:307, 2 2013. ISSN 1523-3774. doi: 10.1007/s11926-012-0307-x.

[147] Maria Dall’Era, Miriam G. Cisternas, Kurt Snipes, Lisa J. Herrinton, Caroline Gordon, and Charles G. Helmick. The incidence and prevalence of systemic lupus erythematosus in san francisco county, california: The california lupus surveillance project. Arthritis & Rheumatology, 69 (10):1996–2005, 2017. doi: https://doi.org/10.1002/art.40191. URL https://onlinelibrary.wiley.com/doi/abs/10.1002/art.40191.

[148] Milena A Gianfrancesco, Maria Dall’Era, Louise B Murphy, Charles G Helmick, Jing Li, Stephanie Rush, Laura Trupin, and Jinoos Yazdany. Mortality among minority populations with systemic lupus erythematosus, including asian and hispanic/latino persons—california, 2007–2017. Morbidity and Mortality Weekly Report, 70(7):236, 2021.

[149] Gracie Himmelstein, David Bates, and Li Zhou. Examination of Stigmatizing Language in the Electronic Health Record. JAMA Network Open, 5(1):e2144967–e2144967, 01 2022. ISSN 2574-3805. doi: 10.1001/jamanetworkopen.2021.44967. URL https://doi.org/10.1001/jamanetworkopen.2021.44967.

[150] Daniel Schönberger. Artificial intelligence in healthcare: a critical analysis of the legal and ethical implications. International Journal of Law and Information Technology, 27(2):171–203, 05 2019. ISSN 0967-0769. doi: 10.1093/ijlit/eaz004. URL https://doi.org/10.1093/ijlit/eaz004.

[151] Christopher J. Kelly, Alan Karthikesalingam, Mustafa Suleyman, Greg Corrado, and Dominic King. Key challenges for delivering clinical impact with artificial intelligence. BMC Medicine, 17(1):195, 12 2019. ISSN 1741-7015. doi: 10.1186/s12916-019-1426-2. URL https://bmcmedicine.biomedcentral.com/articles/10.1186/s12916-019-1426-2.

[152] Vincenzo Lagani, George Kortas, and Ioannis Tsamardinos. Biomarker signature identification in “omics” data with multi-class outcome. Computational and Structural Biotechnology Journal, 6(7):e201303004, 2013. ISSN 2001-0370. doi: https://doi.org/10.5936/csbj.201303004. URL https://www.sciencedirect.com/science/article/pii/S2001037014601136.

[153] Marzyeh Ghassemi, Luke Oakden-Rayner, and Andrew L Beam. The false hope of current approaches to explainable artificial intelligence in health care. The Lancet Digital Health, 3:e745–e750, 11 2021. ISSN 25897500. doi: 10.1016/S2589-7500(21)00208-9.

[154] Sandeep Reddy. Explainability and artificial intelligence in medicine. The Lancet Digital Health, 4:e214–e215, 4 2022. ISSN 25897500. doi: 10.1016/S2589-7500(22)00029-2.

[155] Natalia Norori, Qiyang Hu, Florence Marcelle Aellen, Francesca Dalia Faraci, and Athina Tzovara. Addressing bias in big data and ai for health care: A call for open science. Patterns, 2(10):100347, 2021. ISSN 2666-3899. doi: https://doi.org/10.1016/j.patter.2021.100347. URL https://www.sciencedirect.com/science/article/pii/S2666389921002026.

[156] Gómez-González E and Gomez Gutierrez E. Artificial intelligence in medicine and healthcare: applications, availability and societal impact. Scientific analysis or review KJ-NA-30197-EN-N (online), European Union, Luxembourg (Luxembourg), 2020.

[157] European Comission. Regulatory framework proposal on artificial intelligence, 2022. URL https://digital-strategy.ec.europa.eu/en/policies/regulatory-framework-ai.

[158] European Commission, Content Directorate-General for Communications Networks, and Technology. Ethics guidelines for trustworthy AI. Publications Office, 2019. doi: doi/10.2759/346720.

[159] Roberto V. Zicari, James Brusseau, Stig Nikolaj Blomberg, Helle Collatz Christensen, Megan Coffee, Marianna B. Ganapini, Sara Gerke, Thomas Krendl Gilbert, Eleanore Hickman, Elisabeth Hildt, Sune Holm, Ulrich Kühne, Vince I. Madai, Walter Osika, Andy Spezzatti, Eberhard Schnebel, Jesmin Jahan Tithi, Dennis Vetter, Magnus Westerlund, Renee Wurth, Julia Amann, Vegard Antun, Valentina Beretta, Frédérick Bruneault, Erik Campano, Boris Düdder, Alessio Gallucci, Emmanuel Goffi, Christoffer Bjerre Haase, Thilo Hagendorff, Pedro Kringen, Florian Möslein, Davi Ottenheimer, Matiss Ozols, Laura Palazzani, Martin Petrin, Karin Tafur, Jim Tørresen, Holger Volland, and Georgios Kararigas. On assessing trustworthy ai in healthcare. machine learning as a supportive tool to recognize cardiac arrest in emergency calls. Frontiers in Human Dynamics, 3, 2021. ISSN 2673-2726. doi: 10.3389/fhumd.2021.673104. URL https://www.frontiersin.org/articles/10.3389/fhumd.2021.673104.

[160] Media Secretary of State for Digital, Culture and Sport. Establishing a pro-innovation approach to regulating ai, 2022. URL https://www.gov.uk/government/publications/establishing-a-pro-innovation-approach-to-regulating-ai.

[161] Gary S Collins, Paula Dhiman, Constanza L Andaur Navarro, Jie Ma, Lotty Hooft, Johannes B Reitsma, Patricia Logullo, Andrew L Beam, Lily Peng, Ben Van Calster, Maarten van Smeden, Richard D Riley, and Karel GM Moons. Protocol for development of a reporting guideline (tripod-ai) and risk of bias tool (probast-ai) for diagnostic and prognostic prediction model studies based on artificial intelligence. BMJ Open, 11(7), 2021. ISSN 2044-6055. doi: 10.1136/bmjopen-2020-048008. URL https://bmjopen.bmj.com/content/11/7/e048008.

[162] Samantha Cruz Rivera, Xiaoxuan Liu, An-Wen Chan, Alastair K Denniston, and Melanie J Calvert. Guidelines for clinical trial protocols for interventions involving artificial intelligence: the spirit-ai extension. BMJ, 370, 2020. doi: 10.1136/bmj.m3210. URL https://www.bmj.com/content/370/bmj.m3210.

[163] Xiaoxuan Liu, Samantha Cruz Rivera, David Moher, Melanie J Calvert, and Alastair K Denniston. Reporting guidelines for clinical trial reports for interventions involving artificial intelligence: the consort-ai extension. BMJ, 370, 2020. doi: 10.1136/bmj.m3164. URL https://www.bmj.com/content/370/bmj.m3164.

[164] Viknesh Sounderajah, Hutan Ashrafian, Robert M Golub, Shravya Shetty, Jeffrey De Fauw, Lotty Hooft, Karel Moons, Gary Collins, David Moher, Patrick M Bossuyt, Ara Darzi, Alan Karthikesalingam, Alastair K Denniston, Bilal Akhter Mateen, Daniel Ting, Darren Treanor, Dominic King, Felix Greaves, Jonathan Godwin, Jonathan Pearson-Stuttard, Leanne Harling, Matthew McInnes, Nader Rifai, Nenad Tomasev, Pasha Normahani, Penny Whiting, Ravi Aggarwal, Sebastian Vollmer, Sheraz R Markar, Trishan Panch, and Xiaoxuan Liu. Developing a reporting guideline for artificial intelligence-centred diagnostic test accuracy studies: the stard-ai protocol. BMJ Open, 11 (6), 2021. ISSN 2044-6055. doi: 10.1136/bmjopen-2020-047709. URL https://bmjopen.bmj.com/content/11/6/e047709.

[165] Pamela Munguía-Realpozo, Ivet Etchegaray-Morales, Claudia Mendoza-Pinto, Socorro Méndez-Martínez, Ángel David Osorio-Peña, Jorge Ayón-Aguilar, and Mario García-Carrasco. Current state and completeness of reporting clinical prediction models using machine learning in systemic lupus erythematosus: A systematic review. Autoimmunity Reviews, page 103294, 2023. ISSN 1568-9972. doi: https://doi.org/10.1016/j.autrev.2023.103294. URL https://www.sciencedirect.com/science/article/pii/S1568997223000289.

[166] Hussein Ibrahim, Xiaoxuan Liu, and Alastair K. Denniston. Reporting guidelines for artificial intelligence in healthcare research. Clinical & Experimental Ophthalmology, 49(5):470–476, 2021. doi: https://doi.org/10.1111/ceo.13943. URL https://onlinelibrary.wiley.com/doi/abs/10.1111/ceo.13943.

[167] Lukas Folle, Sara Bayat, Arnd Kleyer, Filippo Fagni, Lorenz A Kapsner, Maja Schlereth, Timo Meinderink, Katharina Breininger, Koray Tascilar, Gerhard Krönke, Michael Uder, Michael Sticherling, Sebastian Bickelhaupt, Georg Schett, Andreas Maier, Frank Roemer, and David Simon. Advanced neural networks for classification of MRI in psoriatic arthritis, seronegative, and seropositive rheumatoid arthritis. Rheumatology, 03 2022. ISSN 1462-0324. doi: 10.1093/rheumatology/keac197. URL https://doi.org/10.1093/rheumatology/keac197.keac197.

[168] Vincenzo Venerito, Giacomo Emmi, Luca Cantarini, Pietro Leccese, Marco Fornaro, Claudia Fabiani, Nancy Lascaro, Laura Coladonato, Irene Mattioli, Giulia Righetti, Danilo Malandrino, Sabina Tangaro, Adalgisa Palermo, Maria Letizia Urban, Edoardo Conticini, Bruno Frediani, Florenzo Iannone, and Giuseppe Lopalco. Validity of machine learning in predicting giant cell arteritis flare after glucocorticoids tapering. Frontiers in Immunology, 13, 2022. ISSN 1664-3224. doi: 10.3389/fimmu.2022.860877. URL https://www.frontiersin.org/articles/10.3389/fimmu.2022.860877.

