## Supplemental File Categories for "Understanding the role and adoption of artificial intelligence techniques in rheumatology research: an in-depth review of the literature"

García et al. (2023)

### 1 Classification

#### 1.1 Categories

1. **Disease identification and prediction**: the disease prediction task can be seen as an individualisation of disease classification, in which the classification between healthy (i.e., controls) and unhealthy patients is the main objective. This distinction is usually based on a threshold cutoff point. In this section, studies that attempt to predict the disease status rather than to classify different rheumatic and musculoskeletal diseases (RMDs) are detailed. Predictive models that address early diagnosis are an important branch of this topic since a late diagnosis can be paired with increased flares and organ dysfunction, according to the therapeutic window of opportunity concept previously introduced. The existence of diseases with a similar course, as well as, the lack of specific symptoms, diagnostic criteria, and/or validated biomarkers can hinder the early diagnosis of some diseases, such as axial spondyloarthritis (axSpA) or juvenile idiopathic arthritis, compromising the long-term wellbeing of patients. An important task on this topic is the development of algorithms capable of identifying patients in the electronic health record (EHR), facilitating the construction of specific datasets for epidemiological and observational studies. A fraction of the RMDs are considered to be rare diseases, and some of them have a poor prognosis and an increased risk of death. For this reason, some authors have put their efforts into developing pipelines that take advantage of statistical learning techniques capable of detecting patients, while avoiding manual and time-consuming chart reviews. Hence, natural language processing has become particularly relevant for tackling this task.

2. **Disease classification**: predictive models for disease classification have gained interest due to their ability to discriminate the pathology of a patient among diseases that share a similar course, symptoms, or early manifestations. For example, in this last scenario, the diagnosis of rheumatoid arthritis (RA) or osteoarthritis (OA) may not be clear when starting from an early inflammatory arthritis status, hence, this kind of task often relies on immune or genetic data. Random forest (RF) and support vector machines algorithms have been proposed for this task due to their power, versatility, and ability to handle non-linear data. The relevance of disease classification models lies behind their ability to assist physicians in determining the specific disease of a patient. With the model results, the physicians will follow the most suitable therapeutic strategy for that concrete disease. The choice of inappropriate treatment, due to misdiagnosis, can jeopardise the remission status and maintain or worsen the pathological manifestations produced by the true underlying disease. Furthermore, a delayed diagnosis may displace the therapeutic window of opportunity [1], compromising future outcomes and patient recovery. Therefore, making the correct diagnosis is imperative.

3. **Patient stratification and disease subtype identification**: patient stratification and disease subtype identification have also been interesting areas for rheumatology researchers. The key idea of these studies is to stratify patients into meaningful subgroups that share similar characteristics (e.g., histological, molecular features), and that are different from other subgroups of patients. Therefore, patients who belong to different subgroups can benefit from specific treatments and care, according to the different mechanisms involved in the pathogenesis of the disease. These studies are particularly relevant when the disease to be studied has significant individual variability in terms of clinical, laboratory, and immunological abnormalities. Unsupervised algorithms and clustering techniques, such as hierarchical clustering,

are essential for this task.

Different manifestations of the disease are also considered on this topic. For instance, patients who present a concrete pathology can be prone to the development of different associated pathologies, such as malignancies or cardiovascular events in pSS patients. In this context, the control group is usually made up of patients with the main pathology, but without the associated manifestation. Graph-based techniques have been used to assess the strength of the association between the different predictors to generate prototypical variable profiles capable of discriminating between different disease phenotypes. Other manifestations considered were the different degrees of anatomical damage, since they could serve as a footprint for persistent inflammatory activity. Several indicators or scales have been analysed (i.e., erosions in RA or axSpA; Kellgren and Lawrence (KL) scale in OA). In this situation, radiological data are almost indispensable for quantifying such a measure, and, therefore convolutional neural network models become particularly relevant.

4. **Disease progression and activity**: Forecasting how the disease will evolve, either in terms of disease activity, health status or mortality, can assist doctors in providing vigilant monitoring and focusing resources on the patients most in need. Due to their inherent characteristics, these studies typically have a longitudinal design and predictive models are the most widespread solution to meet this objective, although other unsupervised approaches, such as topic modelling have been also employed. This topic is of particular importance in potentially life-threatening rheumatologic diseases, that is, autoimmune disorders (e.g., RA, SLE). Complex indices [2], are often required to estimate the activity of the disease, such as SLEDAI or CDAI.

5. **Treatment response**: the response variability to treatment among different patients, especially in complex treatments such as biological DMARDs, is currently a promising research challenge. The polygenic response to some of these treatments requires large datasets from which meaningful statistical associations can be identified. Together with the polygenic response, other factors, such as the dosage, are involved in the fluctuating treatment response. Similarly, response to treatment may be quantified differently depending on the disease explored. For example, in RA the treat-to-target approach is commonly used. Moreover, the time horizon of the prediction is variable. While some studies try to predict an outcome from a few months to years after starting the treatment, others try to predict the response before the initiation of the treatment. Data science techniques can be extremely helpful for modelling and describing such statistical associations. For instance, defining a rescue therapy or prescribing an alternative treatment before it is too late can improve the patient's well-being and control the progression of the disease.

   Different research avenues for treatment response can exist. Hence, researchers have addressed different scenarios such as non-responder patients (i.e., drug resistance) or adverse event reactions derived from a medication. The starting dose of treatment is also a research question addressed in treatment response predictive model studies. Finally, studies focused on the patient's perception of treatment are also included in this topic. Since the variety of topics within this category is high, multiple, and heterogeneous artificial intelligence algorithms have been employed.

6. **Predictors identification**: the identification of predictors associated with a dependent variable is the core of many investigations. Usually, this problem is tackled with linear and logistic regression models, as well as, with regularization methods such as Ridge or Lasso regression. On the other hand, decision trees, RF, and other highly interpretable algorithms that provide a variable importance measure are com-

monly chosen options for addressing this task. The identification of predictors has usually been the first step taken to address other research questions, as well as, to build predictive models. In the medical field, the identification of clinical variables that can be related to the development of a disease, a worse prognosis, or a better health-related quality of life is of particular importance. For instance, this task has gained attention in a variety of diseases such as pseudogout, primary Sjögren's syndrome (pSS), or uveitis.

When a study tackles more than one topic, it is assigned according to the main research objective of the AI technique.

#### 1.2 Subcategories

Although a common approach is to enrich the dataset with predictors of different natures (e.g., demographic, clinical, molecular biomarkers or radiological); a distinction between types of predictors and their role in the study is intended. As occurred in previous section, it is sometimes unclear how to classify a study depending on the type of variables involved, as there can be a mix of data sources. Therefore, the following classification was proposed:

1. **Clinical, demographic and biomarker data accessible within routine clinical practice**: clinical and demographic predictors, such as sex, age, diagnoses, treatments, comorbidities, and age of symptom onset, are commonly used to build disease classification predictive models due to their easy accessibility and immediacy in routine clinical practice. Historically, traditional statistics models have benefited from these kinds of predictors, as they were relatively easy to obtain and manipulate. However, they are an essential source of information for conducting epidemiological research with traditional and modern statistical learning approaches. Since some laboratory values (e.g., blood, urine or serum levels), can play a key role in autoimmune diseases such as antibodies titer; biomarkers that are usually collected during routine clinical practice for patient management are also included in this category. The data considered in this category encompass the following sources: billing codes; discharge summaries, radiology reports, ambulatory notes, admission notes, progress notes and pathology reports; disease classification criteria; EHR format-free text notes and structured and codified data; health surveys and health-related quality of life questionnaires; medical and pharmacy claims history; patient-reported outcomes; physical hospital records; public databases; social media data; and sensor and wearable data. With the previous data sources, comorbidity; demographic; disease activity index; diagnosis; functional ability; histological and biopsy data; laboratory data; lifestyle; physical activity; medical procedures; quality of life; and treatment data have been used in the different studies.

2. **Complex molecular biomarkers**: changes in cellular and tissue metabolism and composition may appear because of inflammatory disease. Therefore, the levels of certain metabolites or antibodies may be increased. Differences in these metabolites and in their concentrations can constitute a useful fingerprint to differentiate diseases, acting as a biomarker signature.

   As a result, multiple immunological, histological, and genomic molecular biomarkers have been studied or are being investigated. Since some classical antibodies can be shared between different diseases, such as rheumatoid factor (i.e., RA and pSS) additional and unambiguous biomarkers are desirable to identify concrete pathologies. MicroRNAs, T-cell receptors, aminoacids levels, DNA methylation patterns, type I interferon signature, chemokines, cytokines and proinflammatory molecules may be useful fingerprints for identifying specific diseases and/or their prognosis. A challenge in studies that use these molecular signatures is to face the *high dimensionality problem*, this is, a scenario where the number of potential predictors is much higher than the number of observations (commonly known as $p>>n$). Therefore, preprocessing steps like feature selection play a key role in studies in which this casuistry occurs.

The biomarkers considered in this category include: bone degradation markers; cytokines/chemokines; differentially methylated CpG sites; growth factors; IFN signature; IgG-Gal ratio; matrix metalloproteinases; miRNAs; serum metabolome and lipidome; T-cell receptors; and urinary protein biomarkers.

The acquisition techniques encompass the following ones: bead-based multiplex, individual enzyme, and fluorescent microparticle-based immunoassays; DNA methylation; ELISAs; flow cytometry; gene expression analysis/profiles; genome-wide single nucleotide polymorphism; H nuclear magnetic resonance-based metabolomic; immune phenotypes; nucleic-acid programmable protein arrays; and tandem mass spectrometry.

These data were obtained with the following platform/systems/arrays technologies: 3D-Gene Human miRNA Oligo Chip/Labeling kit; AXIMA Resonance MALDI MS; HumanCoreExome array; Illumina HiSeq 2500 platform; Illumina Human Methylation 450k array (HM450k); Illumina HumanHap550; Human610-Quad arrays; Illumina HumanHT-12-v4 Expression BeadChips; Illumina HumanOmniExpress array; Illumina NextSeq500; Illumina TruSeq RNA; Infinium MethylationEPIC Bead-Chip kit; Kiloplex Quantibody protein array; Luminex Human Magnetic Assay; Meso Scale Discovery electrochemiluminescence assay; NanoString nCounter gene expression; V-PLEX Cytokine Panel 1 Human Kit.

3. **Medical images**: an image is understood as a matrix of pixels, each of them with an intensity value. When working with radiological images, deep learning (DL) is one of the most promising data science techniques due to its ability to capture complex interactions and patterns from the data. Computer vision for image segmentation, image registration, anatomical measurement, and pathological identification and detection are some of the capabilities DL can offer to rheumatology researchers who work with medical images. Computer vision has also been applied as an intermediate step in other research pipelines. For example, DL techniques have been used to segment parts of the body that could be used in a subsequent step of a classification model. The structure and topology of the network differ between studies. Parameters and hyperparameter optimisation, such as the number of hidden layers, the number of neural units, the penalisation and regularisation (e.g., data augmentation, dropout), the learning rate, the activation and loss functions, or the number of epochs, define the complexity of the networks. Therefore, the performance of the final model can be highly determined by the preprocessing steps, the network infrastructure, and its parameters. Moreover, the use of previously pre-trained networks and transfer learning is also a well-extended option, which facilitates the use and implementation of this promising technique, reducing the training time and the number of training examples.

   Depending on the methodology followed by the researcher, radiological information can be used alone or complement other demographic and clinical characteristics in ensemble methods. DL is usually preferred when the image is used solely to train the model, in fact, already published results have demonstrated that algorithms trained exclusively on image data are robust enough to provide remarkable results. However, when relevant radiological information is extracted from the image after a feature selection process (i.e., a radiologist measure and quantification, the distance between structures, histogram features, etc.), and algorithms are trained with these data combined with other clinical and demographic parameters, machine learning (ML) classifiers can be used. Research studies combining DL and ML techniques for training and benchmarking algorithms have been published [3]. In these studies, the performance of the algorithms is usually compared to the radiologist's criterion.

Due to the key role that radiological images play in the diagnosis and staging of certain diseases such as knee OA or osteoporosis, many of the RMDs research studies employing DL focus on these diseases. However, research on other diseases can also be carried out with the aid of such techniques.

Finally, the data source variability of medical imaging-based studies is high. Apart from well-extended image techniques such as computed tomography, magnetic resonance, X-rays, ultrasound (US); other less common image techniques such as optoacoustic [4], photomicrographs [5], smartphone photos [3], and thermograms [6] have also been employed in research studies.

The classification proposed could have also considered data from sensors, wearables, and activity trackers (e.g., accelerometer and gyroscope), this is, sensor and signal data. However, the number of articles in which this type of data is used is limited. Therefore, these studies were discussed in the *Clinical and demographic* category.

If a study combines predictors that fall into more than one of the subcategories, the article is classified according to the most "complex" variable. For instance, if a research study considers clinical variables and only a concrete genotype (i.e., ABCB1 genotypes) not collected during routine clinical practice, the article will be classified into the "Complex molecular biomarkers" subcategory. On the other hand, studies that use radiological measures, but not the image itself, together with clinical variables will be classified into the "Clinical, demographic and biomarker data accessible within routine clinical practice" subcategory.

To conclude this introduction, as an example of dataset enrichment with different data sources, authors in [7] combined 82, clinical, demographic, laboratory and image measures (i.e., US) to predict intravenous immunoglobulin resistance in Kawasaki disease using different ML models.

#### References


[1] Leonie E Burgers, Karim Raza, and Annette H van der Helm - van Mil. "Window of opportunity in rheumatoid arthritis – definitions and supporting evidence: from old to new perspectives". In: *RMD Open* 5.1 (2019). DOI: 10.1136/rmdopen-2018-000870. eprint: https://rmdopen.bmj.com/content/5/1/e000870.full.pdf. URL: https://rmdopen.bmj.com/content/5/1/e000870.

[2] Sociedad Española de Reumatología. *Activity indices, questionnaires and other measurement instruments in Rheumatology.* 2021. URL: https://www.ser.es/profesionales/que-hacemos/investigacion/herramientas/catalina/.

[3] Mark Reed, Timothy Le Souëf, and Elliot Rampono. "A pilot study of a machine-learning tool to assist in the diagnosis of hand arthritis". In: *Internal Medicine Journal* 52.6 (2022), pp. 959–967. DOI: https://doi.org/10.1111/imj.15173. eprint: https://onlinelibrary.wiley.com/doi/pdf/10.1111/imj.15173. URL: https://onlinelibrary.wiley.com/doi/abs/10.1111/imj.15173.

[4] Suhanyaa Nitkunanantharajah, Katja Haedicke, Tonia B. Moore, et al. "Three-dimensional optoacoustic imaging of nailfold capillaries in systemic sclerosis and its potential for disease differentiation using deep learning". In: *Scientific Reports* 10.1 (2020), p. 16444. ISSN: 2045-2322. DOI: 10.1038/s41598-020-73319-2. URL: https://doi.org/10.1038/s41598-020-73319-2.

[5] Vincenzo Venerito, Orazio Angelini, Gerardo Cazzato, et al. "A convolutional neural network with transfer learning for automatic discrimination between low and high-grade synovitis: a pilot study". In: *Internal and Emergency Medicine* 16 (6 2021), pp. 1457–1465. ISSN: 1970-9366. DOI: 10.1007/s11739-020-02583-x. URL: https://doi.org/10.1007/s11739-020-02583-x.

[6] Shawli Bardhan and Mrinal Kanti Bhowmik. "2-Stage classification of knee joint thermograms for rheumatoid arthritis prediction in subclinical inflammation". In: *Australasian Physical & Engineering Sciences in Medicine* 42.1 (2019), pp. 259–277. ISSN: 1879-5447. DOI: 10.1007/s13246-019-00726-9. URL: https://doi.org/10.1007/s13246-019-00726-9.

[7] Tengyang Wang, Guanghua Liu, and Hongye Lin. "A machine learning approach to predict intravenous immunoglobulin resistance in Kawasaki disease patients: A study based on a Southeast China population". In: *PLOS ONE* 15.8 (Aug. 2020), pp. 1–15. DOI: 10.1371/journal.pone.0237321. URL: https://doi.org/10.1371/journal.pone.0237321.
